# Integrative multi-omics analysis of childhood aggressive behavior

**DOI:** 10.1101/2021.09.13.21263063

**Authors:** Fiona A. Hagenbeek, Jenny van Dongen, René Pool, Peter J. Roetman, Amy C. Harms, Jouke Jan Hottenga, Cornelis Kluft, Olivier F. Colins, Catharina E.M. van Beijsterveldt, Vassilios Fanos, Erik A. Ehli, Thomas Hankemeier, Robert R. J. M. Vermeiren, Meike Bartels, Sébastien Déjean, Dorret I. Boomsma

## Abstract

This study introduces and illustrates the potential of an integrated multi-omics approach in investigating the underlying biology of complex traits such as childhood aggressive behavior. In 645 twins (cases=42%), we trained single- and integrative multi-omics models to identify biomarkers for subclinical aggression and investigated the connections among these biomarkers. Our data comprised transmitted and two non-transmitted polygenic scores (PGSs) for 15 traits, 78772 CpGs, and 90 metabolites. The single-omics models selected 31 PGSs, 1614 CpGs, and 90 metabolites, and the multi-omics model comprised 44 PGSs, 746 CpGs, and 90 metabolites. The predictive accuracy for these models in the test (*N*=277, cases=42%) and independent clinical data (*N*=142, cases=45%) ranged from 43% to 57%. We observed strong connections between DNA methylation, amino acids, and parental non-transmitted PGSs for ADHD, Autism Spectrum Disorder, intelligence, smoking initiation, and self-reported health. Aggression-related omics traits link to known and novel risk factors, including inflammation, carcinogens, and smoking.

## Introduction

Omics studies can lead to an improved understanding of the biological mechanisms contributing to mental health and disorders (Jakovljevic and Jakovljevic 2019). Different omics technologies, e.g., genomics, epigenomics, transcriptomics, or metabolomics, assess different aspects of the development and progression of complex traits and disorders. The analysis of a single omics layer provides a unique, but likely incomplete, picture of the underlying biology, whereas studies combining multiple omics layers may lead to more comprehensive insights into human biology, because the different omics layers are interrelated and interact (Wörheide et al. 2021). Multi-omics analyses can aid in biomarker discovery, diagnosis, patient classification or subtyping, evaluation of treatment response and uncovering novel insights into disease biology (Pinu et al. 2019; Subramanian et al. 2020).

Here, we present an integrative multi-omics analysis of childhood aggressive behavior. Human aggression is a complex and heterogenous behavior encompassing hostile, destructive, or injurious behavior aimed at causing physical or emotional harm to others (Anderson and Bushman 2002; Siever 2008). In several disruptive behavioral disorders, such as conduct and oppositional defiant disorders or intermittent explosive disorder, inappropriate levels of aggressive behavior are observed (Radwan and Coccaro 2020). In humans, high co-occurrence with other social, behavioral, and emotional problems is reported (Bartels et al. 2018; Whipp et al. 2021a). Childhood aggressive behavior puts a burden on children and their parents/caretakers, and is predictive of multiple adverse outcomes later in life, such as antisocial personality disorder (Whipp et al. 2019), criminal convictions (Kassing et al. 2019), lower educational attainment (Vuoksimaa et al. 2020), or negative interpersonal relationships (Fergusson et al. 2005).

Large-scale genome-wide association (GWA) studies on aggression have not yet identified significant single nucleotide polymorphisms (SNPs) (Odintsova et al. 2019; Ip et al. 2021). Gene-based analysis for childhood aggression yielded three significantly associated genes (*ST3GAL3, PCDH7*, and *IPO13*) (Ip et al. 2021). The polygenic score for childhood aggression does not only significantly explain childhood aggressive behavior at age 7 (Ip et al. 2021), but it also associated with aggression at age 12 to 41 in a Dutch sample and age 38 to 48 in an Australian sample (van der Laan et al. 2021). Small-scale epigenetic studies (sample size range: 41-260) showed that DNA methylation differences in various tissues are associated with aggression and related traits in children and adults (Guillemin et al. 2014; Cecil et al. 2018b, a; Mitjans et al. 2019). The first large-scale epigenome-wide association study (EWAS) meta-analysis across child and adult cohorts (N = 15,324) reported 13 significant sites in peripheral blood for broad aggression across the lifespan (van Dongen et al. 2021). Metabolomics studies (sample size range: 77-725) detected plasma and serum metabolites associated with aggression and related traits in adults (Gulsun et al. 2016; Chen et al. 2020; Whipp et al. 2021b). A study in 1,347 twins and 183 clinical cases found significant associations of urinary metabolites with childhood aggression (Hagenbeek et al. 2020). These single omics approaches hint at potentially important biological pathways for human aggression, with most of these findings awaiting replication.

In this study, we aim to integrate multiple omics layers to construct a multi-omics biomarker panel for childhood aggressive behavior and explore the correlations among the omics traits included in this panel. We collected biological samples in a subproject of ACTION (Aggression in Children: Unraveling gene-environment interplay to inform Treatment and InterventiON strategies): the ACTION Biomarker Study (Boomsma 2015; Bartels et al. 2018; Hagenbeek et al. 2020). The ACTION Biomarker Study comprised a cohort of twins from the Netherlands Twin Register (NTR) (Ligthart et al. 2019) and a clinical cohort of children referred to a youth psychiatry clinic (LUMC-Curium, the Netherlands). The genome-wide SNP and DNA methylation (Illumina EPIC 850K array) data, and the urinary amines and organic acids as measured in these cohorts were previously included in a genome-wide genetic and epigenetic association study and a metabolomics study of (childhood) aggression (Hagenbeek et al. 2020; van Dongen et al. 2021; Ip et al. 2021). We expand on these single-omics studies by integrating these data with a third metabolomics dataset, focusing on urinary steroid hormones. We calculated transmitted and non-transmitted polygenic scores (PGSs) for childhood aggression and a series of genetically correlated traits, such as Attention-Deficit Hyperactivity Disorder (ADHD), smoking, and intelligence. The non-transmitted PGSs assess the indirect effects of genetic variants that were not transmitted from parents to offspring, i.e., genetic nurture, and capture the effects of the environment created by parents beyond the genetic intergeneration transmission (Bates et al. 2018; Kong et al. 2018). Thus, if alleles not transmitted from parents to offspring affect offspring outcomes, this indicates that the offspring’s home-environment is influenced by parentental genotypes, and the home-environment in turn affects offspring outcomes (Branje et al. 2020).

We employed an analytical design comprising three phases: 1) single-omics analyses; 2) pairwise cross-omics analyses; and 3) multi-omics analyses (**Fig. 1**) (Duruflé et al. 2020). First, we built single-omics biomarker panels in the twin cohort, with 70% of the twin data for model training, 30% of the twin data for model testing, and an independent clinical cohort for follow-up. Second, we examined the pairwise cross-omics connections of those omics traits selected by the single-omics models in the training data. Third, using the same data split for model training and testing, we compared three multi-omics models, with different assumptions on the correlations among the omics traits and describe the multi-omics connections of the selected omics traits.

**Fig. 1.**
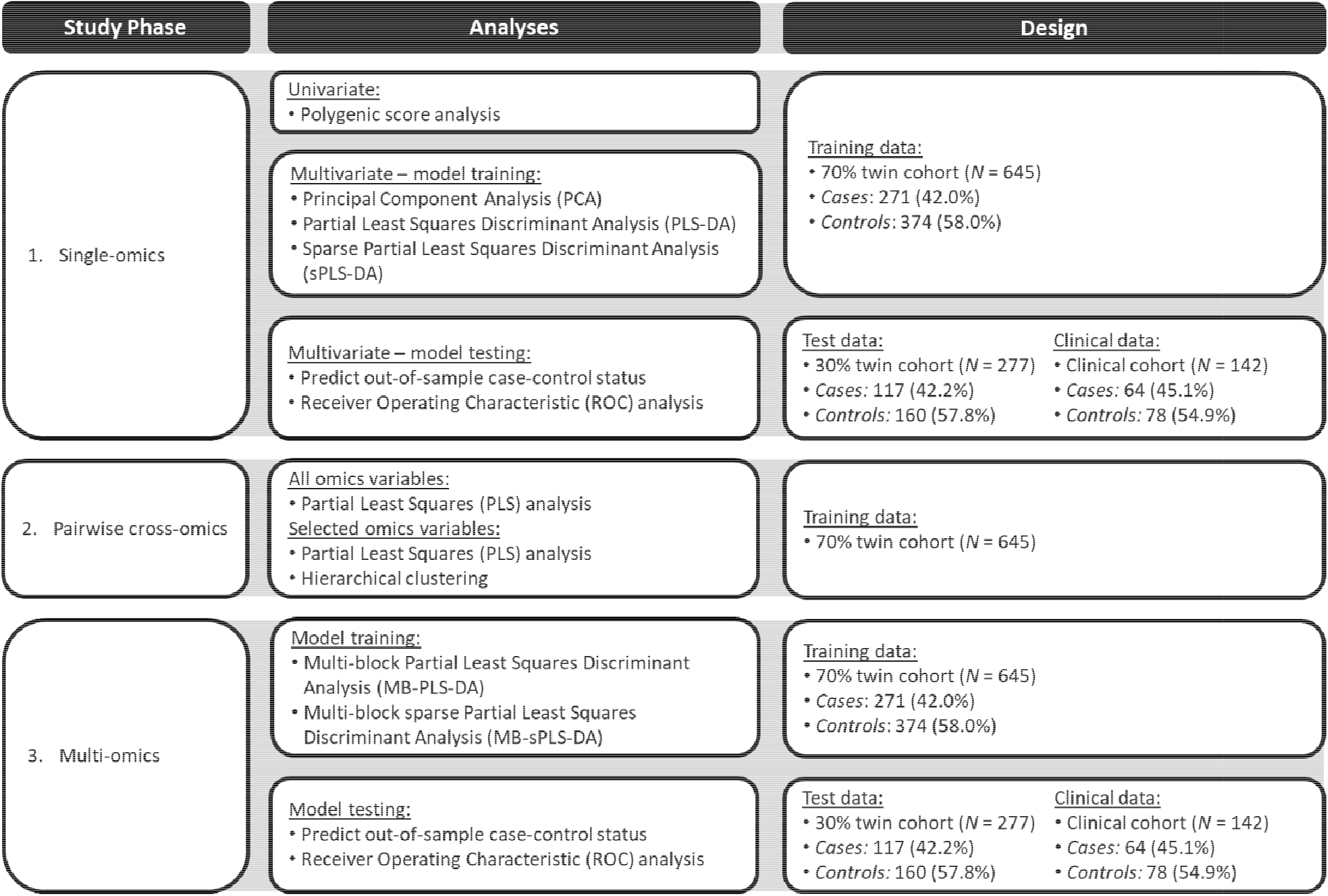
Overview of the biomarker identification approach for childhood aggression—details of statistical analyses and data included in each analysis. We employed an analytical design comprising three phases: 1) single-omics analyses; 2) pairwise cross-omics analyses; and 3) multi-omics analyses. First, we performed univariate polygenic score (PGS) analysis in 70% of the twin data and built multivariate single-omics biomarker panels in the twin cohort, with 70% of the twin data for model training (training data), 30% of the twin data and the clinical cohort for model testing (test data). Second, we examined the overall pairwise cross-omics correlations and the pairwise correlations of those omics variables selected by the single-omics models in the training data. Third, using the same data, we compared three multi-omics models, with different assumptions of the correlations among the omics blocks, and describe the multi-omics relationships of the selected omics variables. We offer the analytical details in the **Methods** section.

## Materials and methods

### Study population and procedures

Participants included 1,494 twins (747 complete pairs) from the Netherlands Twin Register (Ligthart et al. 2019), and 189 children referred to the LUMC-Curium youth psychiatry clinic in the Netherlands that took part in the ACTION Biomarker Study (Aggression in Children: Unraveling gene-environment interplay to inform Treatment and InterventiON strategies) (Boomsma 2015; Bartels et al. 2018; Hagenbeek et al. 2020). Both cohorts collected first-morning urine samples and buccal-cell swabs with standardized protocols (for details, see **Appendix A**). In the twin cohort, we also collected buccal-cell swabs from parents and siblings of the twins. The current study included participants if they had aggression status, complete omics data for all three omics layers, and all relevant covariates (**Table S1**), as the current software does not allow for missing data. Parents provided written informed consent for their children and twin parents provided written informed consent for their own participation. Study approval was obtained from the Central Ethics Committee on Research Involving Human Subjects of the VU University Medical Center, Amsterdam (NTR 25th of May 2007 and ACTION 2013/41 and 2014.252), an Institutional Review Board certified by the U.S. Office of Human Research Protections (IRB number IRB00002991 under Federal-wide Assurance-FWA00017598; IRB/institute codes), and the Medical Ethical Committee of Leiden University Medical Center (B17.031, B17.032 and B17.040).

### Aggressive behavior

Parents and teachers rated aggressive behavior on the Aggressive Behavior syndrome scale of the Achenbach System of Empirically Based Assessment (ASEBA) Child Behavior Checklist (CBCL) or Teacher Report Form (TRF) (Achenbach et al. 2017). We have described selection and definition of aggression cases and controls previously (Hagenbeek et al. 2020). In brief, we selected twin pairs based on concordance (case-case or control-control) and discordance (case-control pairs) for mother-(93%) or teacher-rated (7%) aggressive behavior at ages 3, 7, and/or 10 years. We matched concordant control pairs on postal code to the case-case and case-control pairs, and collection of biological samples within regions in the Netherlands was around the same time. Cases were defined by mother-or teacher-rated sex-and age-specific T-scores of 65 or higher (subclinical levels), or on mother-rated age-specific thresholds on the item scores at age 3 (≥ 13), age 7 (≥ 5), or age 10 (≥ 4) (*N* = 388, **Table S1**). We denoted individuals with scores below these thresholds as controls (*N* = 534, **Table S1**). In the clinical cohort, children with a parent-rated sex-specific T-score of ≥ 70 (clinical levels) were classified as cases (*N* = 64), and children with T-scores of < 65 were classified as low scoring controls (*N* = 78, **Table S1**). We excluded children with T-scores in the subclinical range (T-scores of ≥ 65 and < 70) from this study (*N* = 35).

### Omics measurements

Detailed information on omics measurements is included in **Appendix B**. Genotyping was performed on Affymetrix Axiom or Illumina GSA arrays (Ehli et al. 2017; Beck et al. 2019), and genome-wide SNP data were available for 3,334 participants, including 1,702 parents and siblings of twins (AXIOM = 909, GSA = 2,425). Transmitted and two non-transmitted polygenetic scores (PGS) were calculated for childhood aggression and 14 other traits that showed a significant (*p* < 0.02) genetic correlation of ≤ -0.40 or ≥ 0.40 with childhood aggression (Ip et al. 2021) (**Table 1**). Thus, in total we calculated 45 PGSs: a transmitted, non-transmitted maternal, and non-transmitted paternal PGS for each trait. The effects of sex, age at biological sample collection, genotyping platform, and the first 10 genetic principal components (PCs) were regressed on the standardized PGSs and we included residuals in the analyses.

**Table 1.**
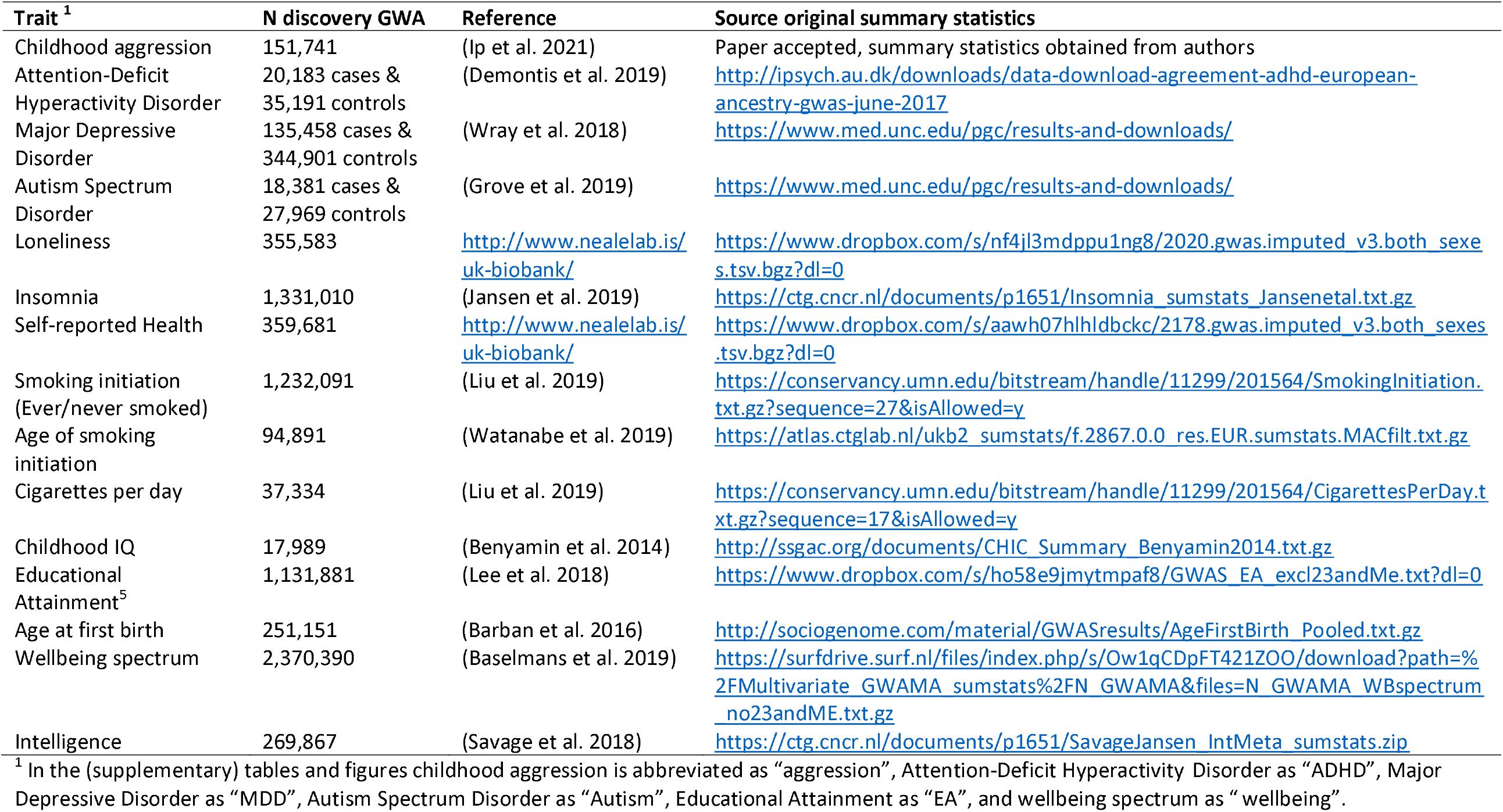
Overview of the discovery genome-wide association studies to calculate polygenetic scores.

Genome-wide DNA methylation was measured on the Infinium MethylationEPIC BeadChip Kit (Illumina, San Diego, CA, USA (Moran et al. 2016)). Quality Control (QC) and normalization were carried out with pipelines developed by the Biobank-based Integrative Omics Study (BIOS) consortium (Sinke et al. 2019). From the 787,711 autosomal methylation probes that survived QC, the top 10% most variable probes were included in the analyses (**Data S1**). Residual methylation levels were obtained by regressing the effects of sex, age, percentages of epithelial and natural killer cells, EPIC array row, and bisulfite sample plate, from the methylation β-values.

Urinary metabolomics data were generated on: 1) a liquid chromatography mass spectrometry (LC-MS) platform targeting amines; 2) a LC-MS platform targeting steroid hormones; and 3) a gas chromatography (GC) MS platform targeting organic acids. We excluded metabolites with a relative standard deviation of the QC samples larger than 15% and retain 60 amines, 10 steroids, and 20 organic acids in our analyses. After QC, we normalized metabolite levels to the sample-median and inverse normal rank transformed. We analyzed residuals obtained by regressing the effects of sex and age on the normalized and transformed urinary metabolites.

### Statistical analyses

To define multi-omics biomarker panels capable of discriminating between cases and controls and explore the connections among the omics traits included in these panels, we employed an analytical design comprising three phases: 1) single-omics analyses; 2) pairwise cross-omics analyses; and 3) multi-omics analyses (**Fig. 1**). To avoid overfitting of the single-and multi-omics models, we randomly split the twin sample at the twin pair level into two subsets: 70% of the data for model training (training data), and 30% of the data for model testing (test data; **Table 2**). The clinical validity of the final single- and multi-omics models was evaluated in the clinical cohort (**Table 2**). We carried analyses out in the mixOmics R package (version 6.12.1) implemented in the R programming language (version 4.0.2) (R Core Team; Rohart et al. 2017).

**Table 2.** Demographics of the training, test, and clinical data.

|  | Training data (70% twin cohort) |  |  | Test data (30% twin cohort) |  |  | Clinical data (Clinical cohort) |  |  |
| --- | --- | --- | --- | --- | --- | --- | --- | --- | --- |
|  | Controls | Cases | Total | Controls | Cases | Total | Controls | Cases | Total |
| *N* (%) | 374<br>(58.0%) | 271<br>(42.0%) | 645 (100%) | 160<br>(57.8%) | 117<br>(42.2%) | 277 (100%) | 78 (54.9%) | 64 (45.1%) | 142 (100%) |
| *N* (%) complete twin pairs | 128 | 80 | 293 | 56 | 35 | 128 | - | - | - |
| Mean (*SD*) age | 9.4 (1.9) | 9.6 (1.8) | 9.5 (1.9) | 9.6 (1.9) | 9.6 (1.9) | 9.6 (1.9) | 10.8 (1.7) | 9.6 (1.7) | 10.2 (1.8) |
| Range age | 6.1 - 12.7 | 6.1 - 12.9 | 6.1 - 12.9 | 5.6 - 12.6 | 5.8 - 12.8 | 5.6 - 12.8 | 6.5 - 13.4 | 6.3 - 13.3 | 6.3 - 13.4 |
| *N* (%) females | 199<br>(53.2%) | 116<br>(42.8%) | 315<br>(48.8%) | 81 (50.6%) | 55 (47.0%) | 136<br>(49.1%) | 20 (25.6%) | 19 (29.7%) | 39 (27.5%) |
| *N* (%) MZ twins | 299<br>(80.0%) | 228<br>(84.1%) | 527<br>(81.7%) | 118<br>(73.7%) | 108<br>(92.3%) | 226<br>(81.6%) | - | - | - |
| Mean (*SD*) aggression score <sup>1</sup> | 3.3 (4.1) | 7.3 (5.8) | 5.0 (5.3) | 3.1 (4.0) | 7.6 (6.6) | 5.0 (5.7) | 6.1 (3.4) | 21.1 (4.8) | 12.9 (8.5) |

Notes: MZ, monozygotic.
<sup>1</sup> Measured with the mother-rated Aggressive Behavior syndrome scale of the Achenbach System of Empirically Based Assessment (ASEBA) Child Behavior Checklist (CBCL). The ASEBA CBCL Aggressive Behavior scores in the clinical cohort include 90% mother report and 10% father report.

#### Phase 1: Single-omics analyses

##### Univariate polygenic score analyses

In the training data, we first associated the transmitted- and non-transmitted PGSs with childhood aggression through generalized estimating equation (GEE) models. GEE models tested the association of each transmitted and non-transmitted PGS separately on the continuous mother-rated sum scores of the ASEBA CBCL Aggressive Behavior syndrome scale as assessed at the time of biological sample collection. All models included sex, age, genotype array, and the first 10 genetic principal components as covariates. We corrected for the correlation structure within families by using the “exchangeable” correlation structure, obtaining robust variance estimators (Rogers and Stoner 2016). A False Discovery Rate (FDR) of 5% for 45 PGSs was used to correct for multiple testing (p.adjust function in R), setting the significance threshold to *q* ≤ 0.05, i.e., 5% of the significant results will be false positives (Benjamini and Hochberg 1995).

##### Multivariate single-omics models

To get a first insight into the dimensionality of the metabolomics data (90 variables), DNA methylation data (78,772 variables), and the PGSs (45 variables), we ran Principal Component Analysis (PCA) within each omics block in the training data (**Table S2**). To assess the ability of each of the three omics layers to correctly classify aggression status, we applied Partial Least Square Discriminant Analysis (PLS-DA) in the training data. PLS-DA involves iteratively constructing successive latent components, where each component is a linear combination of the included omics variables (Rohart et al. 2017). For each component, PLS-DA aims to maximize the covariance between the residual X matrix, containing the omics data, and the Y matrix, containing the sample classification (i.e., case-control status coded as a dummy variable). The mixOmics software requires a user-defined maximum number of components in PLS-DA models. We chose this maximum based on the number of PCs as determined by the elbow method in the PCA (**Fig. S1; Table S2**). To find the optimal number of components to keep in each PLS-DA model, we employed 10-fold cross-validation (CV) with 100 repeats (*perf* function; **Table S3; Fig. S2**).

Next, we applied sparse PLS-DA (sPLS-DA) to reduce the number of variables in each omics block contributing to each component. sPLS-DA includes Least Absolute Shrinkage and Selection Operator (LASSO) penalization (e.g., *L1* penalization). LASSO shrinks the coefficients of less importance, often highly correlated, variables to zero, removing these variables from the model (Tibshirani 1996). Thus, for each component, sPLS-DA finds the maximum covariance between the residual X matrix containing a subset of the omics data with non-zero coefficients (variable selection) and the Y matrix (Lê Cao et al. 2011). We assessed the variable selection via 10-fold CV with 100 repeats (*tune* function), keeping at least two components in the final model (**Table S3; Fig. S3**). CV again obtained the performance of the final sPLS-DA model using the *perf* function (10 folds, 100 repeats; **Table S3; Fig. S4**).

The ability of the final single-omics models to predict out-of-sample case-control status was evaluated in the test and clinical datasets (*predict* function). For each new observation in the test and clinical datasets, this function calculates the predicted class (case/control) by estimating their predicted dummy variable (of the case-control status) using the maximum, Mahalanobis, or Centroids (Euclidian) distance (see Rohart et al. 2017). When using the maximum distance, the predicted class of a new observation is the class for which we observed the largest predicted dummy value. Both the Mahalanobis and centroids distances are centroid-based distances that predict the class of a new observation, so that the distance between its centroid and predicted scores is minimal. To predict out-of-sample case-control status, we used the best performing prediction distance, as was determined during model training (see **Table S3**).

The misclassification rates of the models, that combine the number of cases classified as controls (false negative rate) and the number of controls classified as cases (false positive rate), was used to evaluate how well the final models predicted case-control status. We employed a balanced misclassification rate, the balanced error rate (BER), that corrects for imbalances in the number of cases and controls. We used a confusion matrix, comparing the true cases and controls with the predicted cases and controls, to calculate the sensitivity (number of cases correctly classified as cases [true positive rate]), specificity (number of controls correctly classified as controls [true negative rate]), and accuracy (overall correct classification). As an alternative, Receiver Operating Characteristic (ROC) analysis assessed the Area Under the Curve (AUC) in both the test and clinical data. We obtained the ROC curve per component.

#### Phase 2: Pairwise cross-omics analyses

To highlight pairwise cross-omics relationships (i.e., DNA methylation-metabolomics, PGSs-metabolomics, and PGSs-DNA methylation), Partial Least Squares (PLS) regression models were constructed in canonical mode in the training data. Similar to canonical correlation analysis, canonical PLS regression aims to find linear combinations of the variables (canonical variates) to reduce the dimensionality of the data while maximizing the covariance between the variates (Rohart et al. 2017). CV has not yet been implemented for PLS in canonical mode. Consequently, we kept the smallest number of components as kept in either of the respective single-omics models (**Table S3**). Therefore, we included 3 components for the DNA methylation-metabolomics model, and 2 components for the PGS-metabolomics and PGS-DNA methylation models.

The PLS models were run with two different sets omics variables, 1) a PLS model that included all 45 PGSs, 78,772 CpGs, and 90 metabolites (model 1), and 2) a PLS model that only included the 36 PGSs, 1,614 CpGs, and 90 metabolites that were selected by the single-omics sPLS-DA models (model 2; **Data S2**). Model 1 provides insight into the connections among all omics variables, while model 2 provides insight into the connections among those omics variables that best contribute to aggression case-control classification in the single-omics sPLS-DA models. In the remainder of the text we will refer to these connections as ‘correlations’, but note that these are not Pearson correlations between among omics variables. These connections are the correlations between the projected variables onto the space spanned by the components as retained in the analysis, i.e., the correlation among the PLS variates (González et al. 2012). On the omics variables included in model 2, we performed hierarchical clustering with the Ward linkage algorithm on Euclidean distances of the PLS variates and used the ‘dendextend’ R-package (Galili 2015) to extract the two largest clusters for both of the omics blocks included in the PLS models.

#### Phase 3: Multi-omics analyses

The multi-omics analysis was conducted through Data Integration Analysis for Biomarker discovery using Latent cOmponents (DIABLO) in the training data. DIABLO extends PLS-DA to multi-block PLS-DA (MB-PLS-DA), that aims at identifying correlated variables from multiple omics blocks that maximize the sample classification (Singh et al. 2019). The method requires a user-defined ‘design matrix’, that specifies the expected correlations among the omics blocks. The symmetric design matrix has the number of rows and columns equal to the number of omics blocks (i.e., 3), and contains values between 0 and 1. A ‘full’ design matrix denotes strong positive correlations among the omics blocks and sets the values among omics blocks close to or equal to one. A ‘null’ design matrix denotes weak or no correlations among omics blocks by setting values close to or equal to zero. The full design matrix optimizes correlations among the omics blocks, while the null design matrix optimizes the discrimination between samples (Rohart et al. 2017; Singh et al. 2019; Duruflé et al. 2020). We can also specify a design matrix with the empirical correlations among the omics blocks.

We compared a multi-omics model with an empirical design matrix (based on the correlations obtained from the model 1 pairwise cross-omics PLS models; **Table S4**) to models with a null or full design matrix. Based on the results of the single-omics sPLS-DA models (**Table S5**), we chose the maximum number of components to include in the MB-PLS-DA models. We determined the optimal number of components to keep in the MB-PLS-DA model with 10-fold CV and 100 repeats (*perf* function; **Fig. S5**). We assessed the variable selection per component per omics block via 5-fold CV with 50 repeats (*tune* function; **Table S5; Fig. S6**). Performance of the final MB-sPLS-DA model was assessed with 5-fold CV (50 repeats; **Table S5; Fig. S7**).

The ability of the final multi-omics models to predict out-of-sample case-control status was evaluated in the test and independent clinical data (*predict* function), based on the best performing prediction distance as was determined during model training (see **Table S5**). The final multi-omics models were evaluated by their balanced error rates (BER), and the sensitivity, specificity, and accuracy of the models were calculated from their confusion matrices. In the multi-omics models, we calculated the ROC curves per component for each omics block.

### Biological characterization

To facilitate biological interpretation, we describe the connections of the PGSs, CpGs, and metabolites, that were selected by the single-omics sPLS-DA models and those selected by the multi-omics MB-sPLS-DA models. For the multi-omics MB-sPLS-DA models we also identified correlation patterns that included high absolute correlations (|*r*| ≥ 0.60) between omics traits of at least two omics blocks. As for the pairwise cross-omics models, these correlations comprise the correlations among the PLS variates. To construct the correlational patterns we denoted all instances where multiple omics variables were connected through bi-or trivariate correlations as a pattern. An example of a pattern comprising one variable of each omics block: high correlation of a PGS with a CpG and a high correlation of this CpG with a metabolites.

To test for enrichment of methylation sites previously associated with other traits, we performed trait enrichment analysis for all traits (619) in the EWAS atlas on the 18^th^ of June 2021 (Xiong et al. 2022). CpGs served as input for the trait enrichment analysis if 20 or more unique CpGs were selected into the single-omics sPLS-DA model, the multi-omics MB-sPLS-DA models, or included in a multi-omics MB-sPLS-DA high correlation pattern. When fewer than 20 CpGs were selected, we manually retrieved the trait associations with the CpGs from the EWAS atlas. Similarly, we performed trait enrichment analysis or manual retrieval of the CpGs included in the clusters as identified for the pairwise cross-omics analyses for all traits (618) in the EWAS atlas on the 1^st^ of July 2021.

## Results

### Polygenic prediction

Transmitted and non-transmitted PGSs for childhood aggression and 13 other traits were individually not significantly associated with aggressive behavior after multiple testing correction (**Data S3**). The transmitted PGS for ADHD were significantly associated with aggressive behavior *(ß* = 1.16, *SE* = 0.26, *q* = 0.0003), while the non-transmitted PGSs were not (mother: *β* = -0.22, *SE* = 0.24, *q* = 0.83; father: *β* = 0.02, *SE* = 0.23, *q* = 0.98), showing that genetic liability for ADHD associates with increased levels of aggressive behavior.

### Single-omics models for childhood aggression

We built cross-validated single-omics biomarker panels for childhood aggressive behavior based on sPLS-DA models including PGSs, DNA methylation, or metabolomics data. The final models comprised 11 transmitted and 25 non-transmitted PGSs, 1,614 CpGs, and all 90 metabolites (**Table S3; Data S2; Fig. S8**). The PGSs selected in the sPLS-DA model comprised the transmitted PGSs for aggression, ADHD, age at first birth, age at smoking initiation, number of cigarettes per day, educational attainment (EA), insomnia, loneliness, Major Depressive Disorder (MDD), smoking initiation, and wellbeing. The non-transmitted maternal PGSs were selected for aggression, ADHD, age at first birth, age at smoking initiation, Autism Spectrum Disorder (ASD), childhood IQ, number of cigarettes per day, EA, insomnia, intelligence, loneliness, MDD, self-reported health, smoking initiation, and wellbeing, and the non-transmitted paternal PGSs were selected for aggression, ADHD, ASD, childhood IQ, number of cigarettes per day, intelligence, MDD, self-reported health, smoking initiation, and wellbeing (**Data S2**). Trait enrichment analyses for all 1,614 selected CpGs in the sPLS-DA model showed the strongest enrichment for glucocorticoid exposure (i.e., administration of corticosteroid medication (Braun et al. 2019); OR = 18.02, *p* = 5.34×10^−158^), and household socioeconomic status in childhood (OR = 9.88, *p* = 1.18×10^−13^; **Table S6**). All three single-omics models showed low classification accuracy in the test and clinical data (**Table S7; Fig. S9-S10**).

### Pairwise cross-omics models

#### DNA methylation-metabolomics

In the pairwise DNA methylation-metabolomics model, we used hierarchical clustering and found two clusters of CpGs and of metabolites (**Fig. 2a; Data S4**). The DNA methylation cluster 1 contains 1,151 (71.3%) of the CpGs selected by the sPLS-DA models for childhood aggression, and showed the strongest trait enrichments for household socioeconomic status in childhood (OR = 12.48, *p* = 4.20×10^−14^; Table S8). The 463 (28.7%) CpGs included in cluster 2 showed the strongest trait enrichment for glucocorticoid exposure (OR = 51.25, *p* ≤ 1.00×10^−308^; **Table S8**). Metabolite cluster 1 contains 69 (76.7%) of the metabolites, including 55 amines, 7 organic acids and 7 steroids, while metabolite cluster 2 contains 21 metabolites (23.3%), including 5 amines, 13 organic acids, and 3 steroids. The average correlation between the DNA methylation and metabolomics blocks, i.e., the correlation among all PLS variates of all components simultaneously, was 0.18 (*q* = 6.07×10^−15^). We observed the highest absolute correlations between metabolites of cluster 1 and CpGs included in cluster 2 (**Fig. 2a**; **Data S4-S5**). Specifically, the amines 3-methoxytyramine (*r* M = -0.21, *r* range: - 0.23 to 0.20), asymmetric dimethylarginine (ADMA, *r* M = -0.20, *r* range: -0.21 to -0.20), L-glutamic acid (*r* = -0.20), L-phenylalanine (*r* M = -0.20, *r* range: -0.21 to -0.20), O-acetyl-L-serine (*r* = -0.20), and symmetric dimethylarginine (SDMA, *r* M = -0.21, *r* range: -0.22 to -0.20) show negative correlations with these CpGs, indicating that increased levels of these urinary metabolites associated with hypomethylation at cluster 2 CpG sites.

**Fig. 2.**
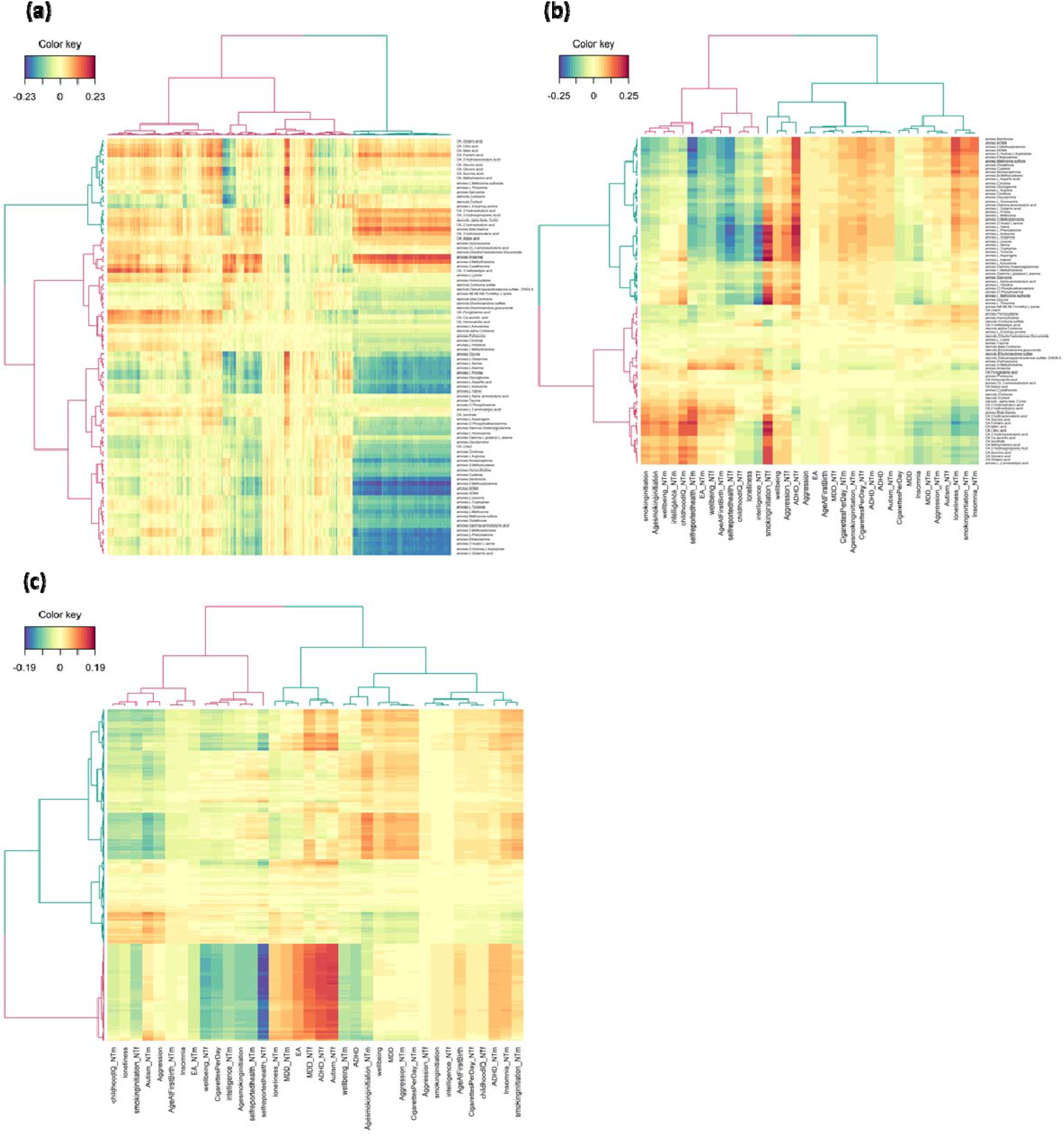
Clustered heatmaps of the relationships obtained by the pairwise cross-omics Partial Least Squares (PLS) regression models including only the omics variables as selected by the single-omics sparse Partial Least Squares Discriminant Analyses (sPLS-DA). We generated the hierarchical clustering using the Ward linkage algorithm on Euclidean distances of the PLS variates. For each dendrogram we identified two clusters (cluster 1 = pink, cluster 2 = blue). We have depicted positive relationships among the omics variables in red and negative relationships in blue. For the polygenic scores (PGSs), the ‘_NTm’ suffix denotes non-transmitted maternal PGSs, the ‘_NTf’ suffix denotes the non-transmitted paternal PGSs, and childhood aggression is abbreviated as “aggression”, Attention-Deficit Hyperactivity Disorder as “ADHD”, Major Depressive Disorder as “MDD”, Autism Spectrum Disorder as “Autism”, Educational Attainment as “EA”, and wellbeing spectrum as “wellbeing”. For the metabolites, the ‘amines.’ prefix shows we measured these metabolites on the Liquid Chromatography Mass Spectrometry (LC-MS) amines platform, the ‘steroids.’ prefix shows we measured these metabolites on the LC-MS steroids platform, and the ‘OA.’ prefix shows we measured these metabolites on the Gas Chromatography (GC-) MS organic acids platform. **(a)** Relationships of the 1,614 CpGs and 90 metabolites included in the 3-component DNA methylation-metabolomics PLS model, where the selected CpGs are represented in the columns and the metabolomics traits in the rows. We included the cluster assignments and the full data matrix in **Data S4-S5. (b)** Relationships of the 36 PGSs and 90 metabolites included in the 2-component PGSs-metabolomics PLS model, where the selected PGSs are represented in the columns and the metabolomics traits in the rows. We included the cluster assignments and the full data matrix in **Table S9** and **Data S6**, respectively. **(c)** Relationships of the 36 PGSs and 1,614 CpGs included in the 2-component PGSs-DNA methylation PLS model, where the selected PGSs are represented in the columns and the CpGs in the rows. We included the cluster assignments and the full data matrix in **Data S7-S8**.

#### PGSs-metabolomics

Hierarchical clustering of the pairwise PGSs-metabolomics model identified two clusters of PGSs and two clusters of metabolites (**Fig. 2b; Table S9**). The PGS cluster 1 contains 23 of the PGSs (63.9%) selected by the sPLS-DA model for childhood aggression. This cluster comprises 8 transmitted PGSs, 9 non-transmitted maternal PGSs, and 6 non-transmitted paternal PGSs. The 13 PGSs (36.1%) included in cluster 2 comprised 3 transmitted PGSs, 6 non-transmitted maternal PGSs, and 4 non-transmitted paternal PGSs. The metabolite cluster 1 contains 46 metabolites (51.1%), all of which were amines (76.7% of all amines). The remaining 14 amines, as well as all steroids and organic acids, are included in metabolite cluster 2 (*N* = 44, 48.9%). The average correlation between the PGSs and metabolomics block, i.e., the correlation among all PLS variates of all components simultaneously, was 0.28 (*q* = 9.57×10^−24^). We observed the highest positive correlations between the non-transmitted paternal ADHD (ADHD_NTf) and smoking initiation PGSs (smokinginitiation_NTf) with both essential and non-essential amino acids (ADHD_NTf *r* M = 0.21, *r* range: 0.21-0.22, smokinginitiation_NTf *r* M = 0.22, *r* range: 0.20-0.26; **Fig. 2b; Data S6; Table S9**). Similarly, the most negative correlations were observed between amino acids and the non-transmitted paternal PGSs for intelligence (intelligence_NTf) and self-reported health (selfreportedhealth_NTf, intelligence_NTf *r* M = -0.21, *r* range: -0.22 to -0.21, selfreportedhealth_NTf *r* M = -0.21, *r* range: -0.22 to -0.20). This shows that characteristics tagged by ADHD, smoking initiation, intelligence, and self-reported health correlated to paternal construction of environments influencing offspring urinary amino acid levels.

#### PGSs-DNA methylation

For the pairwise PGSs-DNA methylation model, we again applied hierarchical clustering to find two clusters for the PGSs and DNA methylation data (**Fig. 2c; Data S7**). PGS cluster 1 contains 22 of the PGSs (61.1%) selected by the sPLS-DA model for childhood aggression. This cluster contained 6 transmitted PGSs, 9 non-transmitted maternal PGSs, and 7 non-transmitted paternal PGSs. The 14 PGSs (38.9%) included in cluster 2 comprised 5 transmitted PGSs, 6 non-transmitted maternal PGSs, and 3 non-transmitted paternal PGSs. The DNA methylation cluster 1 contained 1,142 (70.8%) of the CpGs selected by the sPLS-DA models for childhood aggression, and showed the strongest trait enrichments for household socioeconomic status in childhood (OR = 12.58, *p* = 3.68×10^−14^; **Table S10**). The 472 (29.2%) CpGs included in cluster 2 showed the strongest trait enrichment for glucocorticoid exposure (OR = 50.00, *p* ≤ 1.00×10^−^308; **Table S10**). The average correlation between the PGSs and DNA methylation block, i.e., the correlation among all PLS variates of all components simultaneously, was 0.28 (*q* = 9.57×10^−24^). The most negative correlations were observed between the non-transmitted paternal self-reported health PGS with CpGs of cluster 2 (*r* M = -0.18, *r* range: - 0.19 to -0.15), and the highest positive correlations between the cluster 2 CpGs and the non-transmitted paternal ASD PGS (*r* M = 0.151, *r* range: 0.150-0.154; **Fig. 2c**; **Data S8**). This indicates that characteristics tagged by ASD and self-reported health correlated to paternal construction of environments influencing offspring buccal DNA methylation levels.

### Multi-omics model for childhood aggression

We built multi-omics panels for childhood aggressive behavior based on multi-block sPLS-DA (MB-sPLS-DA) models, including PGSs, DNA methylation, and metabolomics data. Here, we report the multi-omics model with an empirical design matrix and results for the null and full design matrices can be found in **Appendices C** and **D**, respectively. After cross-validation, the optimal 5-component model included 14 transmitted and 30 non-transmitted PGSs, 746 CpGs, and all 90 metabolites (**Table S5; Data S9; Fig. S11**). The multi-omics model selected all 30 non-transmitted PGS, and except for the transmitted PGS for childhood IQ, also all the transmitted PGSs (**Data S9**). Out of the 746 CpGs selected by the multi-omics model 204 (27.3%) were also selected by the single-omics DNA methylation model (**Data S2; Data S9**). Trait enrichment analyses for all selected CpGs in the MB-sPLS-DA model showed the strongest enrichment for gender (OR = 3.90, *p* = 3.97×10^−31^), and breast cancer risk (OR = 342.60, *p* = 1.44×10^−21^; **Table S11**). Multi-omics prediction of aggression case-control status in the test data showed an improvement in the prediction (BER = 0.47-0.52; **Table S12; Fig. S12**) as compared to single-omics models including only the PGSs or metabolomics data, but not as compared to the model including only the DNA methylation data (**Table S7**). In contrast, in the clinical cohort, the average classification accuracy was poorer in the multi-omics model (BER = 0.53-0.57; **Table S12**) than for the single-omics models (**Table S7; Fig. S13**).

The average correlations between each omics block in the multi-omics model, i.e., the correlation among all PLS variates of all components simultaneously, were *r* = 0.19 (*q* = 2.13×10^−27^) for PGSs-DNA methylation, *r* = 0.13 (*q* = 1.78×10^−12^) for PGSs-metabolomics, and *r* = 0.15 (*q* = 3.76×10^−16^) for DNA methylation-metabolomics. We observed high absolute correlations (|*r*| ≥ 0.60) between 2 selected PGSs, 14 CpGs, and 13 metabolite, that we summarize in five sets of correlational patterns (**Fig. 3; Data S10; Table S13**). Correlation pattern 1 comprises high negative correlations of citric and fumaric acid with cg12886033 (chr1:65449013), cg14508705 (chr1:172360182), and cg11710553 (chr4:105892960), and high negative correlations of fumaric acid with cg21432062 (chr3:4908643), cg22848658 (chr6:135354586), and cg15841349 (chr12:129348564). The second correlational pattern is characterized by high negative correlations between isocitrate with cg05056638 (chr8:24800824), cg08415582 (chr8:57030523), cg11206167 (chr5:42924367), and cg20704654 (chr20:30072118). Correlation pattern 3 contains the high positive correlation of homocysteine with cg13784456 (chr10:132970405) and cg06144718 (chr10:133048392). The fourth correlational pattern includes high positive correlations between cg03469862 (chr11:68924853) with the transmitted PGS for ADHD, 3-methoxytyrosin, L-isoleucine, L-leucine, L-phenylalanine, L-tryptophan, L-valine, L-glutamine, L-tyrosine, and L-serine. The final correlation pattern comprises a high negative correlation between the non-transmitted by father PGS for EA and cg09674340 (chr1:202509286).

**Fig. 3.**
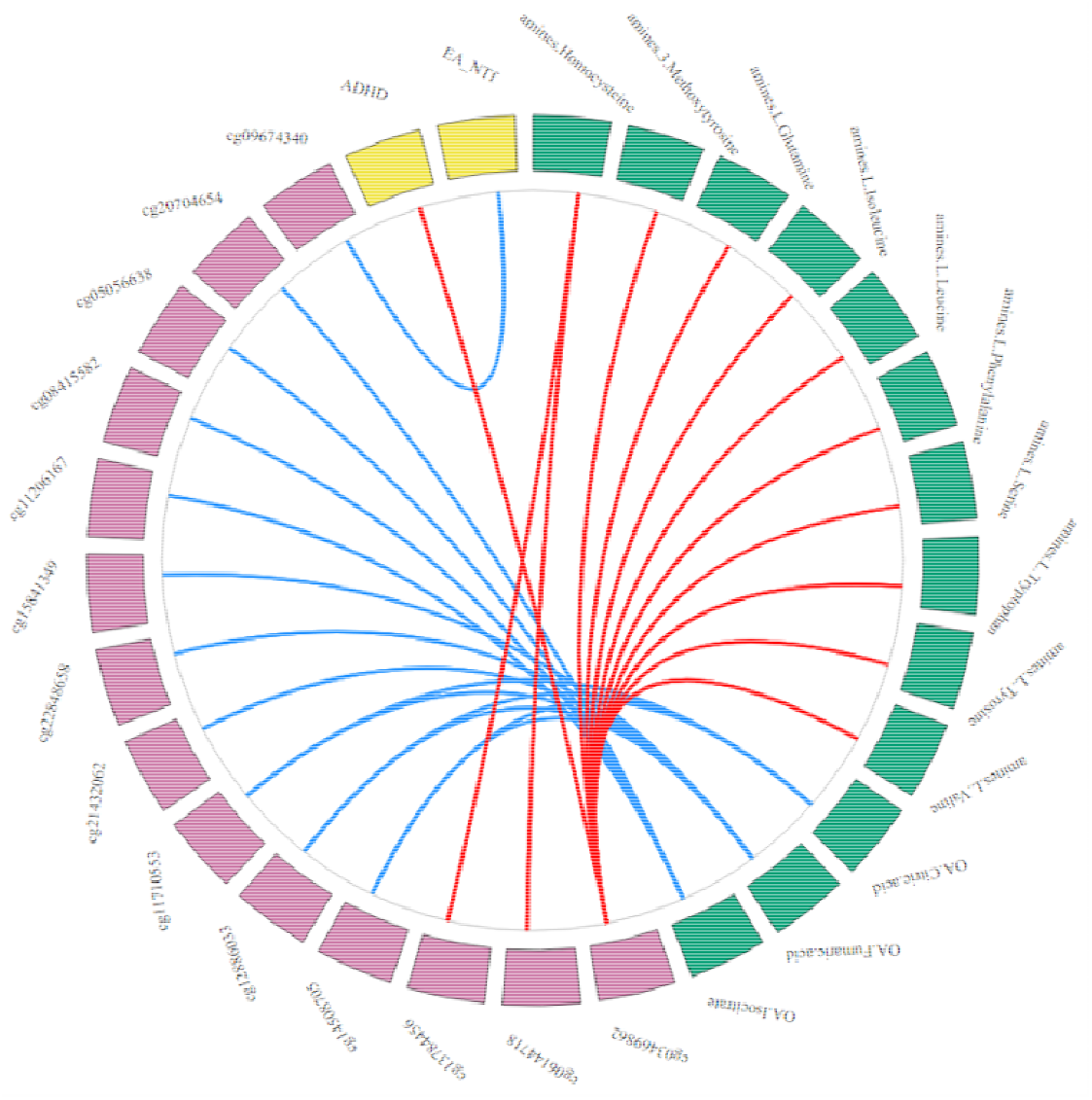
Strong cross-omics connections of the multi-omics traits identified in the 5-component multi-block sparse Partial Least Squares Discriminant Analysis (MB-sPLS-DA) model including the empirical design matrix The outer ring depicts the PGSs, CpGs, and metabolites in yellow, pink, and green, respectively. For the polygenic scores (PGSs), Attention-Deficit Hyperactivity Disorder is abbreviated as “ADHD”, and Educational Attainment as “EA”, and the ‘_NTf’ suffix denotes the non-transmitted paternal PGSs. For the metabolites, the ‘amines.’ prefix shows we measured these metabolites on the Liquid Chromatography Mass Spectrometry (LC-MS) amines platform, and the ‘OA.’ prefix shows we measured these metabolites on the Gas Chromatography (GC-) MS organic acids platform. The inner plot depicts the connetions among the omics variables. Here, we depict only high absolute correlations of the PLS variates (|*r*| ≥ 0.60) between variables of at least two omics blocks, with blue lines reflecting negative correlations and red lines positive correlations. We averaged correlations of the PLS variates across all components in the MB-sPLS-DA model. We included the full data matrix in **Data S10** and the patterns in **Table S15**.

## Discussion

This study comprised an integrative multi-omics analysis of childhood aggressive behavior to identify a multi-omics biomarker panel and to investigate the correlations among the omics blocks included in this panel. In our training data comprising 645 twins (cases = 42.0%, controls = 58.0%) we applied multivariate statistical methods to analyze and integrate transmitted and paternal and maternal non-transmitted PGSs for childhood aggression and for 14 traits genetically correlated with aggression (45 PGSs total), 78,772 CpGs, and 90 metabolites (**Fig. 1**). We build single-omics biomarker panels for each of the omics blocks, that selected 31 PGSs, 1,614 CpGs, and 90 metabolites to discriminate between aggression cases and controls. The markers selected in single-omics models had poor predictive performance in our test (*N* = 277, cases = 42.2%, controls = 57.8%) and clinical datasets (*N* = 142, cases = 45.1%, controls = 54.9%). We explored the pairwise correlations of the omics variables selected by the single-omics models and found that indirect genetic effects for ADHD, ASD, intelligence, smoking initiation, and self-reported health, connected most strongly with buccal DNA methylation and urinary amino acid levels in children and that higher amino acid levels associate with hypomethylation in a cluster of CpG sites.

The multi-omics panel selected 44 PGSs, 746 CpGs, and 90 metabolites, which also had poor predictive performance in our test and clinical datasets. We described five sets of correlational patterns with high absolute correlations (|*r*| ≥ 0.60) of aggression-related omics variables selected by the multi-omics model. Correlation patterns 1 and 3 likely capture the DNA methylation process itself, as homocysteine is involved in the methionine cycle that transfers the methionine methyl group to DNA methyltransferase (Selhub 1999), and tricarboxylic acid (TCA) cycle metabolites, such as citric and fumaric acid, play a role in histone acetylation, and histone and DNA demethylation (Martínez-Reyes and Chandel 2020). The CpGs and their connected metabolites included in patterns 1, 2, and 4 were previously associated with numerous traits, including ageing, inflammatory bowel disease (IBD), prostate cancer, and Myelodysplastic Syndrome (Ooi et al. 2011; Schicho et al. 2012; Dawiskiba et al. 2014; Rist et al. 2017; Lima et al. 2021; Yuan et al. 2021; Xiong et al. 2022). Pattern 5 links a smoking and low birth weight-associated CpG to indirect genetic effects for EA. The negative association of a smoking-associated CpG with indirect genetic effects for EA may indicate that rearing environment influences smoking-related DNA methylation. Pattern 5 may be regarded as being in line with a previous study that reported significant negative correlations of indirect parental EA effect with offspring smoking initiation, number of cigarettes per day, and smoking cessation (Wu et al. 2021), and the well-established phenotypic association of low paternal educational attainment with low offspring birth weight (e.g., Meng and Groth 2018). Overall, the multi-omics correlational patterns associated with a range of traits that link aggression-related omics variables to biological processes related to inflammation, carcinogens, ageing, sex differentiation, intelligence, and smoking.

We explored how sensitive our results are to different specifications of the design matrices and found that these resulted in similar predictive abilities, but selected different numbers of only partially overlapping PGSs and CpGs (**Appendices C** and **D**). In line with expectations, the predictive ability of the null design matrix was slightly better than for the empirical design matrix, reflecting that such a model focuses on selecting discriminatory variables (Singh et al. 2019; Duruflé et al. 2020). We expected that the model with the full design model sacrificed predictive accuracy to select discriminatory variables that are also highly correlated, but this model had the lowest classification error rate as compared to the models with an empirical or null design matrix in the test and clinical data. Multi-omics models with differently specified design matrices differed not only in how many and which PGSs and CpGs were selected, but each multi-omics model provided unique insight into the correlations among the omics variables selected for their association with childhood aggression. In the current study, we relied on the cross-validation results to select the number of variables and components in the model. However, to aid in biological interpretation, other choices can be made.

The single- and multi-omics models selected nearly all the PGSs, with only the transmitted PGSs for ASD, intelligence, childhood IQ, and self-reported health, and the non-transmitted paternal PGSs for age at first birth, EA, insomnia, and loneliness not selected to the single-omics model, and the transmitted PGS for childhood IQ not selected to the multi-omics model (empirical design matrix). The high selection of transmitted PGSs to the biomarker panels likely reflect the genetic correlations of these traits with childhood aggression on which basis we included them in the current study (Ip et al. 2021). The parental non-transmitted PGSs in the genomics block capture the effect of parental genotypes on their offspring’s rearing environment, without confounding by genetic transmission. The non-transmitted PGSs for various traits, including childhood aggression, were consistently retained in the single- and multi-omics models, which is in line with research that shows associations of parenting styles with childhood aggression (Masud et al. 2019).

The CpGs in the single- or multi-omics models did not overlap with the top differentially methylated CpGs for physical aggression as observed in buccal-cells (Cecil et al. 2018b), and were not significantly associated with aggression in a recent blood-based EWAS meta-analysis (van Dongen et al. 2021). That our models selected no CpGs overlapping with these studies was unsurprising, since it was previously shown that the top CpGs from these studies were not associated with aggressive behavior in the buccal DNA methylation data in the NTR and LUMC-Curium cohorts (van Dongen et al. 2021). Trait enrichment of the CpGs selected by the single- and multi-omics models reported enrichment of known aggression risk factors, such as socioeconomic status (Miller and Tolan 2019; Bellair et al. 2019; Hendriks et al. 2020), childhood malnutrition (Liu 2004; Vaughn et al. 2016), and pre-and perinatal risk factors (Van Adrichem et al. 2020). Moreover, we observed trait enrichment for several syndromes, e.g., Down syndrome, that are characterized by intellectual disability, developmental delay, and sometimes by aggressive behaviors among affected individuals. Several of these syndromes, including Klinefelter and Claes-Jensen syndrome, are linked to genetic abnormalities of the sex chromosomes, and CpGs in the single- and mult-omics models are enriched for steroid hormone-related traits, such as glucocorticoid exposure. Together, these results suggest that the CpGs in the single- and multi-omics models reflect the well-established gender differences in aggressive beavior. In addition to the enrichment of glucocorticoid exposure in CpGs selected by the single-omics model, we observed a high correlation between cortisol and cg05153029 (chr20:19769815, *r* = 0.61), that associates with glucocorticoid exposure in the EWAS atlas (full design matrix MB-sPLS-DA model). Cortisol and other hypothalamus-pituitary-adrenal (HPA)-axis molecules have been implicated in aggressive behavior, suggesting a role of the stress response system in aggressive behavior (Hagenbeek et al. 2016). Epigenetic programming of the HPA-axis is influenced by pre- and perinatal factors, such as maternal behavior as observed in rats (Weaver et al. 2004), and maternal stress and early life adversity (Mulligan et al. 2012; Hompes et al. 2013; Jiang et al. 2019), and lower average global DNA methylation levels were reported in patients with Cushing’s Syndrome (CS) in remission, a model for long-standing excessive glucocorticoid (cortisol) exposure (Glad et al. 2017). Thus, epigenetic mechanisms may mediate the association between cortisol and childhood aggression, similarly as the DNA methylation mediates the association between cortisol stress reactivity and childhood trauma (Wrigglesworth et al. 2019).

The single- and multi-omics models selected all 90 LC- and GC-MS metabolites for inclusion, which might be explained by the low expression differences between aggression cases and controls of the urinary amines and organic acids in this sample (Hagenbeek et al. 2020). Another plausible explanation regards the fact that we generated the metabolomics data on three targeted platforms, chosen because they cover relevant metabolites involved in neurotransmitter, inflammation, and steroid hormone pathways that associate with aggression (Hagenbeek et al. 2016). To date, no metabolomics platform can capture the entire metabolome. In addition, within a platform, technical challenges may cause compounds becoming undetectable or not quantifiable. This was the case for the steroid platform being less successful in quantifying (conjugated) sex hormones. Coverage could also be extended to include other metabolite classes, preferably in a hypothesis-free manner by employing non-targeted metabolomics platforms. The five sets of omics variables with high cross-omics correlations in the multi-omics model contained 13 metabolites. These metabolites comprised 8 amino acids, 3 metabolites involved in the TCA cycle, the dopaminergic trace amine 3-methoxytyrosine, and homocysteine, which is involved in cysteine and methionine metabolism. This is in line with previous studies reporting association of metabolites with aggressive behavior (Gulsun et al. 2016; Hagenbeek et al. 2016, 2020; Chen et al. 2020).

This is the first multi-omics study that includes DNA methylation profiles from buccal and metabolomics in urine. Most of the earlier large-scale omics studies were conducted in blood samples. By obtaining omics measurements in easily accessible peripheral tissues (urine and buccal-cells) we could obtain multi-omics data in a substantially larger (∼10-fold) sample compared to most previous multi-omics studies for psychiatric traits, such as depression and suicide risk (Bhak et al. 2019) or post-traumatic stress disorder (Dean et al. 2019), that relied on small training samples (range: 126-165). In the current study, we corrected the omics data for sex, age, and technical covariates, which can be considered as a minimal set of covariates. Future studies may consider including other confounding factors, such as parental smoking, dietary factors, or body mass index. In evaluating the validity of the PGS and multi-omics models in the clinical cohort, the non-transmitted PGSs could not be assessed, as no parental genotypes were available in the clinical cohort. Additional validation in cohorts with complete omics data that applied the same metabolomics and DNA methylation platforms are of large interest, but currently do not exist. Our design is optimal in nearly all other aspects. The twin and clinical cohorts collected all data, including biomarkers, at the same time, using the same arrays and platforms and with similar protocols, thus decreasing unwanted sources of heterogeneity that are often present in sequential multi-omics designs (Wörheide et al. 2021).

By adding a fourth broad exposome block, capturing known risk factors for childhood aggression, such as neighborhood variables (Miller and Tolan 2019), to the multi-omics model, the influence of environmental influences can be explored. Inclusion of other omics layers, such as the transcriptome, proteome, or a microbiome, may give other insights into the biological mechanisms of complex traits like childhood aggression. It should be noted that with the current method, inclusion of additional omics layers will cause an increase in the computational burden. Therefore, reduction of the computational burden might need to be considered in future studies. Parameter reduction may be achieved by including a smaller number of CpGs, by for example including only the top 1% or 5% most variable probes, or by selecting CpGs based on their known association with aggressive behavior, such as the top CpGs from the EWAS meta-analysis for aggressive behavior (van Dongen et al. 2021). While these suggestions, as well as increasing the sample size of training datasets, will benefit future multi-omics investigations into childhood aggression, one of the first steps in aggression research also concerns increasing the sample size of future genome-wide association studies. To understand the underlying biological mechanisms of complex traits we should not put all our efforts into a single approach. By exploring and optimizing various methodologies and merging information from these different approaches we stand the best chance of identifying robust mechanisms.

## Conclusions

Our work entails one of the first applications of multi-omics approaches to childhood psychopathology. The approach we used was developed for dichotomous traits and classification purposes but also gives insight into the how different omics blocks associate with each other. Classification was poor, whereas the multi-omics associations confirm well known associations between childhood aggression and known risk factors as well as provide novel insight into the correlational structure among omics variables from different omics blocks.

## Data Availability

The standardized protocol for large scale collection of urine and buccal-cell samples in the home situation as developed for the ACTION Biomarker Study in children available at http://www.action-euproject.eu/content/data-protocols. The data of the Netherlands Twin Register (NTR) ACTION Biomarker Study may be accessed, upon approval of the data access committee, through the NTR (https://tweelingenregister.vu.nl/information_for_researchers/working-with-ntr-data).

## Declarations

## Acknowledgments

The Netherlands Twin Register (NTR) warmly thanks all twin families for their participation. LUMC-Curium thanks all patients and their parents for participating, and clinicians for their support. NTR and LUMC-Curium are grateful to all researchers involved in the data collection for the ACTION Biomarker Study.

## Ethical approval

The study was conducted according to the guidelines of the Declaration of Helsinki, and approved by the Central Ethics Committee on Research Involving Human Subjects of the VU University Medical Center, Amsterdam (NTR 25th of May 2007 and ACTION 2013/41 and 2014.252), an Institutional Review Board certified by the U.S. Office of Human Research Protections (IRB number IRB00002991 under Federal-wide Assurance-FWA00017598; IRB/institute codes), and the Medical Ethical Committee of Leiden University Medical Center (B17.031, B17.032 and B17.040).

## Informed consent

Parents provided written informed consent for their children and twin parents provided written informed consent for their own participation.

## Consent for publication

Not applicable.

## Code availability

Analysis code is available upon request from the corresponding author.

## Authors’ contributions

Conceptualization, F.A.H., J.D., M.B., S.D. and D.I.B.; methodology, S.D.; formal analysis, F.A.H., R.P. and J.J.H.; investigation, F.A.H., P.J.R., O.F.C., R.R.J.M.V., M.B. and D.I.B.; resources, A.C.H., C.K., E.A.E. and T.H.; data curation, C.E.M.B.; writing—original draft preparation, F.A.H., J.D., R.P., M.B. and D.I.B..; writing—review and editing, all authors; visualization, F.A.H.; supervision, J.D., M.B. and D.I.B.; funding acquisition, A.C.H., C.K., O.F.C., V.F., T.H., R.R.J.M.V., M.B. and D.I.B. All authors have read and agreed to the published version of the manuscript.

