## Supplementary Material for "Integrative multi-omics analysis of childhood aggressive behavior"

**Electronic Supplementary Material**

### **Supplementary Methods**

#### **Appendix A.** Biological sample collection

The standardized protocol for large-scale collection of urine and buccal-cell samples in the home situation as developed for the ACTION Biomarker Study in children is available at <http://www.action-euproject.eu/sites/default/files/NTR_ACTION_UrineDNACollectionProtocol_.pdf>. We asked parents to collect first-morning urine samples of their children with a ‘uritainer’, before the child had washed, as soap residue or moist cloths could influence the measurements. If the child went to the toilet at night, wet the bed, or was suffering from an infection, the flu or common cold, parents had to postpone urine collection. They transferred urine from the ‘uritainer’ to the urine sample tubes (four provided). Parents filled the tubes with a minimum of 1ml of urine for two of the tubes and 10ml of urine for the other two tubes. The tubes could not exceed the limit of 12ml of urine. Next, parents placed the urine tubes in protective blisters and stored in the home freezer (~−18◦C). Researchers transported the urine to the lab in a mobile freezer unit at −18◦C. The lab stored the urine samples at −80◦C until further processing.

The buccal swab collection protocol as employed by the Netherlands Twin Register (NTR) specifies buccal-cells collection on four occasions during two consecutive days: twice in the morning (before breakfast) and twice in the evening (before dinner) (Willemsen et al. 2010). Prior to swabbing, we instructed participants not to eat, brush their teeth, gargle, or rinse their mouth. During each session, participants rubbed four cotton mouth swabs against the inside of the upper and lower lips and the inside of both cheeks. After swabbing participants placed the cotton swaps, tip-down in the provided 15 mL conical tubes (four swabs in each tube) pre-filled with 0.5 mL STE buffer (100 mM sodium chloride, 10 mM Trishydrochloride (pH 8.0) and 10 mM ethylenediaminetetraacetic acid) with proteinase K (0.1 mg/mL) and sodium dodecyl sulfate (SDS) (0.5%) per swab.

#### **Appendix B.** Omics measurements

##### Genomics data

###### Genotyping and imputation

Genotyping was done on Affymetrix AXIOM or Illumina GSA arrays (Ehli et al. 2017; Beck et al. 2019), and genome-wide SNP data were available for 3,334 participants, including 1,702 parents and siblings of twins (AXIOM = 909, GSA = 2425). For each genotyping platform, samples were removed based on the following criteria: 1) DNA sex did not match; 2) Plink heterozygosity F statistic (< -0.10 | > 0.10); and 3) genotyping call rate (< 0.90). Criteria for excluding Single Nucleotide Polymorphisms (SNP) included: 1) minor allele frequency (MAF; <0.005); 2) Hardy-Weinberg Equilibrium (HWE) p-value (< 1×10-5), 3) call rate (< 0.95), 4) percentage of Mendel errors (> 2); and 5) palindromic AT/GC SNPs (MAF 0.4-0.5; Purcell et al. 2007). For each platform, data were aligned with the GoNL reference set (V4). SNPs with an allele frequency difference of larger than 0.10, or mismatching alleles with the reference panel were removed (Boomsma et al. 2014), and samples were removed if DNA Identity By Descent (IBD) state did not match the expected familial relations (PLINKv1.9). Genotypes were then re-aligned, for each platform separately, to the 1000G Phase 3 version 5 reference panel by the PERL based "1000G Imputation preparation and checking" tool v4.3 (<https://www.well.ox.ac.uk/~wrayner/tools/>). The two platforms were subsequently phased (EAGLE v2.4.1) and imputed (MINIMAC3 v2.0.1) following the Michigan Imputation server protocols (Delaneau et al. 2012; Das et al. 2016; Loh et al. 2016). After imputation, the resulting per platform chromosomal files were merged into a single best guess Plink file (PLINK1.9). Data were available for 3,334 participants: 3,149 NTR participants (1,447 twins, 1,702 parents/siblings of twins), and 185 Curium participants.

###### Computation of principal Components

From the best-guess 1000 genomes imputed data, a complete list of all platform-genotyped SNPs on both AXIOM as well as Illumina GSA were extracted from the NTR Curium sample. This makes a single dataset without large SNP missingness where most genotypic information from both platforms is used. These SNPs were also extracted from the 1000G reference panel genotype data with additional filters for MAF (>0.05), and call rate (>0.98). In the 1000G population alone, the SNPs were then LD pruned (PLINK1.9; --indep 50 5 2), and long-range LD regions were removed (Abdellaoui et al. 2013). Then the NTR Curium and 1000 genomes sets were merged for all SNPs that passed the above QC criteria and were present in all sets. Twenty principal components were subsequently calculated using the 1000G populations and then projected on the NTR data (SMARTPCAv7; Price et al. 2006).

###### Transmitted and non-transmitted alleles

The ACTION Biomarker Study comprised 1,506 twins and siblings of twins with two genotyped parents, for whom allele transmission could be established based on the imputed best guess data. SNPs were removed from the imputed data based on the following criteria: 1) MAF (< 0.01); 2) HWE *p*-value (< 1×10^-5^); 3) call rate (> 0.98); 4) SNPs with duplicate positions; 5) SNPs with 3 or more alleles; and 6) non-ACGT SNPs on the autosomes. All the remaining SNPs were used to create the transmitted alleles dataset. The non-transmitted alleles dataset was created by generating a single transmission-disequilibrium test (TDT) pseudo-control genotype for each child (given the two parents; Plink–tucc option) after defining all children as being cases (Clayton 1999). To determine the maternal and paternal transmission of haplotypes, the transmitted and non-transmitted alleles datasets were phased (SHAPEITv2.r904). The resulting haplotypes were converted into mother and father non-transmitted homozygous haploid genotypes for polygenic score analyses of non-transmitted alleles per parent.

###### Calculation polygenic scores

Polygenic scores (PGS) were calculated for transmitted and non-transmitted alleles based on multiple discovery genome-wide association meta-analyses (GWAMAs), that al omitted NTR from the discovery meta-analysis to avoid overlap between the discovery and target samples, including childhood aggression and 14 traits that showed high genetic correlations with aggression (Ip et al. 2021). These traits comprised Attention-Deficit Hyperactivity Disorder (ADHD), Major Depressive Disorder (MDD), Autism Spectrum Disorder (ASD), insomnia, loneliness, educational attainment (EA), self-reported health, wellbeing spectrum, age at first birth, smoking initiation and quantity, age at smoking initiation, intelligence, and childhood IQ (**Table 1**). In the clinical cohort, we calculated only the transmitted PGSs as no parental genotypes were available to calculate the non-transmitted PGSs. The linkage disequilibrium (LD) weighted betas for all 15 traits, with a causal fraction of 0.50, were estimated by LDpred (Vilhjálmsson et al. 2015). We randomly selected 2500 2^nd^ degree unrelated individuals from the NTR as a reference population to calculate the LD patterns. The resulting LD corrected betas were used to calculate the transmitted and non-transmitted PGSs in PLINK1.9. Thus, for each trait we calculated a transmitted, non-transmitted maternal, and non-transmitted paternal PGS (45 PGSs total). We then standardized all PGSs (mean of zero and standard deviation of one).

##### Epigenomics data

###### DNA methylation quality control

Genome-wide methylation data in buccal DNA samples were measured on the Infinium MethylationEPIC BeadChip kit (Illumina, San Diego, CA, USA (Moran et al. 2016)) by the Human Genotyping Facility (HugeF) of ErasmusMC (the Netherlands; <http://www.glimdna.org/>). The ZymoResearch EZ DNA Methylation kit (Zymo Research Corp, Irvine, CA, USA) was used for bisulfite treatment of 500 Ng of genomic DNA obtained from buccal swabs. The Infinium HD Methylation Assay was performed according to the manufacturer’s specification. As previously described (Van Dongen et al. 2018; Odintsova et al. 2019; van Dongen et al. 2021), quality control (QC) and normalization of the methylation data were performed using a pipeline developed by the Biobank-based Integrative Omics Study (BIOS) consortium (Sinke et al. 2019). In short, only samples that passed all five quality criteria of MethylAid were kept (Van Iterson et al. 2014). This resulted in the removal of 173 low-performing samples (11%). We removed an additional 11 samples because of incorrect sample relationships (as assessed with the R package omicsPrint (Van Iterson et al. 2018)) and sex mismatches (as assessed with DNAmArray and meffil (Min et al. 2018) packages). Functional normalization relied on five control probe principal components. Methylation probes were excluded if they overlapped with a SNP or Insertion/Deletion (INDEL), mapped to multiple locations in the genome, or had a success rate of < 0.95 across all samples. Methylation probes were set to missing in a sample if they had an intensity value of zero, bead count < 3, or detection P-value > 0.01. After QC, 787,711 out of 865,859 sites were kept for analysis for 1,424 samples, and missing methylation β-values were imputed with the *imputePCA()* function from the missMDA R package as implemented in the BIOS pipeline for DNA methylation array analysis (Sinke et al.). After imputation, we removed two duplicate NTR samples, and excluded the sample of one Curium-LUMC participant, as this participant (and co-twin) also was present in the NTR dataset.

###### Predicted cellular proportions

After QC and normalization, we applied the cell-type deconvolution algorithm Hierarchical Epigenetic Dissection of Intra-Sample-Heterogeneity (HepiDISH) with the RPC method (reduced partial correlation) to predict cellular proportions in epithelial tissues (Zheng et al. 2018).

##### Metabolomics data

###### Metabolomics measurement protocol

The Metabolomics Facility of the University of Leiden (Leiden, the Netherlands) assessed metabolites in urine on three platforms: one targeting Amines, Steroids and Organic Acids. They took several measures to decrease the analytical error in the data. By pooling aliquots from all urine samples from all children, they created a QC sample. We randomized subjects across batches to ensure random distribution of cases and controls across batches, whilst ensuring that we included twin pairs on the same plate. Each batch included a calibration line, QC samples (every 10 samples), sample replicates and blanks. To compensate for shifts in the sensitivity of the mass spectrometer across batches, in-house developed algorithms were applied using the pooled QC samples. We evaluated the performance and reproducibility of individual metabolites with the Relative Standard Deviation (RSD) of the Quality Control samples (RSDqc). The acceptance criteria for metabolite reporting were RSDqc <15% and background signal <20%. We report metabolites as ‘relative response ratios’ (target area/area of internal standard) after QC correction.

###### LC-MS amines platform

We measured the amine metabolites by ultra-performance liquid chromatography mass spectrometry (UPLC-MS). Prior to chromatic separation, we added methanol to 5 μL of spiked (with internal standards) urine for protein precipitation and the supernatant centrifuged. After sample evaporation (speedvac), we reconstituted the sample in borate buffer (pH 8.5) with AQC reagent. An Agilent 1290 Infinity II LC system (1290 Multicolumn Thermostat and 1290 High Speed Pump, Agilent Technologies, Waldbronn, Germany) with an Accq-Tag Ultra column (Waters Chromatography B.V., Etten-Leur, The Netherlands) achieved chromatic separation. The UPLC was coupled to electrospray ionization on an AB SCIEX quadrupole-ion trap (QTRAP; AB Sciex, Massachusetts, USA). Analytes were monitored in Multiple Reaction Monitoring (MRM) using nominal mass resolution and detected in the positive ion mode. In total, 66 amines we successfully measured.

###### LC-MS steroid platform

We measured the steroid metabolites with UPLC-MS technology. Sample preparation comprised adding internal standards to 90 μL of urine and filtering the samples with a 0.2μm PTFE membrane. Using an Acquity UPLC CSH C18 column (Waters), with a flow of 0.4 mL/min over a 15 min gradient, chromatographic separation was achieved by UPLC (Agilent 1290, San Jose, CA, USA). We transferred samples to a triple quadrupole mass spectrometer (Agilent 6460, San Jose, CA, USA) with electrospray ionization. By switching positive and negative ion mode, analytes were detected in MRM using nominal mass resolution. In total, 13 steroids we successfully measured.

###### GC-MS organic acids platform

We measured the organic acid metabolites using gas chromatography mass spectrometry (GC-MS). In order to extract the organic acids and remove urea, twice liquid-liquid extraction with ethyl acetate applied to 50 μL of spiked (with internal standards) urine. Then, we performed two-step online derivatization procedures. The first reaction comprised oximation with methoxyamine hydrochloride (MeOX, 15 mg/mL in pyridine), and the second reaction comprised N-Methyl-N-(trimethylsilyl)- trifluoroacetamide (MSTFA) silylation. After derivatization, we injected 1 μL of sample into the GC-MS. With helium as a carrier gas (1,7 mL/min), we performed chromatic separation on a 25m (HP-5MS UI) film thickness 30 x 0.25m ID column. We operated the mass spectrometer (Agilent Technologies, Waldbronn, Germany), using a single quadrupole with electron impact ionization (70 eV) in SCAN mode (mass range 50–500). In total, 21 organic acids we successfully measured.

###### Metabolomics data pre-processing for analysis

We reprocessed the metabolomics data for each platform separately. This comprised imputing metabolite measurements that fell below the limit of detection/quantification with half of the value of this limit, or when this limit was unknown with half of the lowest observed level for this metabolite. And we excluded 13 samples in the NTR, and 2 samples in Curium-LUMC from all three metabolomics platforms because the collected urine was not the first-morning urine (e.g., parent-reported time of urine collection was after 12:00 in the afternoon).

### **Supplementary Results**

#### **Appendix C.** Multi-omics model with a null design matrix for childhood aggression

Here, we report the multi-omics model with a null design matrix, and describe results for the empirical design matrix in the main text and for the full design matrix in **Appendix** **D**. After cross-validation the optimal 5-component model included 14 transmitted and 28 non-transmitted PGSs, 1,831 CpGs, and all 90 metabolites, across all components (**Table S5**; **Data S11**; **Fig. S14**). Except for the transmitted PGs for self-reported health and the non-transmitted paternal PGSs for age at first birth and loneliness, the multi-omics model selected all PGSs. Out of the 1,831 CpGs selected by this multi-omics model, the multi-omics model with the empirical design matrix also selected 306 (16.7%) CpGs and the single-omics DNA methylation model also selected 300 (16.4%) CpGs (**Data S2; Data S9**; **Data S11**). Trait enrichment analysis showed the strongest enrichment for Claes-Jensen syndrome (OR = 13.58, *p* = 3.65x10^-39^), and gender (OR = 2.81, *p* = 4.11x10^-35^) (**Table S14**). Multi-omics prediction of aggression case-control status in the test data showed a small improvement in the prediction (BER = 0.48-0.51), as compared to the multi-omics model with an empirical design matrix (**Table S12**; **Fig. S15**). In the clinical data, this multi-omics model also had better average classification accuracy (BER = 0.50-0.52) compared to the multi-omics model with the empirical design matrix (**Table S12**; **Fig. S16**).

The average correlations between each omics block in this multi-omics model, i.e., the correlation among all PLS variates of all components simultaneously, were *r* = 0.06 (*q* = 1.79x10^-03^) for PGSs-DNA methylation, *r* = -0.01 (*q* = 0.63) for PGSs-metabolomics, and *r* = 0.06 (*q* = 5.01x10^-04^) for metabolomics-DNA methylation. These low correlations reflect the use of a null design matrix in this multi-omics model. We observed high absolute correlations (*r* ≥ 0.60) between 3 selected PGSs, 583 CpGs, and 9 metabolites, that are summarized in 4 sets of correlational patterns (**Fig. S17**; **Data S12; Data S13**).

Correlation pattern 1 comprises positive correlations of citric acid with cg26016712 (chr1:28098981, *r* = 0.61) and cg08006125 (chr14:35755071, *r* = 0.60), and negative correlations of citric (*r* = -0.97) and fumaric acid (*r* = -0.68) with the non-transmitted paternal PGS for ADHD (**Fig. S17**; **Data S12; Data S13**). The metabolites also showed high correlations with CpGs in the multi-omics model with an empirical design matrix (**Fig. 3**; **Data S10; Table S13**). The EWAs atlas only included cg26016712 (**Data S13**), where predominantly hypermethylation of this CpG associates with Down syndrome. As compared to healthy controls, increased levels of citric acid in peripheral blood mononuclear cells and lymphoblasts of children with down syndrome have been reported (Convertini et al. 2016). A metabolomics study found associations of several metabolites of the TCA cycle with down syndrome, but plasma and urinary citric acid levels were not significantly increased in children with down syndrome (Caracausi et al. 2018). Both this and the current study suggest that dysregulation of the TCA pathway is involved in down syndrome, which is corroborated by a recent gene expression meta-analysis that found significant upregulation in down syndrome for genes in the TCA cycle (Pecze and Szabo 2021). In humans, no TCA cycle metabolites have been associated with ADHD to date (Bonvicini et al. 2018; Wang et al. 2021).

The second correlational pattern is characterized by high negative correlations between the non-transmitted maternal PGS for smoking initiation with cysteine (*r* = 0.62) and with 49 CpGs from across the genome (*r* M = -0.61, *r* range: -0.64 to -0.60; **Fig. S17**; **Data S12; Data S13**). Correlation pattern 3 also has high correlation between a smoking-related PGS, a metabolite, and multiple CpGs. Specifically, this pattern includes the high positive correlations between the transmitted PGS for smoking initiation and 2-hydroxybutyric acid (2-HB, *r* = 0.72), of both traits with 200 CpGs (PGS *r* M = 0.81, *r* range: 0.67-0.87, 2-HB *r* M = 0.68, *r* range: 0.60-0.75), of 67 CpGs with only the transmitted PGS for smoking initiation (*r* M = 0.70, *r* range: 0.60-0.81), and of cg23617042 (chr4:174158959) with only 2-hydroxybutyric acid (*r* = 0.64; **Fig. S17**; **Data S12; Data S13**). Furthermore, in correlation pattern 3 we also observed high negative correlations of smoking initiation and 2-hydroxybutyric acid with 28 CpGs (PGS *r* M = -0.79, *r* range: -0.86 to -0.71, 2-HB *r* M = -0.66, *r* range: -0.72 to -0.61), and of 4 CpGs with only the transmitted PGS for smoking initiation (*r* M = -0.68, *r* range: -0.71 to -0.64; **Fig. S17**; **Data S12; Data S13**).

Cysteine and 2-hydroxybutyric acid are closely related metabolites. During synthesis of alpha-ketobutyric acid, 2-hydroxybutyric acid is formed as a byproduct, while alpha-ketobutyric acid itself is a byproduct when cystathionine is cleaved into cysteine for incorporation into glutathione. Urinary levels of 2-hydroxybutyric acid likely reflect glutathione synthesis and thus oxidative stress. The relationship between smoking and oxidative stress is well-established (Caliri et al. 2021), with lower plasma cysteine levels reported in smokers (Moriarty 2003), and increased levels in participants 1 month after smoking cessation (Goettel et al. 2017). Several smoking-related metabolites associate with other hypo- and hyper- DNA methylation (Huang et al. 2018). We observed enrichment of predominantly prenatal and fertility-related traits for both the 49 CpGs associated with the non-transmitted by mother PGS for smoking initiation and the 299 CpGs associated with the transmitted PGS for smoking initiation and 2-hydroxybutyric acid (**Table S15**). While many of the enriched traits relate to maternal health or behavior, these CpGs show no enrichment for (maternal) smoking-related traits, suggesting that the non-transmitted genetic predisposition for smoking initiation in mothers associates with an intrauterine environment that influences non-smoking-related offspring DNA methylation levels. Together, these correlation patterns suggest that, while both direct and maternal indirect genetic effects for smoking initiation influence offspring DNA methylation levels, the direct and indirect genetic effects influence unique CpGs.

The final correlation pattern comprises a high positive correlation between cg03469862 (chr11:68924853) with the amino acids L-glutamine (*r* = 0.63), L-isoleucine (*r* = 0.60), L-phenylalanine (*r* = 0.61), L-serine (*r* = 0.62), and L-valine (*r* 0.62; **Fig. S17**; **Data S12; Data S13**). Correlation pattern 4 of the multi-omics model with the empirical design matrix also included these correlations, where we saw high correlations between cg03469862 with these and other metabolites, as well as with the transmitted PGS for ADHD (**Fig. 3**; **Data S10; Table S13**).

#### **Appendix D.** Multi-omics model with a full design matrix for childhood aggression

Here, we report the multi-omics model with a full design matrix, and describe results for the empirical design matrix in the main text and for the null design matrix in **Appendix** **C**. After cross-validation the optimal 2-component model included 10 transmitted and 26 non-transmitted PGSs, 65 CpGs, and all 90 metabolites, across all components (**Table S5**; **Data S14**; **Fig. S18**). This multi-omics model selected the transmitted PGSs for ADHD, age at first birth, age at smoking initiation, ASD, number of cigarettes per day, insomnia, loneliness, MDD, self-reported health, and wellbeing. The non-transmitted maternal PGSs comprised aggression, ADHD, age at first birth, age at smoking initiation, ASD, number of cigarettes per day, EA, intelligence, loneliness, MD, self-reported health, and wellbeing, and the non-transmitted paternal PGSs comprised aggression, ADHD, age at smoking initiation, childhood IQ, number of cigarettes per day, EA, insomnia, intelligence, loneliness, MDD, self-reported health, and wellbeing. Out of the 65 CpGs selected by this multi-omics model, the multi-omics model with the empirical or null design matrix selected none of the CpGs, and the single-omics DNA methylation model selected 7 (10.8%) of these CpGs (**Data S2; Data S9**; **Data S11**; **Data S14**). Trait enrichment analysis showed the strongest enrichment for glucocorticoid exposure (OR = 50.61, *p* = 6.63x10^-26^), primary Sjögren’s syndrome (OR = 8.68, *p* = 1.48x10^-03^), and maternal phthalate exposure (OR = 36.53, *p* = 1.52x10^-03^; **Table S16**). Multi-omics prediction of aggression case-control status in the test data showed a small improvement in the prediction (BER = 0.46-0.47), as compared to the multi-omics models with an empirical or null design matrices (**Table S13**; **Fig. S19**). Similarly, the average classification accuracy (BER = 0.48-0.50) in the clinical data was better in this multi-omics model as compared to the multi-omics models with an empirical or null design matrices (**Table S13**; **Fig. S20**).

The average correlations between each omics block in the multi-omics model, i.e., the correlation among all PLS variates of all components simultaneously, were *r* = 0.17 (*q* = 2.22x10^-09^) for PGSs-DNA methylation, *r* = 0.12 (*q* = 1.13x10^-05^) for PGSs-metabolomics, and *r* = 0.15 (*q* = 1.15x10^-07^) for DNA methylation-metabolomics. These correlations are very similar to those observed in the multi-omics model with the empirical design matrix, suggesting that in using an empirical design matrix as opposed to the full design matrix we compromised little in selecting highly correlated and discriminatory variables across all omics blocks. We observed high absolute correlations (*r* ≥ 0.60) between 1 selected PGS, 3 CpGs, and 22 metabolites, that can we summarized in 2 sets of correlational patterns (**Fig. S21**; **Data S15; Table S17**).

The first correlation pattern consists positive correlations between cg05410331 (chr9:115085154) and the non-transmitted by father PGS for ADHD (*r* = 0.91), and both traits with 3-methoxytyramine (*r* CpG = 0.66, *r* PGS = 0.73), 3-methoxytyrosine (*r* CpG = 0.67, *r* PGS = 0.74), 5-hydroxy-L-tryptophan (*r* CpG = 0.68, *r* PGS = 0.74), asymmetric dimethylarginine (ADMA, *r* CpG = 0.71, *r* PGS = 0.77), ethanolamine (*r* CpG = 0.69, *r* PGS = 0.76), O-acetyl-L-serine (*r* CpG = 0.64, *r* PGS = 0.70), symmetric dimethylarginine (SDMA, *r* CpG = 0.67, *r* PGS = 0.74), serotonin (*r* CpG = 0.69, *r* PGS = 0.75), and various amino acids (CPGs *r* mean = 0.69, *r* range: 0.66-0.72, PGS *r* mean = 0.76, *r* range: 0.73-0.79; **Fig. S21**; **Data S15; Table S17**). This correlation patterns also includes the positive correlations of the non-transmitted by father PGS for ADHD with cysteine (*r* = 0.60), gamma-aminobutyric acid (*r* = 0.62), glutathione (*r* = 0.65), methionine-sulfone (*r* = 0.65), norepinephrine (*r* = 0.63), L-tryptophan (*r* = 0.65), and L-tyrosine (*r* = 0.64; **Fig. S21**; **Data S15; Table S17**). In the pairwise metabolomics-PGS model, we also observed high positive correlations of many of the amino acid included in correlation pattern 1, with the non-transmitted by father PGS for ADHD (*r* M = 0.21, *r* range: 0.21-0.22; **Fig. 2b**; **Data S6**; **Table S9**). Higher cord plasma branched-chain amino acids (BCAAs; i.e., L-leucine, L-isoleucine, and L-valine) levels associate with an increased risk of childhood ADHD (Anand et al. 2021). In addition, using Mendelian Randomization 20 metabolites, including phenylalanine and tyrosine, were shown to have causal effects on ADHD (Yang et al. 2020). Cg05410331 is located in the gene body of the MicroRNA 3134 (MIR3134) and Polypyrimidine Tract Binding Protein 3 (PTBP3) genes. The EWAS atlas reports associations of CpGs in the PTBP3 gene with traits such as smoking, mortality, and aging (**Table S17**). Our observed positive correlation between the non-transmitted by father PGS for ADHD and levels of urinary amino acids and neurotransmitter-related metabolites suggests characteristics tagged by ADHD correlate to paternal construction of environments influencing these offspring urinary metabolites and DNA methylation levels for cg05410331.

Positive correlations between cortisol and cg21444670 (chr17:57048710, *r* = 0.60) and cg05153029 (chr20:19769815, *r* = 0.61) characterize the second correlational pattern (**Fig. S21**; **Data S15; Table S17**). The EWAS atlas only includes cg05153029 (**Table S17**), where hypomethylation of this CpG associates with glucocorticoid exposure. This finding is in line with a study that reported lower average global DNA methylation in patients with Cushing’s Syndrome (CS) in remission as compared to controls, as CS is a model for long-standing excessive cortisol exposure (Glad et al. 2017). The observed positive correlation between urinary cortisol levels and cg05153029 methylation levels (i.e., higher urinary cortisol levels associated with higher DNA methylation levels) appears to be incongruous with the hypothesis that exposure to high levels of cortisol results in hypomethylation. Cg21444670 is located in the body of the Protein Phosphatase, Mg2+/Mn2+ Dependent 1E (*PPM1E*) gene, the EWAS atlas reported associations of CpGs in this gene with traits such aging, asthma, oral squamous cell carcinoma (OSCC), and down syndrome (**Table S17**). The positive correlation between cortisol levels and ageing-related CpGs is consistent with the observation that total daily cortisol production associates with accelerated DNA methylation age in adolescent girls (Davis et al. 2017). The association with asthma-related CpGs is likely because synthetic glucocorticoids are widely used to treat asthma and influence the anti-inflammatory and immunosuppressive properties of cortisol (Scherholz et al. 2019). In line with the positive correlation between cortisol levels and OSCC-related CpGs, elevated levels of morning plasma and salivary cortisol were reported in patients with OSCC (Sharma et al. 2018). In contrast, lower levels of cortisol were found in young adult males with down syndrome (Bricout et al. 2008), though more recent studies found no differences between adolescent with down syndrome and adolescent with typical development (Pitchford et al. 2019; Gutierrez-Hervas et al. 2020).

### **Supplementary Figures**

#### **Fig. S1.** Scree plots of the Principal Components Analysis (PCA) of the three omics blocks.

**(a)** Scree plot of the metabolomics data. **(b)** Scree plot of the DNA methylation data. Here only the 644 Principal Components (PCs) with eigenvalues equal to or larger than one are depicted. **(c)** Scree plot of the transmitted and non-transmitted polygenic scores (PGSs).

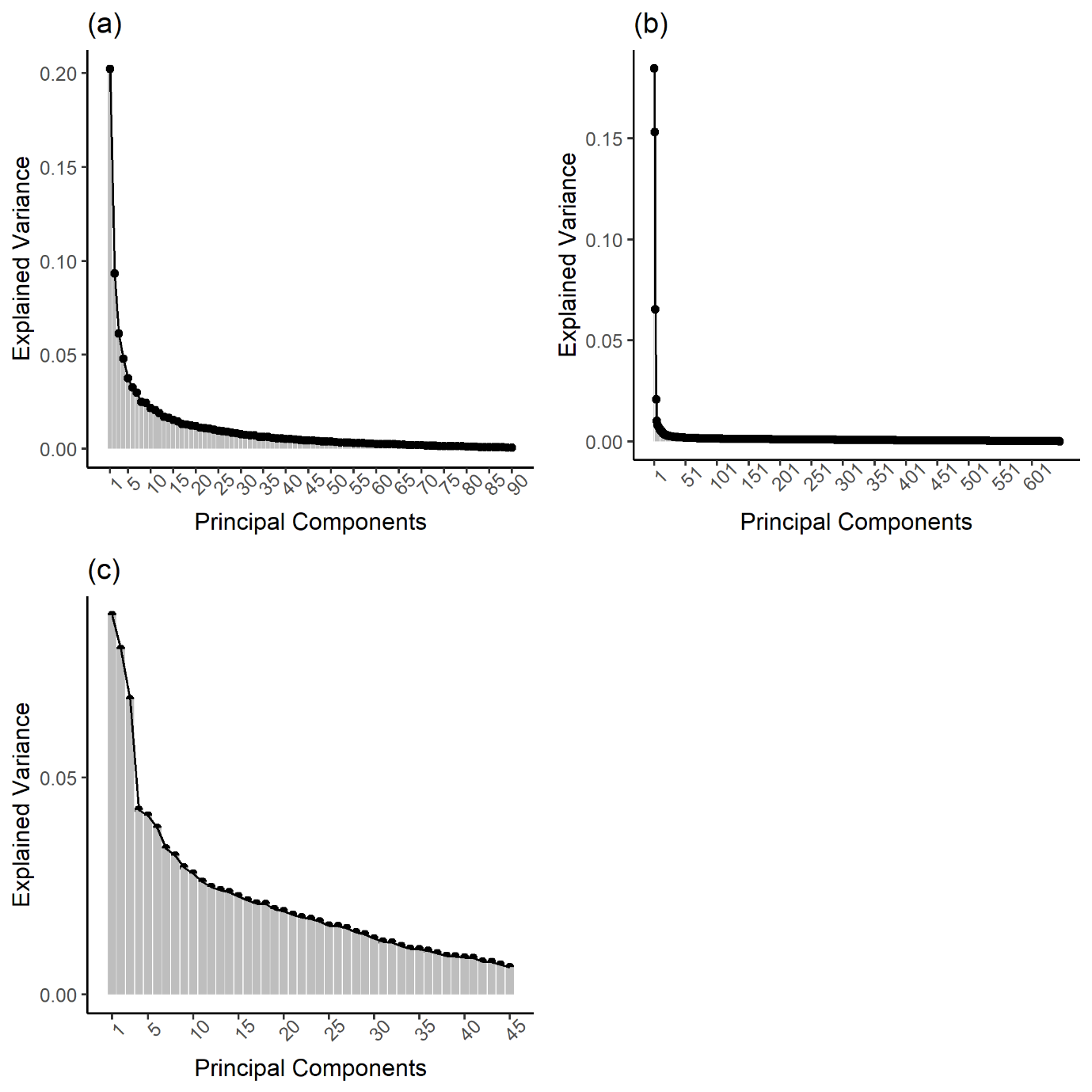

#### **Fig. S2.** Balanced error rates (BER) for the number of components to retain in the Partial Least Squares Discriminant Analyses (PLS-DA) to predict aggression cases and controls in the training data.

**(a)** BER for the 8-component PLS-DA model of the metabolomics data. **(b)** BER for the 6-component PLS-DA model of the DNA methylation data. **(c)** BER for the 4-component PLS-DA model of the transmitted and non-transmitted polygenic scores (PGSs). The BER for the centroids distance is depicted in light grey, for the Mahalanobis distance in dark grey, and of the maximum distance in black.

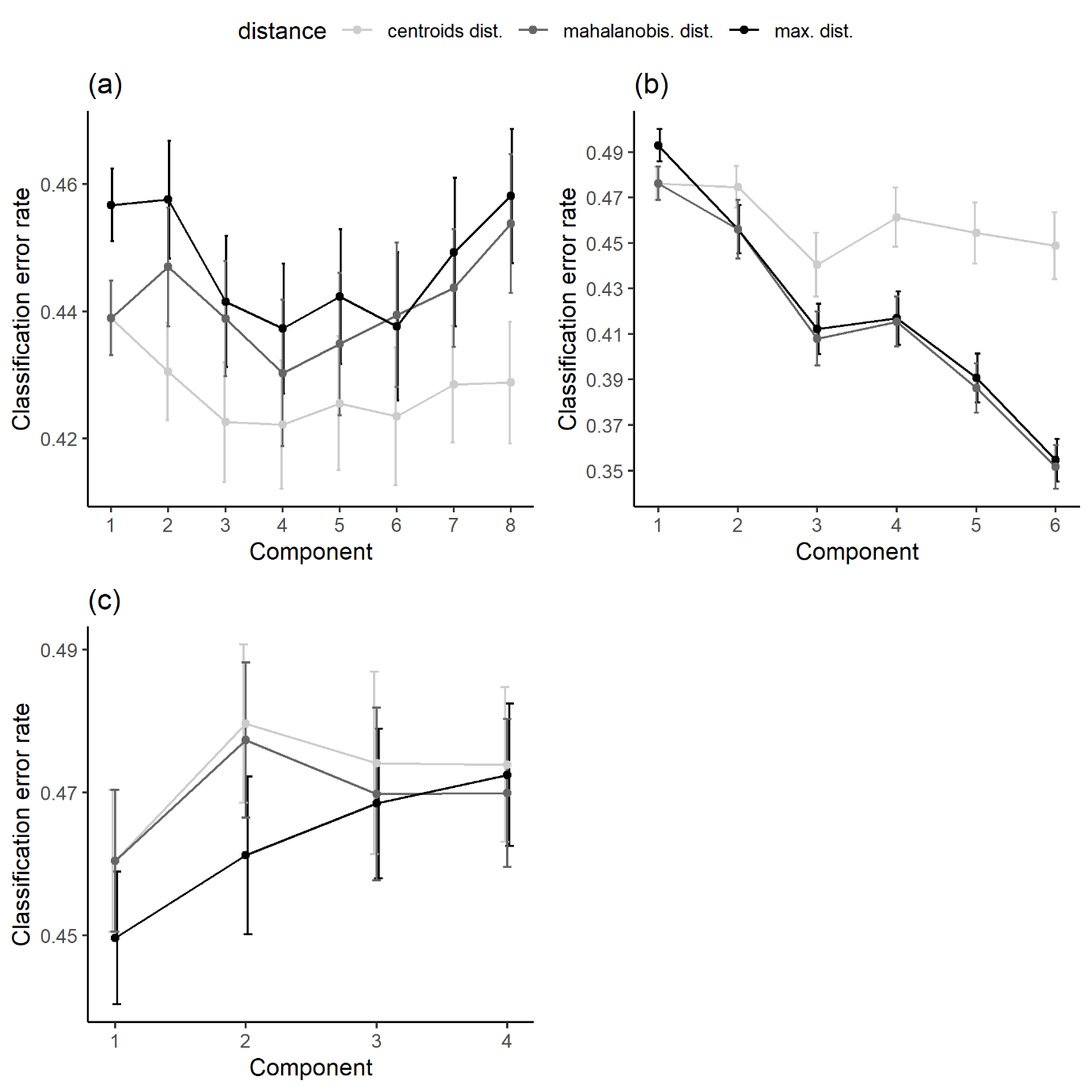

#### **Fig. S3.** Balanced error rates (BER) for the number of omics variables per component to retain in the sparse Partial Least Squares Discriminant Analyses (sPLS-DA) models to predict aggression cases and controls in the training data.

**(a)** BER, with centroids prediction distance, of the 3-component sPLS-DA model for the metabolomics data. **(b)** BER, with Mahalanobis prediction distance, of the 6-component sPLS-DA model for the DNA methylation data. **(c)** BER, with Mahalanobis prediction distance, of the 2-component sPLS-DA model for the transmitted and non-transmitted polygenic scores (PGSs). The BERs for each component are given in shades of gray, ranging from the lightest gray for one component to black for 6 components. The diamonds represent the selected number of features per component by the sPLS-DA model.

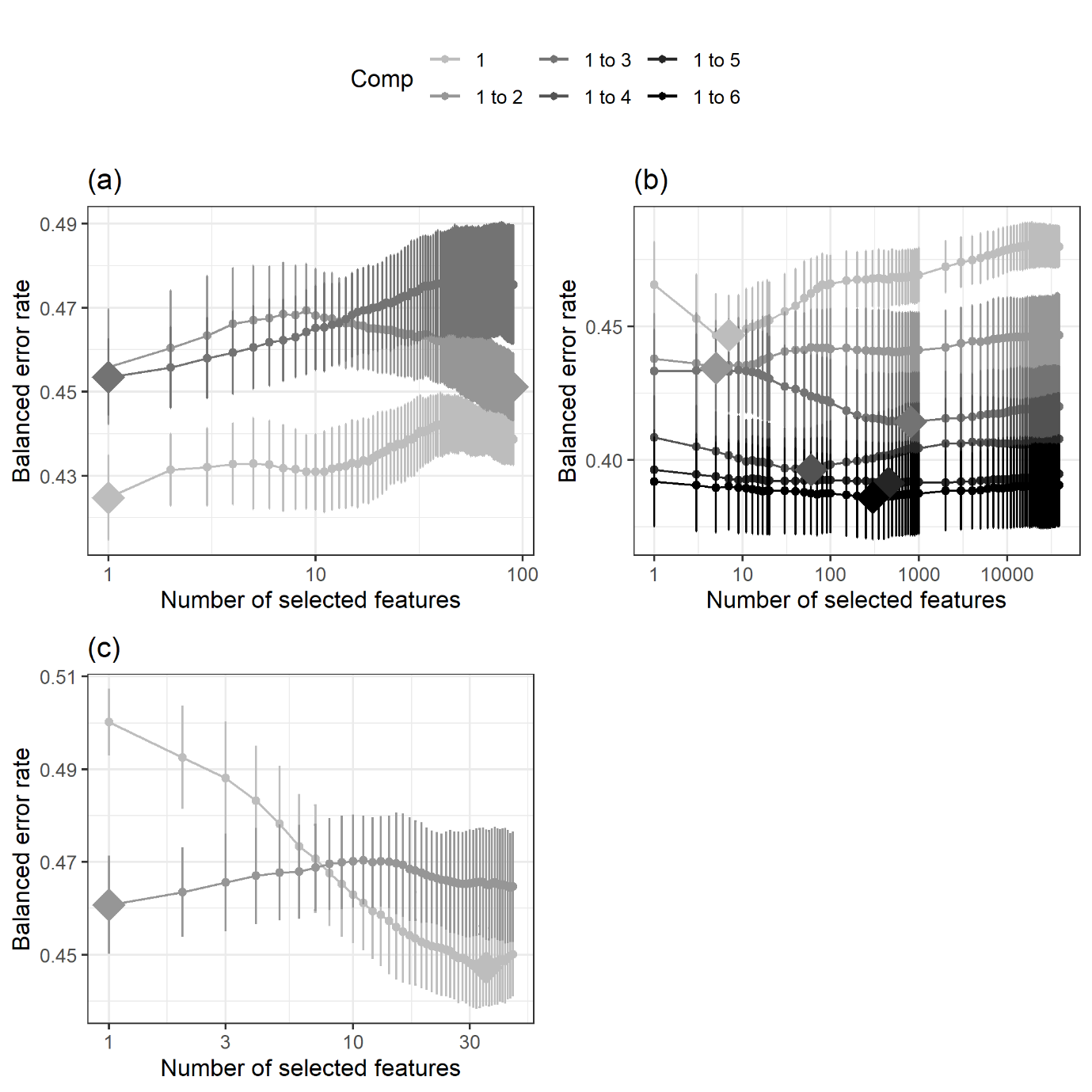

#### **Fig. S4.** Balanced error rates (BER) for the number of components to retain in the final sparse Partial Least Squares Discriminant Analyses (sPLS-DA) to predict aggression cases and controls in the training data.

**(a)** BER, with centroids prediction distance, of the 2-component final sPLS-DA model for the metabolomics data. **(b)** BER, with Mahalanobis prediction distance, of the 6-component final sPLS-DA model for the DNA methylation data. **(c)** BER, with Mahalanobis prediction distance, of the 2-component final sPLS-DA model for the transmitted and non-transmitted polygenic scores (PGSs).

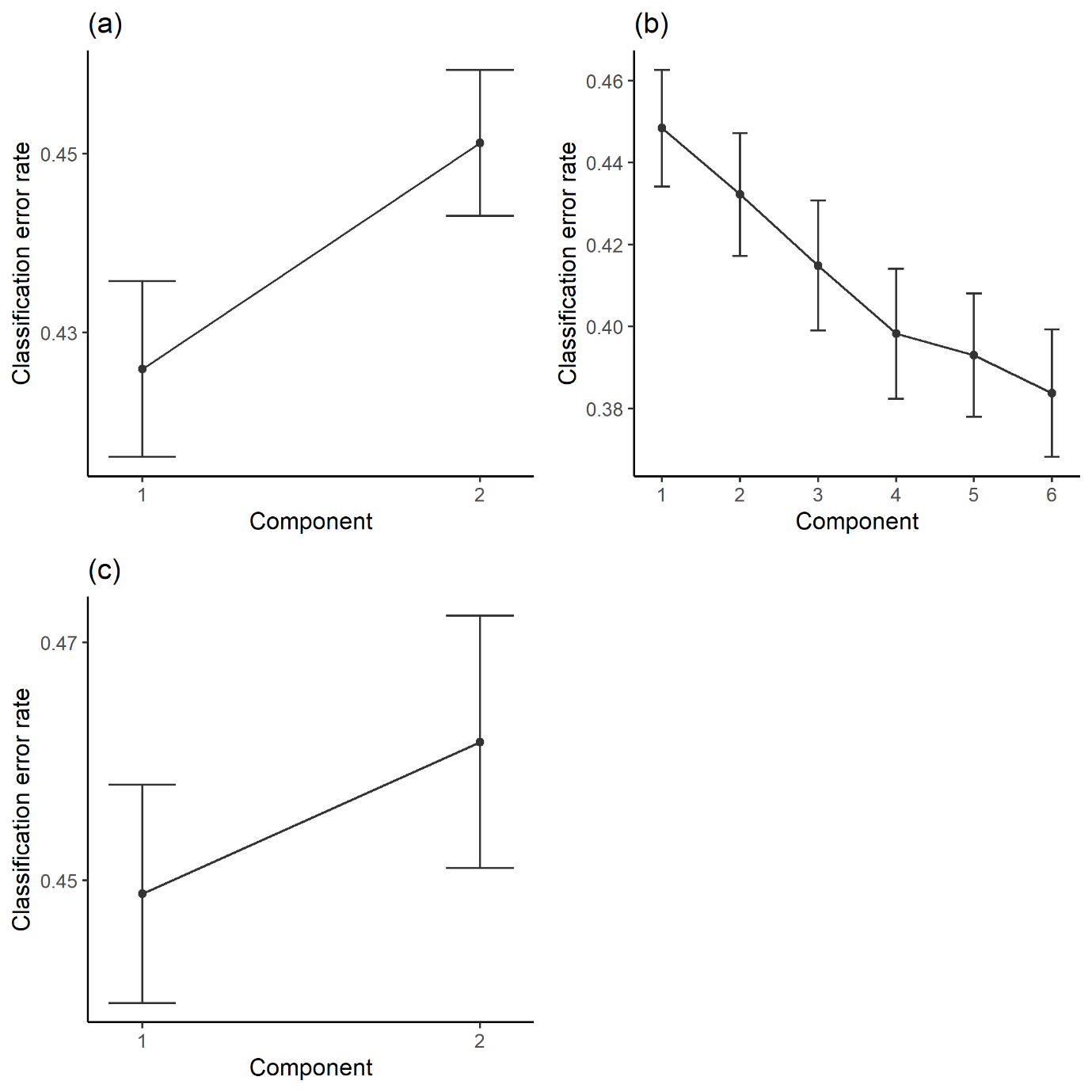

#### **Fig. S5.** Balanced error rates (BER) for the number of components to retain in the multi-block sparse Partial Least Squares Discriminant Analyses (MB-sPLS-DA) models to predict aggression cases and controls in the training data.

**(a)** BER for the 6-component MB-sPLS-DA model with empirical design matrix. **(b)** BER for the 6-component MB-sPLS-DA model with null design matrix. **(c)** BER for the 6-component MB-sPLS-DA model with full design matrix. The BER for the centroids distance is depicted in light grey, for the Mahalanobis distance in dark grey, and of the maximum distance in black.

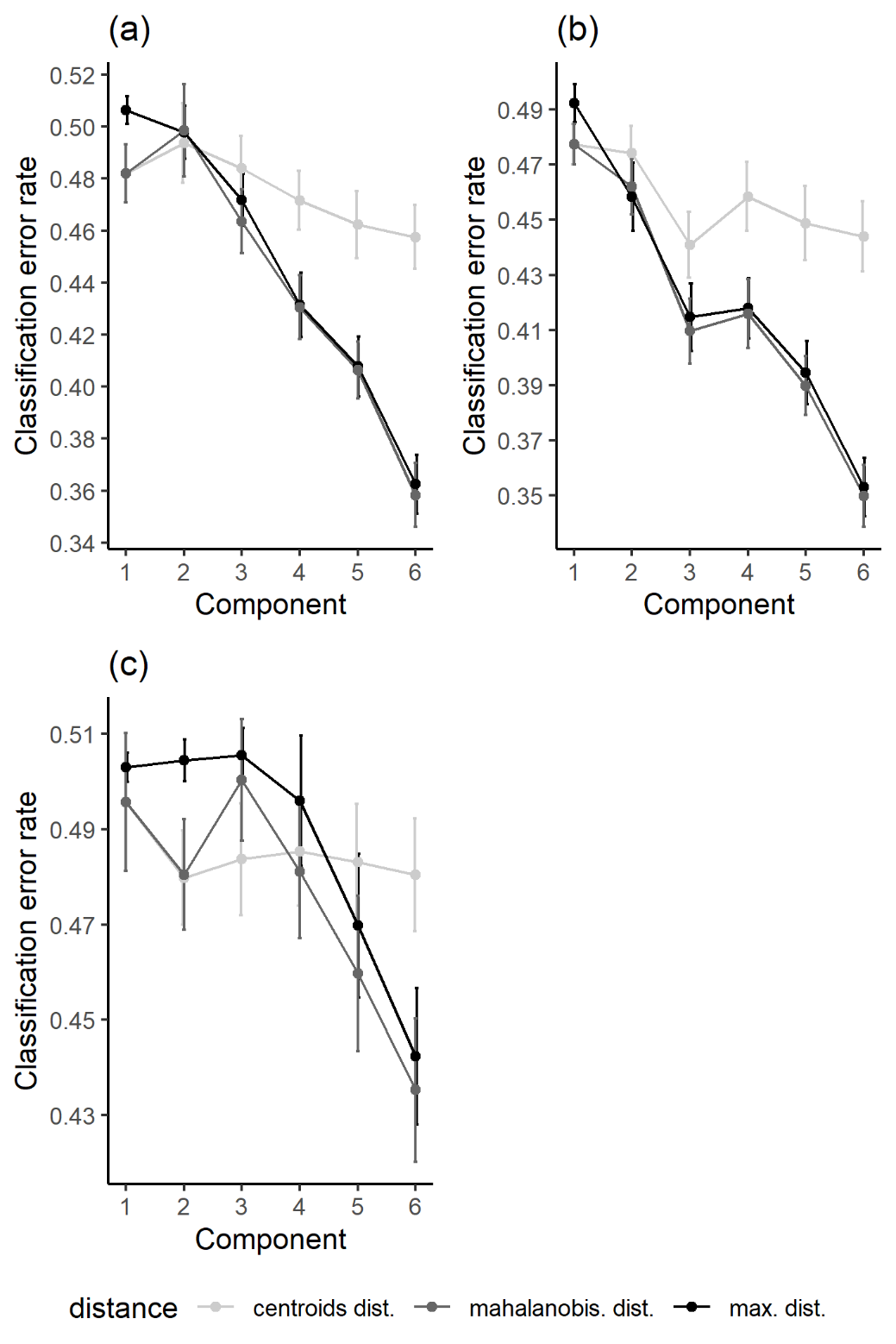

#### **Fig.** **S6.** Balanced error rates (BER) for the number of variables to retain per omics block and per component in the multi-block sparse Partial Least Squares Discriminant Analysis (MB-sPLS-DA) models to predict aggression cases and controls in the training data.

In the model tuning 80 possible sub-models were evaluated, each with a different combination of variables to select per omics block. **(a)** BER, with Mahalanobis prediction distance, of the 6-component MB-sPLS-DA model with empirical design matrix. **(b)** BER, with Mahalanobis prediction distance, of the 6-component MB-sPLS-DA model with null design matrix. **(c)** BER, with Mahalanobis prediction distance, of the 6-component MB-sPLS-DA model with full design matrix.

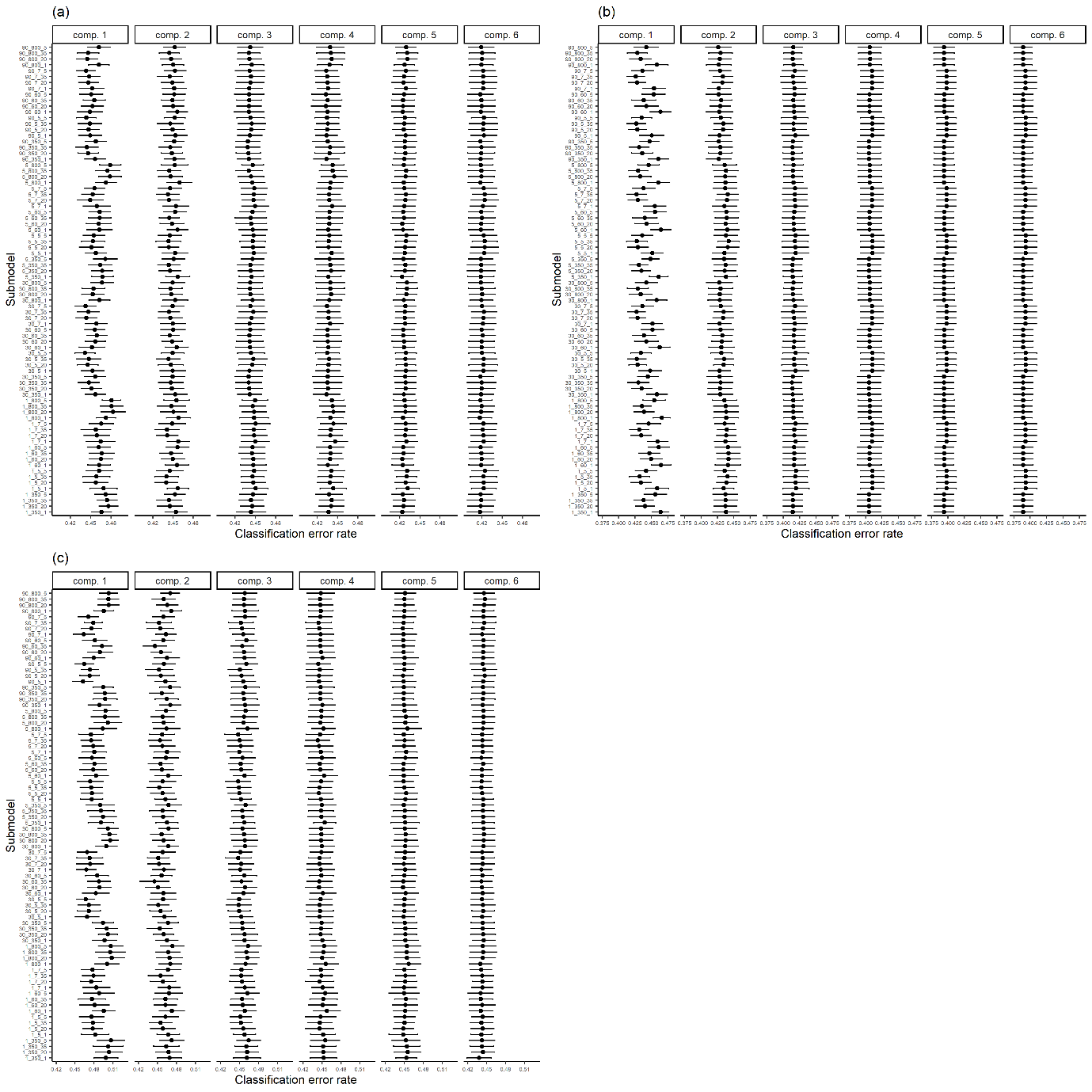

#### **Fig.** **S7.** Balanced error rates (BER) for the number of components to retain in the final multi-block Partial Least Squares Discriminant Analyses (MB-PLS-DA) to predict aggression cases and controls in the training data.

**(a)** BER, with Mahalanobis prediction distance, of the 5-component final MB-sPLS-DA model with empirical design matrix. **(b)** BER, with Mahalanobis prediction distance, of the 5-component final MB-sPLS-DA model with null design matrix. **(c)** BER, with Mahalanobis prediction distance, of the 2-component final MB-sPLS-DA model with full design matrix.

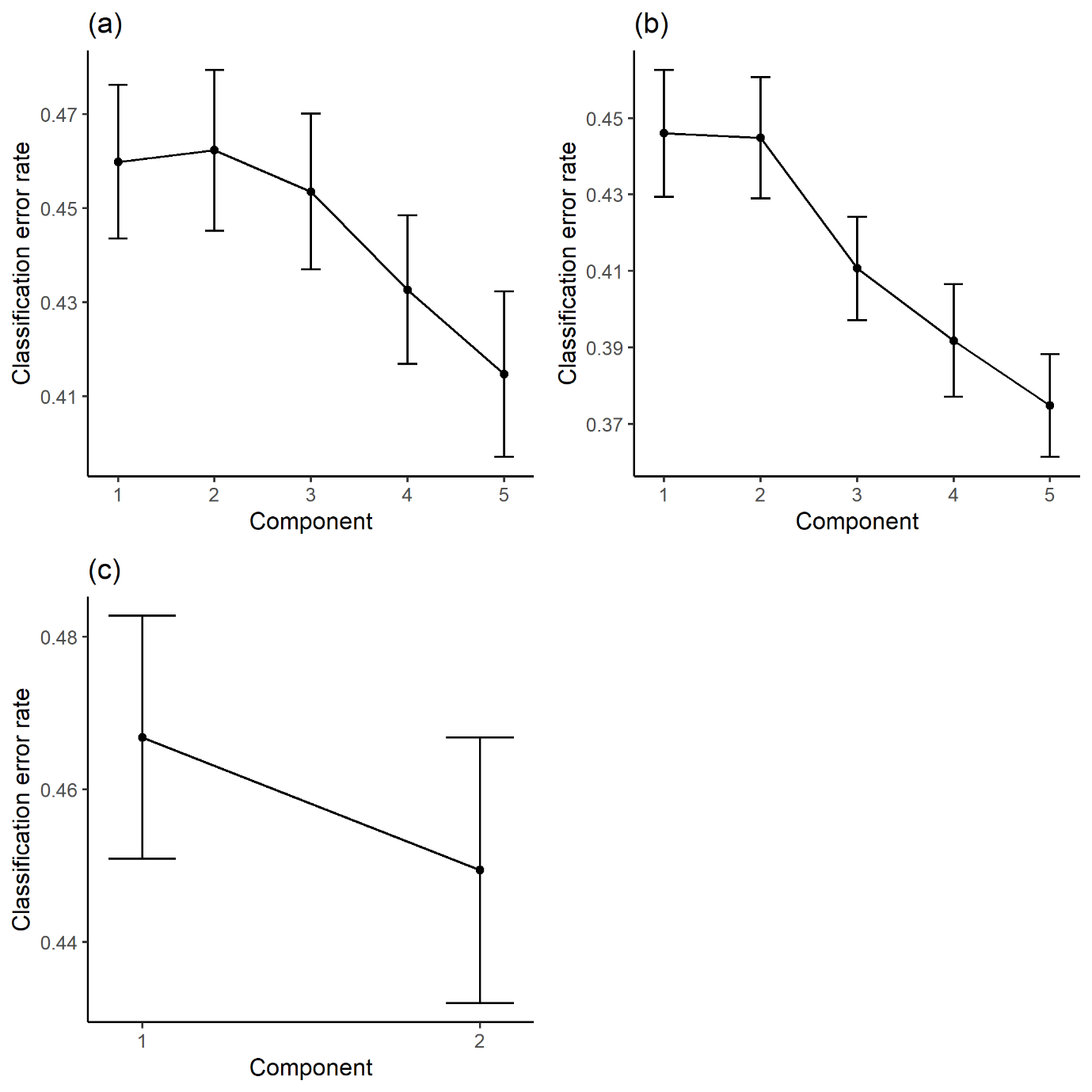

#### **Fig. S8.** Loadings of the sparse Partial Least Squares Discriminant Analysis (sPLS-DA) of the aggression cases and controls in the training data, with 95% confidence ellipses.

The loadings of the aggression controls have been depicted in grey, and the loading scores of the cases have been depicted in black. The first subfigure contains the loadings for the metabolomics data, subfigures 2-16 contains the loadings for the DNA methylation data (depicted as ‘epigenomics’ in figure), and subfigure 17 contains the loadings for the transmitted and non-transmitted polygenic scores (PGSs; depicted as ‘genomics’ in figure).

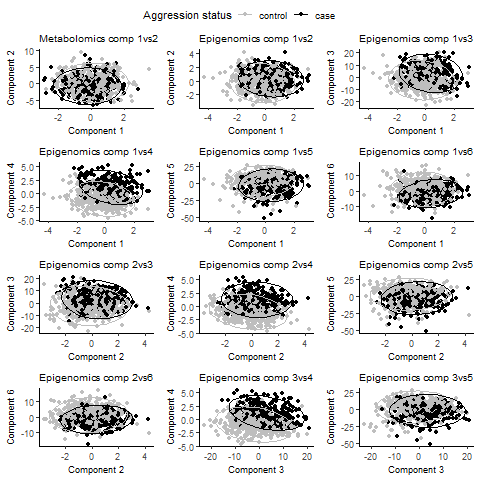

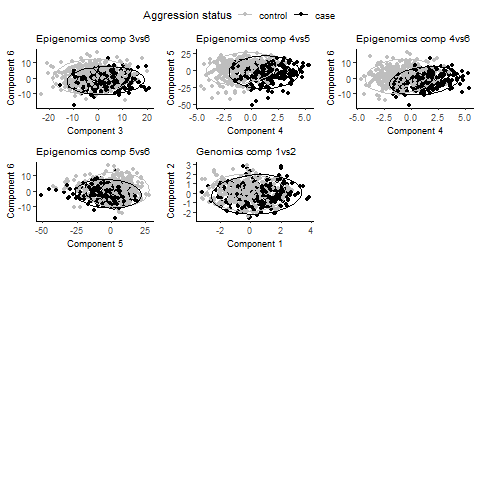

#### **Fig. S9.** Loadings of the sparse Partial Least Squares Discriminant Analysis (sPLS-DA) of the aggression cases and controls as predicted in the test data, with 95% confidence ellipses from the training data.

The loadings of the aggression controls have been depicted in grey, and the loading scores of the cases have been depicted in black. The first subfigure contains the loadings for the metabolomics data, subfigures 2-16 contains the loadings for the DNA methylation data (depicted as ‘epigenomics’ in figure), and subfigure 17 contains the loadings for the transmitted and non-transmitted polygenic scores (PGSs; depicted as ‘genomics’ in figure).

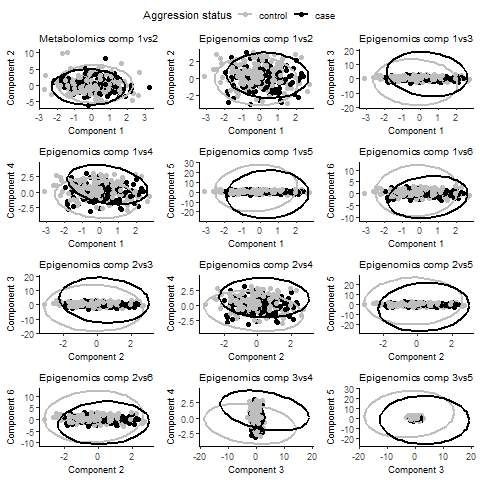

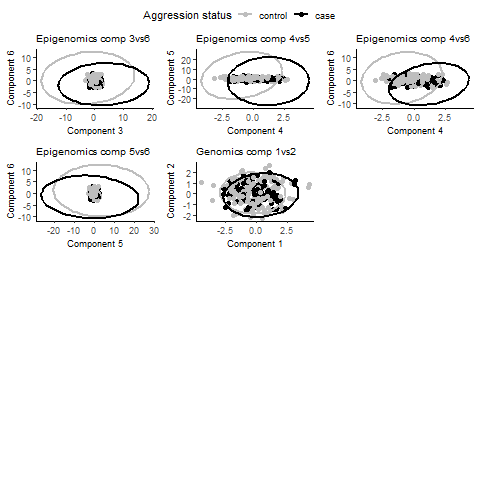

#### **Fig. S10.** Loadings of the sparse Partial Least Squares Discriminant Analysis (sPLS-DA) of the aggression cases and controls as predicted in the clinical data, with 95% confidence ellipses from the training data.

The loadings of the aggression controls have been depicted in grey, and the loading scores of the cases have been depicted in black. The first subfigure contains the loadings for the metabolomics data, subfigures 2-16 contains the loadings for the DNA methylation data (depicted as ‘epigenomics’ in figure), and subfigure 17 contains the loadings for the transmitted and non-transmitted polygenic scores (PGSs; depicted as ‘genomics’ in figure).

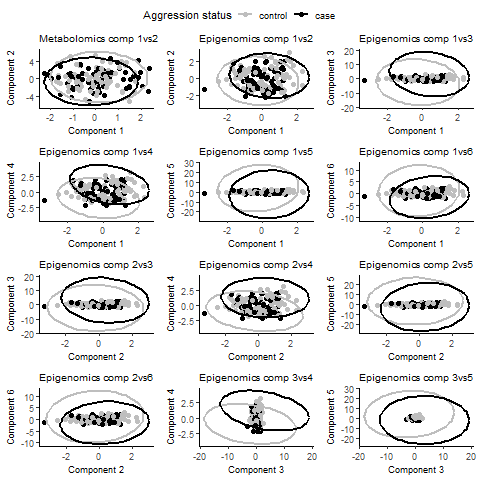

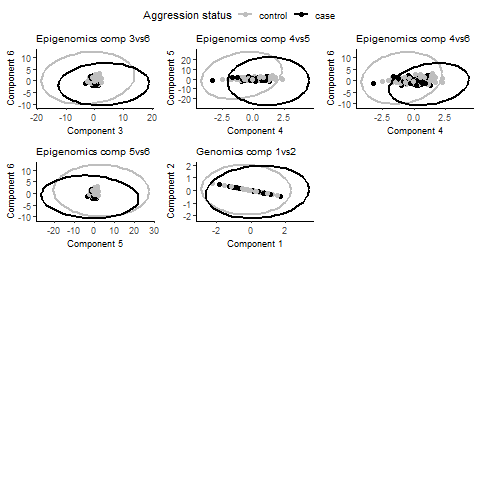

#### **Fig. S11.** Loadings of the 5-component multi-block sparse Partial Least Squares Discriminant Analysis (MB-sPLS-DA), with the empirical design matrix, of the aggression cases and controls in the training data, with 95% confidence ellipses.

The loadings of the aggression controls have been depicted in grey, and the loading scores of the cases have been depicted in black. The first 10 subfigures contain the loadings for the consensus across omics levels, subfigures 11-20 contain the loadings for the metabolomics data only, subfigures 21-30 contain the loadings for the DNA methylation data only (depicted as ‘epigenomics’ in figure), and subfigures 31-40 contain the loadings for the transmitted and non-transmitted polygenic scores only (PGSs; depicted as ‘genomics’ in figure).

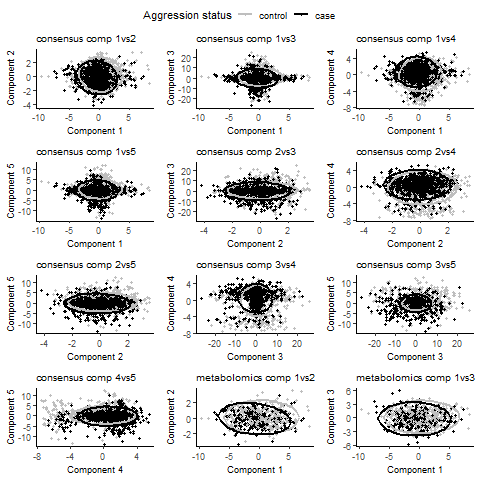

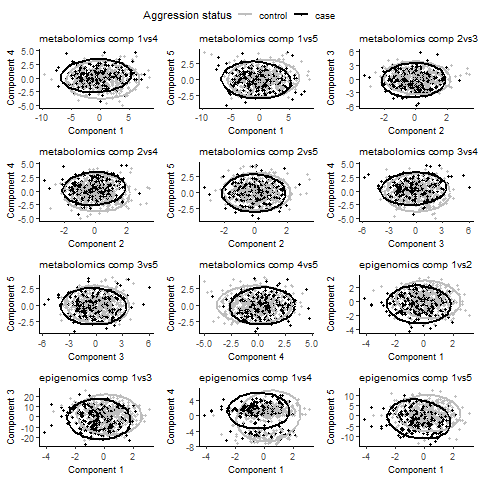

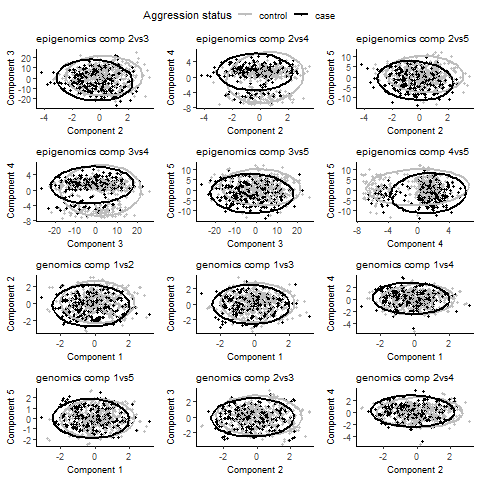

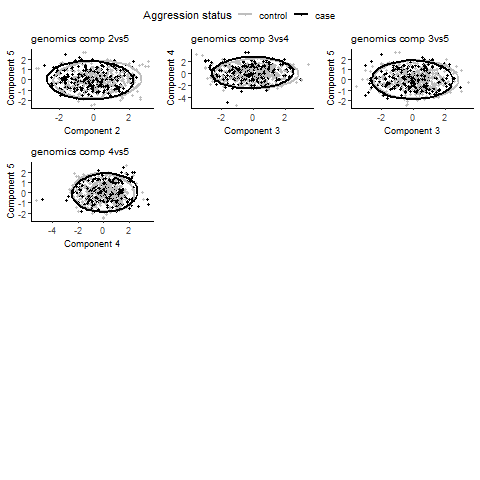

#### **Fig. S12.** Loadings of the 5-component multi-block sparse Partial Least Squares Discriminant Analysis (MB-sPLS-DA), with the empirical design matrix, of the aggression cases and controls as predicted in the test data, with 95% confidence ellipses from the training data.

The loadings of the aggression controls have been depicted in grey, and the loading scores of the cases have been depicted in black. The first 10 subfigures contain the loadings for the consensus across omics blocks, subfigures 11-20 contain the loadings for the metabolomics data only, subfigures 21-30 contain the loadings for the DNA methylation data only (depicted as ‘epigenomics’ in figure), and subfigures 31-40 contain the loadings for the transmitted and non-transmitted polygenic scores only (PGSs; depicted as ‘genomics’ in figure).

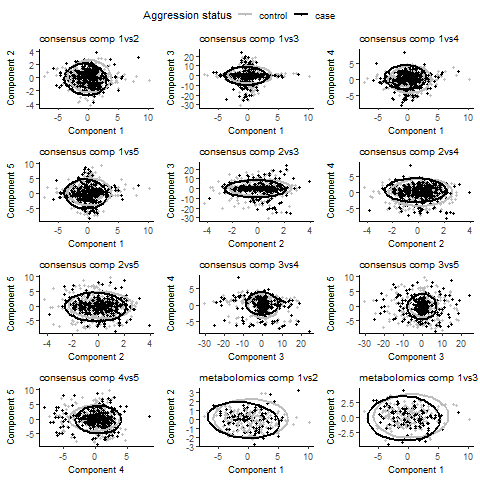

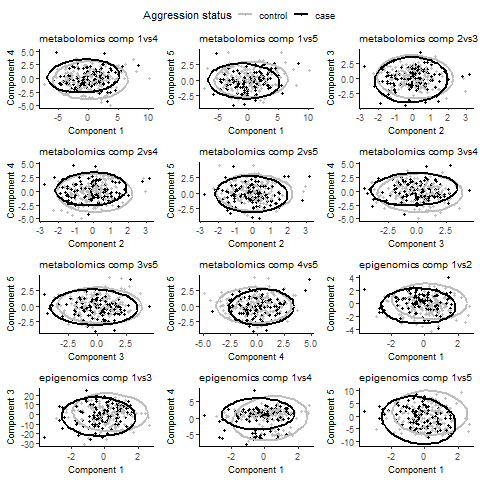

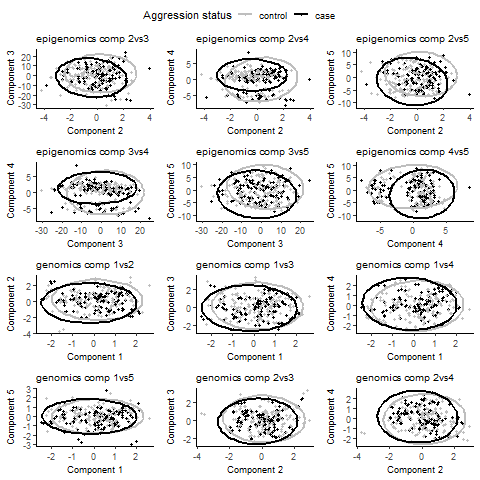

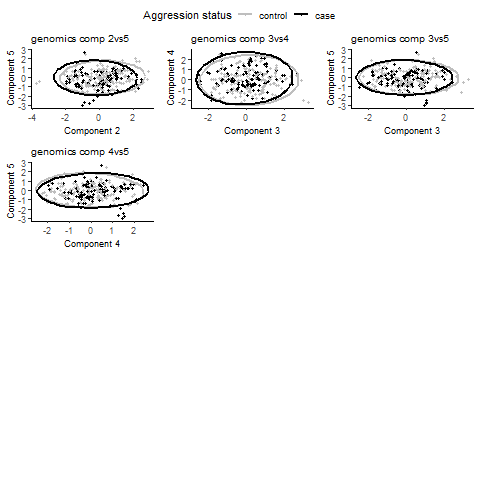

#### **Fig. S13.** Loadings of the 5-component multi-block sparse Partial Least Squares Discriminant Analysis (MB-sPLS-DA), with the empirical design matrix, of the aggression cases and controls as predicted in the clinical data, with 95% confidence ellipses from the training data.

The loadings of the aggression controls have been depicted in grey, and the loading scores of the cases have been depicted in black. The first 10 subfigures contain the loadings for the consensus across omics blocks, subfigures 11-20 contain the loadings for the metabolomics data only, subfigures 21-30 contain the loadings for the DNA methylation data only (depicted as ‘epigenomics’ in figure), and subfigures 31-40 contain the loadings for the transmitted and non-transmitted polygenic scores only (PGSs; depicted as ‘genomics’ in figure).

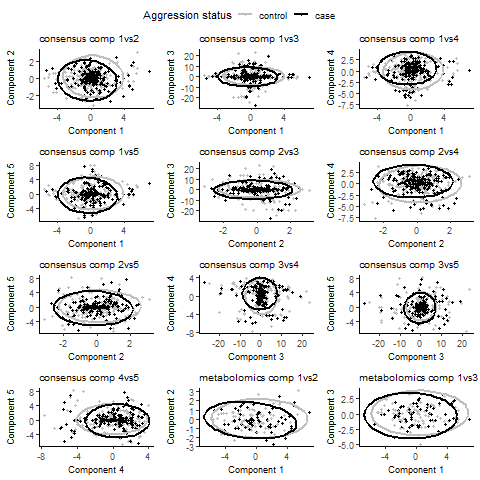

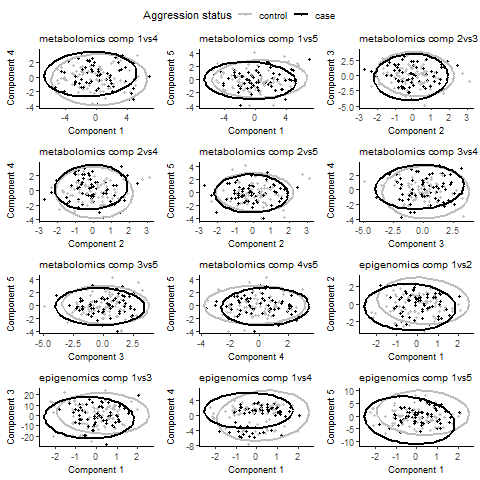

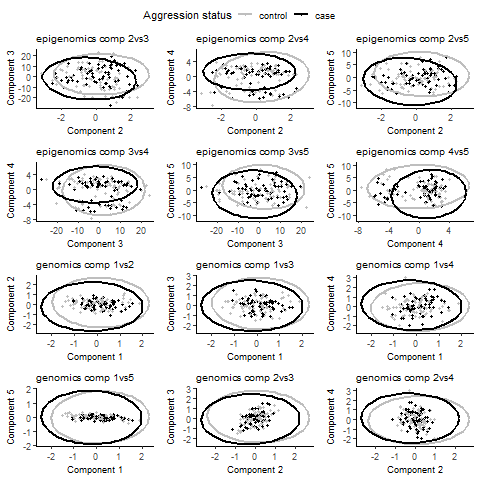

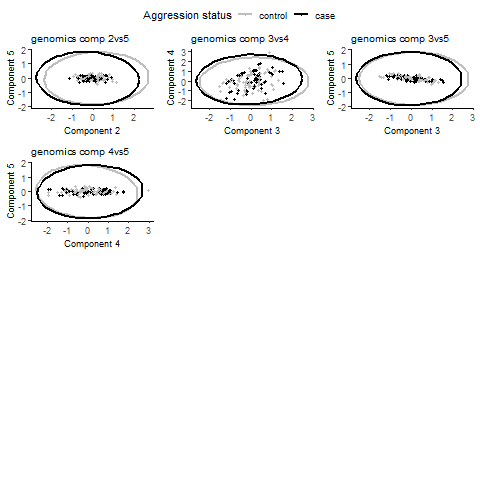

#### **Fig. S14.** Loadings of the 5-component multi-block sparse Partial Least Squares Discriminant Analysis (MB-sPLS-DA), with the null design matrix, of the aggression cases and controls in the training data, with 95% confidence ellipses.

The loadings of the aggression controls have been depicted in grey, and the loading scores of the cases have been depicted in black. The first 10 subfigures contain the loadings for the consensus across omics blocks, subfigures 11-20 contain the loadings for the metabolomics data only, subfigures 21-30 contain the loadings for the DNA methylation data only (depicted as ‘epigenomics’ in figure), and subfigures 31-40 contain the loadings for the transmitted and non-transmitted polygenic scores only (PGSs; depicted as ‘genomics’ in figure).

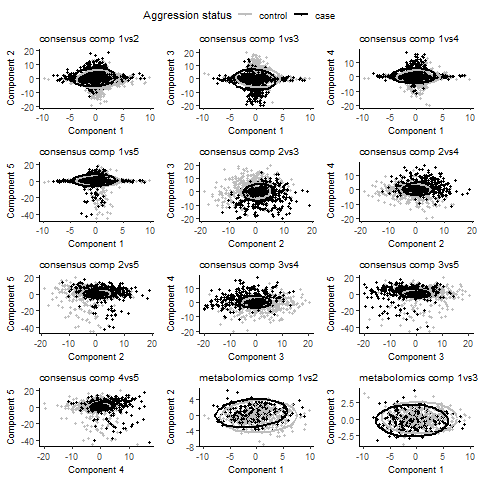

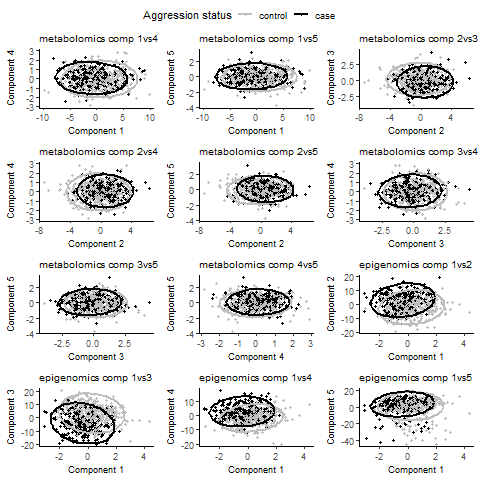

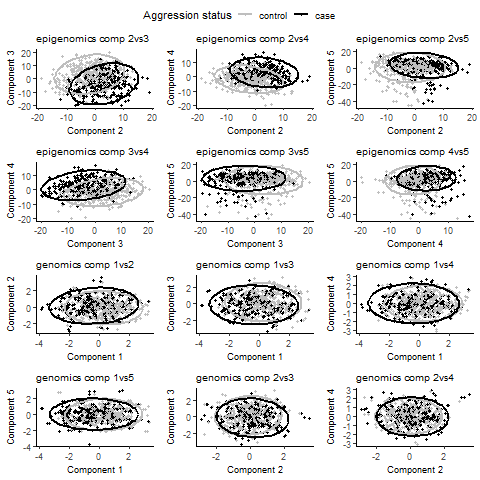

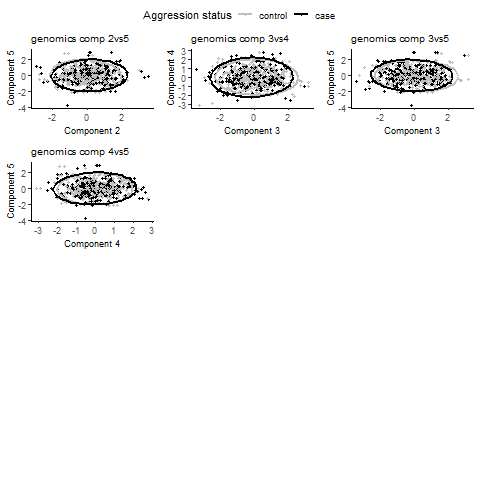

#### **Fig. S15.** Loadings of the 5-component multi-block sparse Partial Least Squares Discriminant Analysis (MB-sPLS-DA), with the null design matrix, of the aggression cases and controls as predicted in the test data, with 95% confidence ellipses from the training data.

The loadings of the aggression controls have been depicted in grey, and the loading scores of the cases have been depicted in black. The first 10 subfigures contain the loadings for the consensus across omics blocks, subfigures 11-20 contain the loadings for the metabolomics data only, subfigures 21-30 contain the loadings for the DNA methylation data only (depicted as ‘epigenomics’ in figure), and subfigures 31-40 contain the loadings for the transmitted and non-transmitted polygenic scores only (PGSs; depicted as ‘genomics’ in figure).

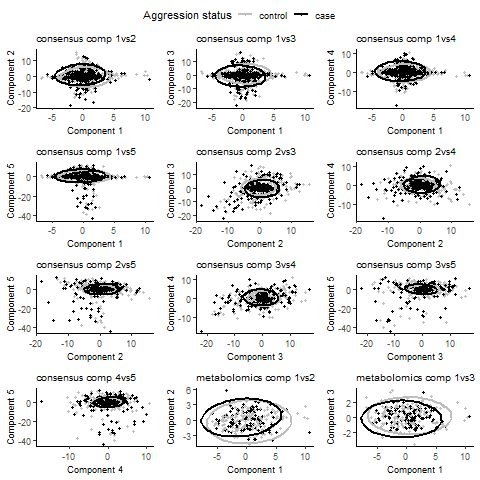

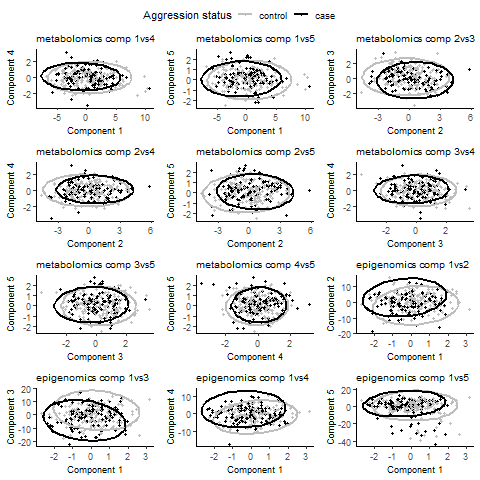

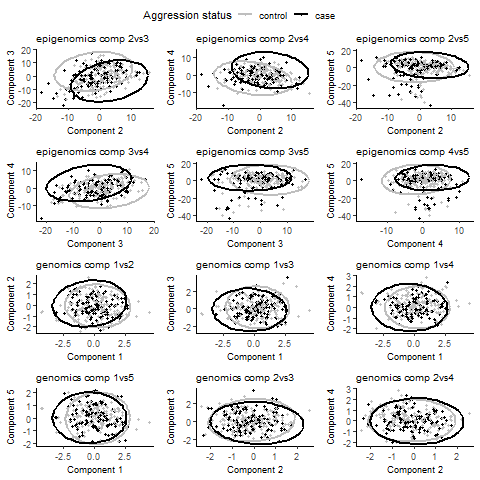

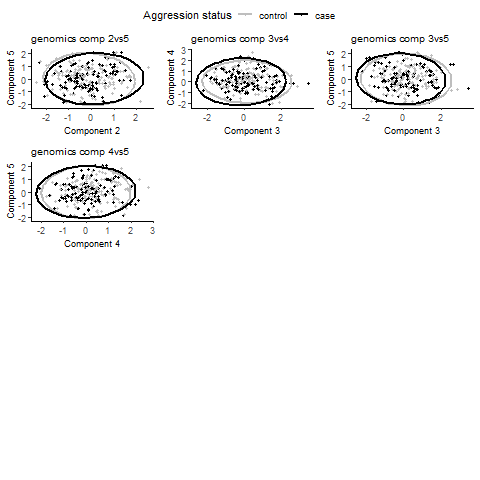

#### **Fig. S16.** Loadings of the 5-component multi-block sparse Partial Least Squares Discriminant Analysis (MB-sPLS-DA), with the null design matrix, of the aggression cases and controls as predicted in the clinical data, with 95% confidence ellipses from the training data.

The loadings of the aggression controls have been depicted in grey, and the loading scores of the cases have been depicted in black. The first 10 subfigures contain the loadings for the consensus across omics blocks, subfigures 11-20 contain the loadings for the metabolomics data only, subfigures 21-30 contain the loadings for the DNA methylation data only (depicted as ‘epigenomics’ in figure), and subfigures 31-40 contain the loadings for the transmitted and non-transmitted polygenic scores only (PGSs; depicted as ‘genomics’ in figure).

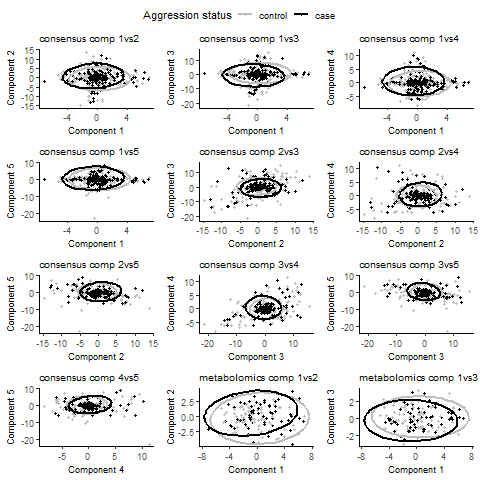

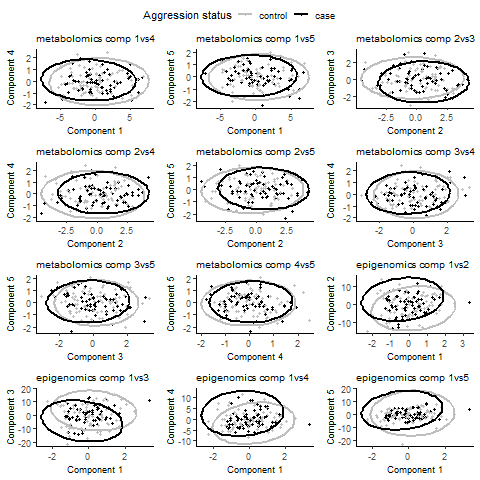

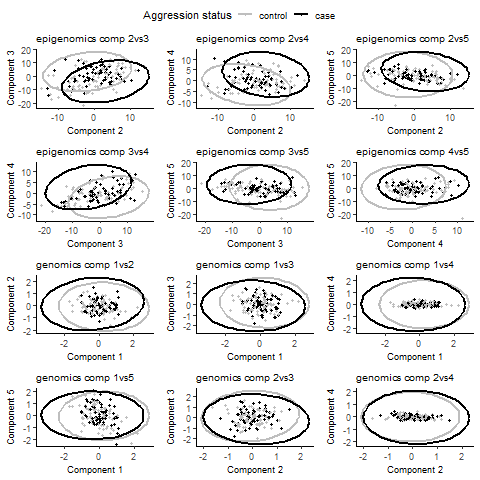

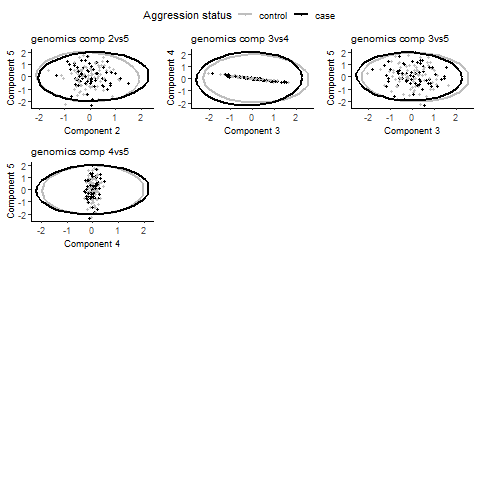

#### **Fig. S17.** Strong cross-omics connections of the multi-omics traits identified in the 5-component multi-block sparse Partial Least Squares Discriminant Analysis (MB-sPLS-DA) model including the null design matrix.

The outer ring depicts the metabolites, CpGs, and PGSs in green, pink, and yellow, respectively. For the metabolites, the ‘amines.’ prefix indicates these metabolites were measured on the Liquid Chromatography Mass Spectrometry (LC-MS) amines platform, and the ‘OA.’ prefix indicates these metabolites were measured on the Gas Chromatography (GC-) MS organic acids platform. For the polygenic scores (PGSs), the ‘_NTm’ suffix denotes the PGSs non-transmitted by mother, and Attention-Deficit Hyperactivity Disorder as “ADHD”. The inner plot depicts the connections among the omics variables. Here, only high absolute correlations of the PLS variates (r ≥ 0.60) between variables of at least two omics blocks are depicted, with blue lines reflecting negative correlations and red lines positive correlations. Correlations of the PLS variates are averaged across all components in the MB-sPLS-DA model. The full data matrix is included in **Data S12** and the patterns are included in **Data S13**.

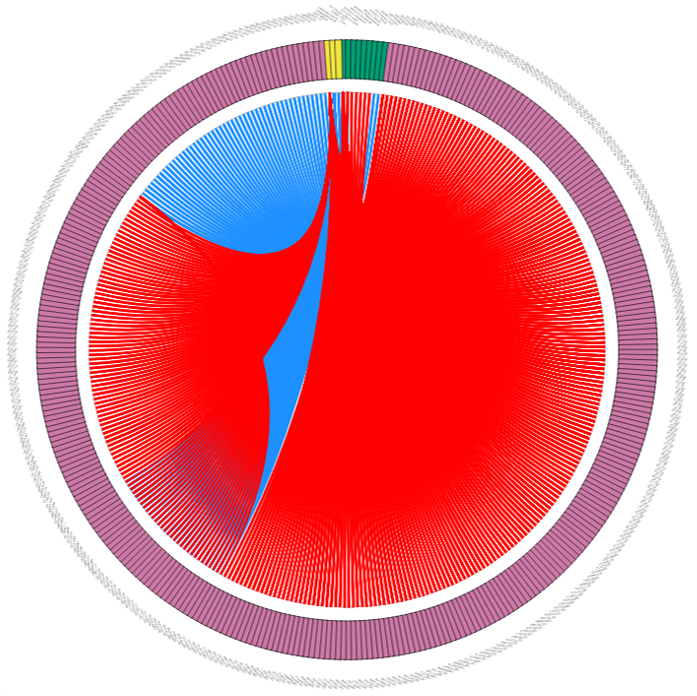

#### **Fig. S18.** Loadings of the 2-component multi-block sparse Partial Least Squares Discriminant Analysis (MB-sPLS-DA), with the full design matrix, of the aggression cases and controls in the training data, with 95% confidence ellipses.

The loadings of the aggression controls have been depicted in grey, and the loading scores of the cases have been depicted in black. The first subfigure contains the loadings for the consensus across omics blocks, subfigure two contains the loadings for the metabolomics data only, subfigure three contains the loadings for the DNA methylation data only (depicted as ‘epigenomics’ in figure), and subfigure four contains the loadings for the transmitted and non-transmitted polygenic scores only (PGSs; depicted as ‘genomics’ in figure).

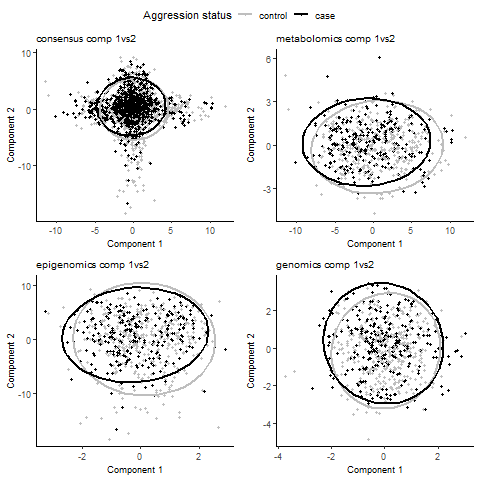

#### **Fig. S19.** Loadings of the 2-component multi-block sparse Partial Least Squares Discriminant Analysis (MB-sPLS-DA), with the full design matrix, of the aggression cases and controls as predicted in the test data, with 95% confidence ellipses from the training data.

The loadings of the aggression controls have been depicted in grey, and the loading scores of the cases have been depicted in black. The first subfigure contains the loadings for the consensus across omics blocks, subfigure two contains the loadings for the metabolomics data only, subfigure three contains the loadings for the DNA methylation data only (depicted as ‘epigenomics’ in figure), and subfigure four contains the loadings for the transmitted and non-transmitted polygenic scores only (PGSs; depicted as ‘genomics’ in figure).

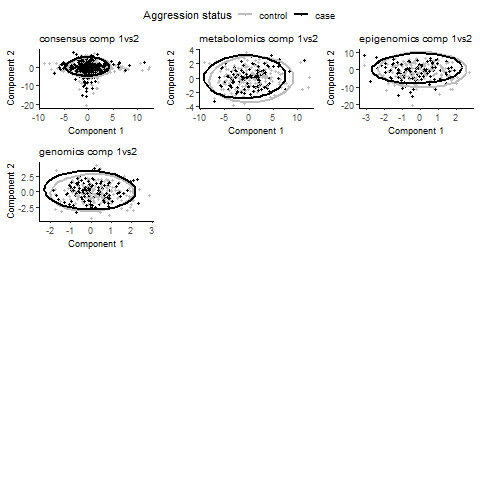

#### **Fig. S20.** Loadings of the 2-component multi-block sparse Partial Least Squares Discriminant Analysis (MB-sPLS-DA), with the full design matrix, of the aggression cases and controls as predicted in the clinical data, with 95% confidence ellipses from the training data.

The loadings of the aggression controls have been depicted in grey, and the loading scores of the cases have been depicted in black. The first subfigure contains the loadings for the consensus across omics blocks, subfigure two contains the loadings for the metabolomics data only, subfigure three contains the loadings for the DNA methylation data only (depicted as ‘epigenomics’ in figure), and subfigure four contains the loadings for the transmitted and non-transmitted polygenic scores only (PGSs; depicted as ‘genomics’ in figure).

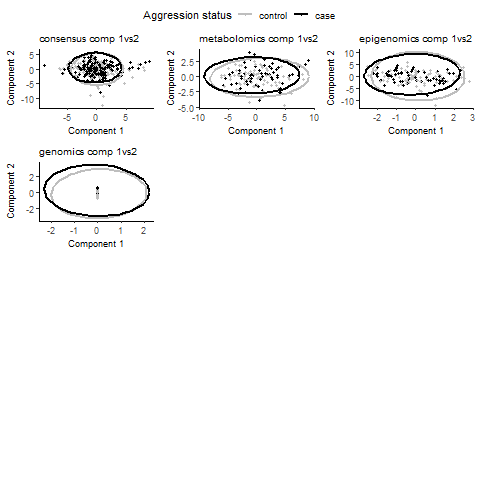

#### **Fig. S21.** Strong cross-omics connections of the multi-omics traits identified in the 2-component multi-block sparse Partial Least Squares Discriminant Analysis (MB-sPLS-DA) model including the full design matrix.

The outer ring depicts the metabolites, CpGs, and PGSs in green, pink, and yellow, respectively. For the metabolites, the ‘amines.’ prefix indicates these metabolites were measured on the Liquid Chromatography Mass Spectrometry (LC-MS) amines platform, and the ‘steroids.’ prefix indicates these metabolites were measured on the LC-MS steroids platform. For the polygenic scores (PGSs), Attention-Deficit Hyperactivity Disorder is abbreviated as “ADHD”, and the ‘_NTf’ suffix denotes the PGSs non-transmitted by father. The inner plot depicts the connections among the omics traits. Here, only high absolute correlations of the PLS variates (r ≥ 0.60) between variables of at least two omics blocks are depicted, with blue lines reflecting negative correlations and red lines positive correlations. Correlations are averaged across all components in the MB-sPLS-DA model. The full data matrix is included in **Data S15** and the patterns are included in **Table S19**.

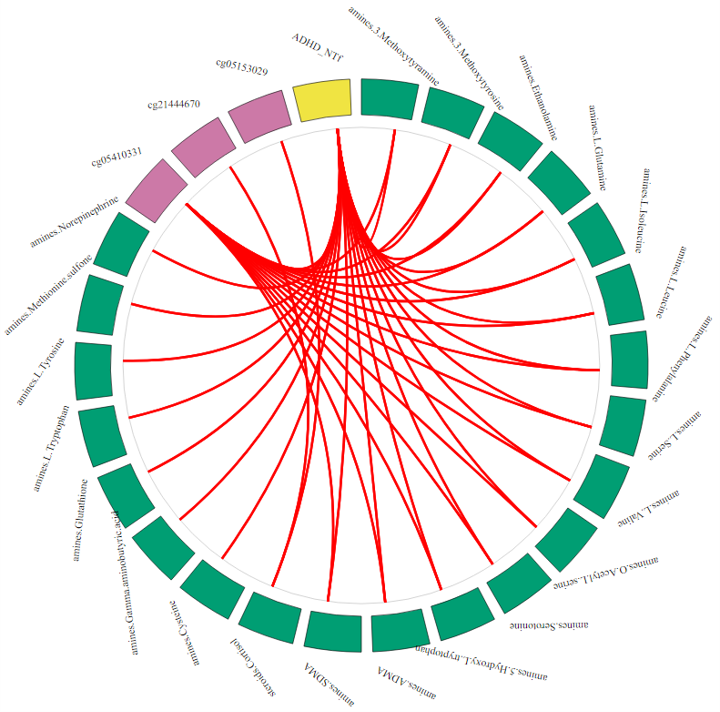

### **Supplementary Tables**

#### **Table S1.** Demographics for the twin and clinical cohorts with complete multi-omics data.

|  | **Twin cohort** | | | | **Clinical cohort** | | |
| --- | --- | --- | --- | --- | --- | --- | --- |
|  | **Controls** | **Cases** | **Total** | **Controls** | | **Cases** | **Total** |
| *N* (%) | 534 (57.9%) | 388 (42.1%) | 922 (100%) | 78 (54.9%) | | 64 (45.1%) | 142 (100%) |
| *N* (%) complete twin pairs | 184 | 115 | 421 | - | | - | - |
| Mean (*SD*) age | 9.4 (1.9) | 9.6 (1.9) | 9.5 (1.9) | 10.8 (1.7) | | 9.6 (1.7) | 10.2 (1.8) |
| Range age | 5.6 - 12.7 | 5.8 - 12.9 | 5.6 - 12.9 | 6.5 - 13.4 | | 6.3 - 13.3 | 6.3 - 13.4 |
| *N* (%) females | 280 (52.4%) | 171 (44.1%) | 451 (48.9%) | 20 (25.6%) | | 19 (29.7%) | 39 (27.5%) |
| *N* (%) MZ twins | 417 (78.1%) | 336 (86.6%) | 753 (81.7%) | - | | - | - |
| Mean (*SD*) aggression score ^1^ | 3.2 (4.1) | 7.4 (6.0) | 5.0 (5.4) | 6.1 (3.4) | | 21.1 (4.8) | 12.9 (8.5) |

Notes: MZ, monozygotic.

^1^ Measured with the mother-rated Aggressive Behavior syndrome scale of the Achenbach System of Empirically Based Assessment (ASEBA) Child Behavior Checklist (CBCL). The ASEBA CBCL Aggressive Behavior scores in the clinical cohort include 90% mother report and 10% father report.

#### **Table S2.** Number of Principal Components (PCs) that capture the dimensionality of the three omics blocks.

This table provides the number of traits per omics block. For each omics block the number of PCs that explain 80% or more of the cumulative variance, and the number of PCs with eigenvalues larger or equal to 1 are given. Furthermore, the optimal number of PCs based on visual inspection of the scree plots is provided. See **Fig. S1** for the scree plots for each of the omics block.

|  | **N traits** | **N PCs 80% cum. Var.** | **N PCs eigenvalue ≥1** | **N PCs scree plot** |
| --- | --- | --- | --- | --- |
| Metabolomics | 90 | 27 | 22 | ~8 |
| DNA methylation | 78,772 | 226 | 644 | ~6 |
| Polygenic scores | 45 | 26 | 16 | ~4 |

**Table S3.** Overview of the number of components and omics variables per component to retain in the (sparse) Partial Least Squares Discriminant Analyses ((s)PLS-DA) for the three omics blocks.

For the PLS-DA analyses the number of components used to initialize the models is given, and the optimal number of components (N comp) selected through 10-fold Cross Validation (CV) with 100 repeats. For the tuning of the sPLS-DA models, the number of components and number of omics variables per component to initialize the model tuning is given, followed by the optimal number of omics variables per component and optimal number of components selected through 10-fold CV with 100 repeats. For the final sPLS-DA models the number of components and number of omics variables per component are given, followed by the optimal number of components selected through 10-fold CV with 100 repeats.

|  |  | **PLS-DA** | | **Tune sPLS-DA** | | | | | **Final sPLS-DA** | | |
| --- | --- | --- | --- | --- | --- | --- | --- | --- | --- | --- | --- |
|  | **N variables** | **N comp** | **Optimal N comp** | **N comp** | **Prediction distance** | **N variables per comp** | **Optimal N variables/comp** | **Optimal N comp** | **N comp** | **N variables per comp** | **Optimal N comp** |
| Metabolomics | 90 | 8 | 3 | 3 | Centroids | 1 through 90 | 1, 89, 1 | 1 | 2 | 1, 89 | 1 |
| DNA methylation | 78,772 | 6 | 6 | 6 | Mahalanobis | 1, 3, 5, 7, 9, 11, 13, 15, 17, 19, 20, 30, 40, 50, 60, 70, 80, 90, 100, 150, 200, 250, 300, 350, 400, 450, 500, 550, 600, 650, 700, 750, 800, 850, 900, 950, 1000, 2000, 3000, 4000, 5000, 6000, 7000, 8000, 9000, 10000, 11000, 12000, 13000, 14000, 15000, 16000, 17000, 18000, 19000, 20000, 21000, 22000, 23000, 24000, 25000, 26000, 27000, 28000, 29000, 30000, 31000, 32000, 33000, 34000, 35000, 36000, 37000, 38000, 39000 | 7, 5, 800, 60, 450, 300 | 6 | 6 | 7, 5, 800, 60, 450, 300 | 6 |
| Polygenic scores | 45 | 4 | 1 | 2 | Maximum | 1 through 45 | 35, 1 | 1 | 2 | 35, 1 | 1 |

#### **Table S4.** Pearson correlations (*r*) of the Partial Least Squares (PLS) variates of the pairwise cross-omics models.

The correlations were obtained for two models: model 1, which included all omics variables, and model 2, which included only the omics variables that were selected by the single-omics sPLS-DA models (see **Data S2**). A False Discovery Rate (FDR) of 5% for 3 models was used to correct for multiple testing separately for the model 1 and model 2 analyses, setting the significance threshold to q ≤ 0.05.

| **Model (omics layer X and Y)** | **Model 1** | | | **Model 2** | | |
| --- | --- | --- | --- | --- | --- | --- |
|  | ***r*** | ***p*** | ***q*** | ***r*** | ***p*** | ***q*** |
| DNA methylation & Metabolomics | 0.18 | 3.19x10^-15^ | 3.19x10^-15^ | 0.18 | 6.07x10^-15^ | 6.07x10^-15^ |
| Polygenic scores & Metabolomics | 0.28 | 5.31x10^-24^ | 7.97x10^-24^ | 0.28 | 3.20x10^-24^ | 9.57x10^-24^ |
| Polygenic scores & DNA methylation | 0.29 | 7.66x10^-27^ | 2.30x10^-26^ | 0.28 | 6.38x10^-24^ | 9.57x10^-24^ |

#### **Table S5**. Overview of the number of components and number of omics variables per component to retain in the multi-omics (sparse) multi-block Partial Least Squares Discriminant Analyses (MB-(s)PLS-DA).

For the MB-PLS-DA analyses the number of components used to initialize the model is given, this is based on the number of components in the final sPLS-DA single-omics models with the largest number of components (i.e., 6 components for the epigenomics data; see **Table S3**). The optimal number of components (N comp) selected through 10-fold Cross Validation (CV) with 100 repeats is also provided for the MB-PLS-DA analyses. For the tuning of the MB-sPLS-DA models, the number of components and number of omics variables per component to initialize the model tuning is given, followed by the optimal number of omics variables per component and optimal number of components selected through 5-fold CV with 50 repeats. For the final MB-sPLS-DA models the number of components and number of omics variables per component are given, followed by the optimal number of components selected through 5-fold CV with 50 repeats.

| **Data** | | | **MB-PLS-DA** | | **Tune MB-sPLS-DA** | | | | | **Final MB-sPLS-DA** | | |
| --- | --- | --- | --- | --- | --- | --- | --- | --- | --- | --- | --- | --- |
| **Design matrix** | **Omics block** | **N variables** | **N comp** | **Optimal N comp** | **N comp** | **Prediction distance** | **N variables/comp** | **Optimal N variables/comp** | **Optimal N comp** | **N comp** | **N variables/comp** | **Optimal N comp** |
| Empirical | Metabolomics | 90 | 6 | 6 | 6 | Mahalanobis | 1, 5, 30, 90 | 30, 1, 90, 90, 5 | 5 | 5 | 30, 1, 90, 90, 5 | 5 |
|  | DNA methylation | 78,772 |  |  |  |  | 5, 7, 60, 350, 800 | 5, 5, 350, 60, 350 |  |  | 5, 5, 350, 60, 350 |  |
|  | Polygenic scores | 45 |  |  |  |  | 1, 5, 20, 35 | 1, 35 |  |  | 1, 35 |  |
| Null | Metabolomics | 90 | 6 | 6 | 6 | Mahalanobis | 1, 5, 30, 90 | 90, 90, 30, 1, 1 | 5 | 5 | 90, 90, 30, 1, 1 | 5 |
|  | DNA methylation | 78,772 |  |  |  |  | 5, 7, 60, 350, 800 | 7, 800, 350, 350, 350 |  |  | 7, 800, 350, 350, 350 |  |
|  | Polygenic scores | 45 |  |  |  |  | 1, 5, 20, 35 | 35, 5 |  |  | 35, 5 |  |
| Full | Metabolomics | 90 | 6 | 6 | 6 | Mahalanobis | 1, 5, 30, 90 | 90, 30, 5, 5, 90 | 2 | 2 | 90, 30, 5, 5, 90 | 2 |
|  | DNA methylation | 78,772 |  |  |  |  | 5, 7, 60, 350, 800 | 5, 60, 5, 7, 7 |  |  | 5, 60, 5, 7, 7 |  |
|  | Polygenic scores | 45 |  |  |  |  | 1, 5, 20, 35 | 1, 35 |  |  | 1, 35 |  |

**Table S6**. EWAS atlas enrichment analysis results for all CpGs selected into the DNA methylation sparse Partial Least Squares Discriminant Analysis (sPLS-DA) model.

Enriched traits based on enrichment analysis with 1,614 CpGs selected by the 6-component DNA methylation sPLS-DA model. The fourth column (DMC) shows how many of the 1,614 CpGs have been previously associated with the trait in the first column. The fifth column (background) shows how many CpGs have previously been associated with the trait in column 1. The last column (%) shows the percentage of CpGs previously associated with the trait in column 1 that were also selected by the DNA methylation sPLS-DA model.

| **Trait** | **OR** | ***p*** | **DMC** | **Background** | **%** |
| --- | --- | --- | --- | --- | --- |
| glucocorticoid exposure | 18.02 | 5.34E-158 | 96 | 3468 | 2.8% |
| household socioeconomic status in childhood | 9.88 | 1.18E-13 | 10 | 620 | 1.6% |
| systemic lupus erythematosus (SLE) | 2.46 | 4.06E-10 | 32 | 7848 | 0.4% |
| Psoriasis | 2.63 | 5.04E-07 | 18 | 4138 | 0.4% |
| ancestry | 1.90 | 7.31E-07 | 36 | 10618 | 0.3% |
| gestational diabetes mellitus | 2.19 | 1.29E-06 | 24 | 6599 | 0.4% |
| breast cancer risk | 54.53 | 1.72E-06 | 2 | 24 | 8.3% |
| infertility | 2.17 | 1.85E-05 | 19 | 5281 | 0.4% |
| Claes-Jensen syndrome | 4.03 | 1.88E-05 | 7 | 1050 | 0.7% |
| asthma | 1.55 | 8.10E-05 | 36 | 13639 | 0.3% |
| Severe acute malnutrition | 119.55 | 2.30E-04 | 1 | 6 | 16.7% |
| post- to prefenofibrate treatment ratio for high sensitivity CRP | 99.81 | 3.06E-04 | 1 | 7 | 14.3% |
| Parkinson's disease (PD) | 6.55 | 4.03E-04 | 3 | 278 | 1.1% |
| leukoaraiosis (LA) | 4.56 | 5.06E-04 | 4 | 531 | 0.8% |
| t(1;11) translocation | 49.96 | 9.80E-04 | 1 | 13 | 7.7% |
| primary Sjögren’s Syndrome (pSS) | 2.05 | 1.17E-03 | 12 | 3526 | 0.3% |
| polycystic ovary syndrome (PCOS) | 8.89 | 1.28E-03 | 2 | 137 | 1.5% |
| osteoarthritis (OA) | 2.58 | 1.56E-03 | 7 | 1634 | 0.4% |
| ankylosis spondylitis (AS) | 7.36 | 2.49E-03 | 2 | 165 | 1.2% |
| renal clear cell carcinoma survival | 27.28 | 2.91E-03 | 1 | 23 | 4.3% |
| Alzheimer's disease (AD) | 2.60 | 3.09E-03 | 6 | 1392 | 0.4% |
| Facial anomalies syndrome (ICF) | 6.71 | 3.45E-03 | 2 | 181 | 1.1% |
| occupational pesticide exposure | 22.22 | 4.23E-03 | 1 | 28 | 3.6% |
| maternal socioeconomic status | 20.69 | 4.82E-03 | 1 | 30 | 3.3% |
| Down syndrome | 1.33 | 8.30E-03 | 38 | 14649 | 0.3% |
| follicular thyroid carcinoma | 1.61 | 9.58E-03 | 15 | 5575 | 0.3% |

#### **Table S7**. Prediction parameters of the sparse Partial Least Squares Discriminant Analyses (sPLS-DA) models in the test and clinical data.

Balanced error rates (BER), prediction sensitivity, specificity, and accuracy, and model Area Under the Curve (AUC) of the prediction of the sPLS-DA models in the test and clinical data per component. The AUC *p*-values have been adjusted separately for the test and clinical data for multiple testing using the FDR of 5% for 10 tests (*q*).

| **Block** | **Component** | **BER** | **Sensitivity** | **Specificity** | **Accuracy** | **AUC** | **AUC *p*** | **AUC *q*** |
| --- | --- | --- | --- | --- | --- | --- | --- | --- |
| **Test data** | | | | | | | | |
| Metabolomics | 1 | 0.52 | 47.9 | 48.1 | 48.0 | 0.48 | 0.54 | 0.84 |
| Metabolomics | 2 | 0.54 | 44.4 | 48.1 | 46.6 | 0.48 | 0.54 | 0.84 |
| DNA methylation | 1 | 0.44 | 52.1 | 60.6 | 57.0 | 0.58 | 0.02 | 0.19 |
| DNA methylation | 2 | 0.51 | 47.0 | 51.2 | 49.5 | 0.50 | 0.97 | 0.97 |
| DNA methylation | 3 | 0.49 | 52.1 | 49.4 | 50.5 | 0.51 | 0.75 | 0.84 |
| DNA methylation | 4 | 0.46 | 52.1 | 55.0 | 53.8 | 0.52 | 0.63 | 0.84 |
| DNA methylation | 5 | 0.47 | 53.8 | 51.2 | 52.3 | 0.52 | 0.52 | 0.84 |
| DNA methylation | 6 | 0.45 | 52.1 | 57.5 | 55.2 | 0.53 | 0.43 | 0.84 |
| Polygenic scores | 1 | 0.50 | 23.1 | 76.2 | 53.8 | 0.52 | 0.51 | 0.84 |
| Polygenic scores | 2 | 0.51 | 20.5 | 76.9 | 53.1 | 0.51 | 0.76 | 0.84 |
| **Clinical data** | | | | | | | | |
| Metabolomics | 1 | 0.48 | 50.0 | 53.8 | 52.1 | 0.49 | 0.84 | 0.93 |
| Metabolomics | 2 | 0.51 | 48.4 | 48.7 | 48.6 | 0.51 | 0.81 | 0.93 |
| DNA methylation | 1 | 0.49 | 50.0 | 51.3 | 50.7 | 0.50 | 0.97 | 0.97 |
| DNA methylation | 2 | 0.48 | 46.9 | 56.4 | 52.1 | 0.49 | 0.76 | 0.93 |
| DNA methylation | 3 | 0.55 | 42.2 | 48.7 | 45.8 | 0.44 | 0.22 | 0.37 |
| DNA methylation | 4 | 0.58 | 35.9 | 48.7 | 43.0 | 0.40 | 0.04 | 0.22 |
| DNA methylation | 5 | 0.58 | 37.5 | 47.4 | 43.0 | 0.39 | 0.03 | 0.22 |
| DNA methylation | 6 | 0.55 | 37.5 | 52.6 | 45.8 | 0.43 | 0.14 | 0.28 |
| Polygenic scores | 1 | 0.51 | 7.8 | 89.7 | 52.8 | 0.58 | 0.10 | 0.26 |
| Polygenic scores | 2 | 0.50 | 12.5 | 87.2 | 53.5 | 0.58 | 0.10 | 0.26 |

#### **Table S8**. EWAS atlas enrichment analysis results for the CpGs included in cluster 1 and 2 of the DNA methylation-metabolomics Partial Least Squares (PLS) model.

Enriched traits based on enrichment analysis with 1,151 CpGs that were included in cluster 1 and 463 CpGs that were included in cluster 2 of the 3-component DNA methylation-metabolomics PLS model. Cluster assignment is provided in **Data S4**. The fourth column (DMC) shows how many of the CpGs have been previously associated with the trait in the first column. The fifth column (background) shows how many CpGs have previously been associated with the trait in column 1. The last column (%) shows the percentage of CpGs previously associated with the trait in column 1 that were also included in cluster 1 or cluster 2 of the DNA methylation-metabolomics PLS model.

| **trait** | **OR** | **p** | **DMC** | **Background** | **%** |
| --- | --- | --- | --- | --- | --- |
| **Cluster 1** | | | | | |
| household socioeconomic status in childhood | 12.48 | 4.20E-14 | 9 | 620 | 1.5% |
| psoriasis | 3.70 | 1.18E-10 | 18 | 4138 | 0.4% |
| ancestry | 2.46 | 2.12E-10 | 33 | 10618 | 0.3% |
| gestational diabetes mellitus | 2.96 | 4.92E-10 | 23 | 6599 | 0.3% |
| systemic lupus erythematosus (SLE) | 2.71 | 1.77E-09 | 25 | 7848 | 0.3% |
| asthma | 2.04 | 5.00E-09 | 33 | 13639 | 0.2% |
| down syndrome | 1.88 | 1.75E-07 | 38 | 14649 | 0.3% |
| Claes-Jensen syndrome | 5.67 | 4.14E-07 | 7 | 1050 | 0.7% |
| breast cancer risk | 76.62 | 4.54E-07 | 2 | 24 | 8.3% |
| leukoaraiosis (LA) | 6.40 | 5.26E-05 | 4 | 531 | 0.8% |
| osteoarthritis (OA) | 3.63 | 5.68E-05 | 7 | 1634 | 0.4% |
| follicular thyroid carcinoma | 2.27 | 5.77E-05 | 15 | 5575 | 0.3% |
| Parkinson's disease (PD) | 9.20 | 6.62E-05 | 3 | 278 | 1.1% |
| atopy | 2.18 | 7.22E-05 | 16 | 6198 | 0.3% |
| severe acute malnutrition | 168.18 | 1.17E-04 | 1 | 6 | 16.7% |
| infertility | 2.24 | 1.26E-04 | 14 | 5281 | 0.3% |
| post- to prefenofibrate treatment ratio for high sensitivity CRP | 140.38 | 1.56E-04 | 1 | 7 | 14.3% |
| Alzheimer's disease (AD) | 3.65 | 1.76E-04 | 6 | 1392 | 0.4% |
| papillary thyroid carcinoma | 2.09 | 2.46E-04 | 15 | 6073 | 0.2% |
| polycystic ovary syndrome (PCOS) | 12.49 | 3.65E-04 | 2 | 137 | 1.5% |
| ankylosis spondylitis (AS) | 10.34 | 7.28E-04 | 2 | 165 | 1.2% |
| exercise | 3.07 | 7.90E-04 | 6 | 1652 | 0.4% |
| facial anomalies syndrome (ICF) | 9.42 | 1.02E-03 | 2 | 181 | 1.1% |
| maternal smoking | 1.92 | 1.30E-03 | 14 | 6153 | 0.2% |
| renal clear cell carcinoma survival | 38.29 | 1.50E-03 | 1 | 23 | 4.3% |
| occupational pesticide exposure | 31.20 | 2.19E-03 | 1 | 28 | 3.6% |
| Kabuki syndrome (KS) | 2.68 | 2.41E-03 | 6 | 1891 | 0.3% |
| maternal socioeconomic status | 29.05 | 2.50E-03 | 1 | 30 | 3.3% |
| preterm birth | 1.58 | 3.15E-03 | 19 | 10662 | 0.2% |
| breastfeeding | 6.74 | 3.35E-03 | 2 | 252 | 0.8% |
| folic acid and vitamin B12 supplementation | 6.20 | 4.50E-03 | 2 | 274 | 0.7% |
| type 2 diabetes (T2D) | 1.80 | 5.95E-03 | 12 | 5633 | 0.2% |
| SETD1B-related syndrome | 2.17 | 6.93E-03 | 7 | 2721 | 0.3% |
| maternal Hepatitis B virus (HBV) infection | 5.11 | 8.69E-03 | 2 | 332 | 0.6% |
| IL-13 treatment | 2.81 | 9.18E-03 | 4 | 1203 | 0.3% |
| myalgic encephalomyelitis/chronic fatigue syndrome | 1.68 | 9.40E-03 | 13 | 6500 | 0.2% |
| **Cluster 2** | | | | | |
| glucocorticoid exposure | 51.25 | 1.00E-308 | 92 | 3468 | 2.7% |
| t(1;11) translocation | 74.23 | 1.68E-11 | 1 | 13 | 7.7% |
| substance-use risk | 16.25 | 1.91E-05 | 1 | 65 | 1.5% |
| treated with assisted reproductive technology | 10.44 | 1.49E-04 | 1 | 118 | 0.8% |
| maternal hypertensive disorders in pregnancy | 3.12 | 8.55E-03 | 1 | 922 | 0.1% |

#### **Table S9.** Hierarchical cluster assignment for the 36 polygenic scores (PGSs) and for the 90 metabolites in the 2-component PGSs-metabolomics Partial Least Squares (PLS) model.

The hierarchical clustering was generated using the Ward linkage algorithm on Euclidean distances of the PLS variates. For the PGSs, the ‘_NTm’ suffix denotes PGSs non-transmitted by mother, the ‘_NTf’ suffix denotes the PGSs non-transmitted by father, and childhood aggression is abbreviated as “aggression”, Attention-Deficit Hyperactivity Disorder as “ADHD”, Major Depressive Disorder as “MDD”, Autism Spectrum Disorder as “Autism”, Educational Attainment as “EA”, and wellbeing spectrum as “wellbeing”. For the metabolites, the ‘amines.’ prefix indicates these metabolites were measured on the Liquid Chromatography Mass Spectrometry (LC-MS) amines platform, the ‘steroids.’ prefix indicates these metabolites were measured on the LC-MS steroids platform, and the ‘OA.’ prefix indicates these metabolites were measured on the Gas Chromatography (GC-) MS organic acids platform.

| **metabolites** | **Clusters metabolites** | **PGSs** | **Clusters PGSs** |
| --- | --- | --- | --- |
| amines.1.Methylhistidine | 1 | AgeAtFirstBirth | 1 |
| amines.3.Methoxytyramine | 1 | ADHD | 1 |
| amines.3.Methoxytyrosine | 1 | ADHD_NTm | 1 |
| amines.Citrulline | 1 | ADHD_NTf | 1 |
| amines.Cysteine | 1 | Aggression | 1 |
| amines.Ethanolamine | 1 | Aggression_NTm | 1 |
| amines.Gamma.aminobutyric.acid | 1 | Aggression_NTf | 1 |
| amines.Glutathione | 1 | Autism_NTm | 1 |
| amines.Glycine | 1 | Autism_NTf | 1 |
| amines.Glycylglycine | 1 | CigarettesPerDay | 1 |
| amines.Glycylproline | 1 | CigarettesPerDay_NTm | 1 |
| amines.L.Alanine | 1 | CigarettesPerDay_NTf | 1 |
| amines.L.Alpha.aminobutyric.acid | 1 | EA | 1 |
| amines.L.Arginine | 1 | Insomnia | 1 |
| amines.L.Asparagine | 1 | Insomnia_NTm | 1 |
| amines.L.Aspartic.acid | 1 | loneliness_NTm | 1 |
| amines.L.Glutamic.acid | 1 | MDD | 1 |
| amines.L.Glutamine | 1 | MDD_NTm | 1 |
| amines.L.Histidine | 1 | MDD_NTf | 1 |
| amines.L.Homoserine | 1 | Agesmokinginitiation_NTm | 1 |
| amines.L.Isoleucine | 1 | smokinginitiation_NTm | 1 |
| amines.L.Kynurenine | 1 | smokinginitiation_NTf | 1 |
| amines.L.Leucine | 1 | wellbeing | 1 |
| amines.L.Methionine | 1 | AgeAtFirstBirth_NTm | 2 |
| amines.L.Methionine.sulfoxide | 1 | EA_NTm | 2 |
| amines.L.Phenylalanine | 1 | childhoodIQ_NTm | 2 |
| amines.L.Proline | 1 | childhoodIQ_NTf | 2 |
| amines.L.Serine | 1 | intelligence_NTm | 2 |
| amines.L.Threonine | 1 | intelligence_NTf | 2 |
| amines.L.Tryptophan | 1 | loneliness | 2 |
| amines.L.Tyrosine | 1 | Agesmokinginitiation | 2 |
| amines.L.Valine | 1 | smokinginitiation | 2 |
| amines.Methionine.sulfone | 1 | selfreportedhealth_NTm | 2 |
| amines.Norepinephrine | 1 | selfreportedhealth_NTf | 2 |
| amines.O.Acetyl.L.serine | 1 | wellbeing_NTm | 2 |
| amines.O.Phosphoethanolamine | 1 | wellbeing_NTf | 2 |
| amines.Ornithine | 1 |  |  |
| amines.S.Methylcysteine | 1 |  |  |
| amines.Sarcosine | 1 |  |  |
| amines.Serotonine | 1 |  |  |
| amines.5.Hydroxy.L.tryptophan | 1 |  |  |
| amines.ADMA | 1 |  |  |
| amines.Gamma.Glutamylglutamine | 1 |  |  |
| amines.Gamma.L.glutamyl.L.alanine | 1 |  |  |
| amines.O.Phosphoserine | 1 |  |  |
| amines.SDMA | 1 |  |  |
| amines.3.Methylhistidine | 2 |  |  |
| amines.Anserine | 2 |  |  |
| amines.Cystathionine | 2 |  |  |
| amines.DL.3.aminoisobutyric.acid | 2 |  |  |
| amines.Homocitrulline | 2 |  |  |
| amines.Homocysteine | 2 |  |  |
| amines.Hydroxylysine | 2 |  |  |
| amines.L.2.aminoadipic.acid | 2 |  |  |
| amines.L.4.hydroxy.proline | 2 |  |  |
| amines.L.Lysine | 2 |  |  |
| amines.N6.N6.N6.Trimethyl.L.lysine | 2 |  |  |
| amines.Putrescine | 2 |  |  |
| amines.Taurine | 2 |  |  |
| amines.Beta.Alanine | 2 |  |  |
| OA.2.hydroxybutyric.acid | 2 |  |  |
| OA.Adipic.acid | 2 |  |  |
| OA.Citric.acid | 2 |  |  |
| OA.Glutaric.acid | 2 |  |  |
| OA.Glycolic.acid | 2 |  |  |
| OA.Malic.acid | 2 |  |  |
| OA.Succinic.acid | 2 |  |  |
| OA.Fumaric.acid | 2 |  |  |
| OA.Methylmalonic.acid | 2 |  |  |
| OA.Pyroglutamic.acid | 2 |  |  |
| OA.Isocitrate | 2 |  |  |
| OA.3.Hydroxybutyric.acid | 2 |  |  |
| OA.3.Hydroxyisobutyric.acid | 2 |  |  |
| OA.3.hydroxyisovaleric.acid | 2 |  |  |
| OA.Glyceric.acid | 2 |  |  |
| OA.Uracil | 2 |  |  |
| OA.Homovanillic.acid | 2 |  |  |
| OA.Cis.aconitic.acid | 2 |  |  |
| OA.3.methyladipic.acid | 2 |  |  |
| OA.3.Hydroxypropionic.Acid | 2 |  |  |
| steroids.beta.Cortolone | 2 |  |  |
| steroids.alpha.Cortolone | 2 |  |  |
| steroids.Cortisol | 2 |  |  |
| steroids.Cortisone | 2 |  |  |
| steroids.Cortisone.sulfate | 2 |  |  |
| steroids.Dehydroepiandrosterone.sulfate..DHEA.S. | 2 |  |  |
| steroids.DihydroTestosterone.Glucuronide | 2 |  |  |
| steroids.Etiocholanolone.glucuronide | 2 |  |  |
| steroids.Etiocholanolone.sulfate | 2 |  |  |
| steroids..alpha.beta..Cortol | 2 |  |  |

#### **Table S10**. EWAS atlas enrichment analysis results for the CpGs included in cluster 1 and 2 of the polygenic scores (PGSs)-DNA methylation Partial Least Squares (PLS) model.

Enriched traits based on enrichment analysis with 1,142 CpGs that were included in cluster 1 and 472 CpGs that were included in cluster 2 of the 3-component PGSs-DNA methylation PLS model. Cluster assignment is provided in **Data S7**. The fourth column (DMC) shows how many of the CpGs have been previously associated with the trait in the first column. The fifth column (background) shows how many CpGs have previously been associated with the trait in column 1. The last column (%) shows the percentage of CpGs previously associated with the trait in column 1 that were also included in cluster 1 or cluster 2 of the DNA methylation-PGS PLS model.

| **Trait** | **OR** | ***p*** | **DMC** | **Background** | **%** |
| --- | --- | --- | --- | --- | --- |
| **Cluster1** | | | | | |
| household socioeconomic status in childhood | 12.58 | 3.68E-14 | 9 | 620 | 1.5% |
| psoriasis | 3.73 | 9.55E-11 | 18 | 4138 | 0.4% |
| ancestry | 2.48 | 1.55E-10 | 33 | 10618 | 0.3% |
| gestational diabetes mellitus | 2.99 | 3.87E-10 | 23 | 6599 | 0.3% |
| asthma | 2.06 | 3.55E-09 | 33 | 13639 | 0.2% |
| systemic lupus erythematosus (SLE) | 2.62 | 9.94E-09 | 24 | 7848 | 0.3% |
| Claes-Jensen syndrome | 5.72 | 3.78E-07 | 7 | 1050 | 0.7% |
| breast cancer risk | 77.21 | 4.40E-07 | 2 | 24 | 8.3% |
| down syndrome | 1.80 | 1.51E-06 | 36 | 14649 | 0.2% |
| leukoaraiosis (LA) | 6.45 | 4.98E-05 | 4 | 531 | 0.8% |
| follicular thyroid carcinoma | 2.29 | 5.03E-05 | 15 | 5575 | 0.3% |
| osteoarthritis (OA) | 3.66 | 5.23E-05 | 7 | 1634 | 0.4% |
| atopy | 2.20 | 6.27E-05 | 16 | 6198 | 0.3% |
| Parkinson's disease (PD) | 9.27 | 6.35E-05 | 3 | 278 | 1.1% |
| severe acute malnutrition | 169.56 | 1.15E-04 | 1 | 6 | 16.7% |
| post- to prefenofibrate treatment ratio for high sensitivity CRP | 141.48 | 1.54E-04 | 1 | 7 | 14.3% |
| Alzheimer's disease (AD) | 3.68 | 1.64E-04 | 6 | 1392 | 0.4% |
| papillary thyroid carcinoma | 2.10 | 2.16E-04 | 15 | 6073 | 0.2% |
| polycystic ovary syndrome (PCOS) | 12.59 | 3.55E-04 | 2 | 137 | 1.5% |
| infertility | 2.09 | 5.64E-04 | 13 | 5281 | 0.2% |
| ankylosis spondylitis (AS) | 10.42 | 7.07E-04 | 2 | 165 | 1.2% |
| exercise | 3.10 | 7.39E-04 | 6 | 1652 | 0.4% |
| facial anomalies syndrome (ICF) | 9.49 | 9.93E-04 | 2 | 181 | 1.1% |
| maternal smoking | 1.93 | 1.16E-03 | 14 | 6153 | 0.2% |
| renal clear cell carcinoma survival | 38.59 | 1.48E-03 | 1 | 23 | 4.3% |
| occupational pesticide exposure | 31.44 | 2.16E-03 | 1 | 28 | 3.6% |
| Kabuki syndrome (KS) | 2.70 | 2.26E-03 | 6 | 1891 | 0.3% |
| maternal socioeconomic status | 29.28 | 2.46E-03 | 1 | 30 | 3.3% |
| preterm birth | 1.60 | 2.75E-03 | 19 | 10662 | 0.2% |
| breastfeeding | 6.79 | 3.26E-03 | 2 | 252 | 0.8% |
| folic acid and vitamin B12 supplementation | 6.25 | 4.37E-03 | 2 | 274 | 0.7% |
| type 2 diabetes (T2D) | 1.81 | 5.43E-03 | 12 | 5633 | 0.2% |
| SETD1B-related syndrome | 2.19 | 6.49E-03 | 7 | 2721 | 0.3% |
| maternal Hepatitis B virus (HBV) infection | 5.15 | 8.47E-03 | 2 | 332 | 0.6% |
| myalgic encephalomyelitis/chronic fatigue syndrome | 1.70 | 8.58E-03 | 13 | 6500 | 0.2% |
| IL-13 treatment | 2.83 | 8.78E-03 | 4 | 1203 | 0.3% |
| **Cluster 2** | | | | | |
| glucocorticoid exposure | 50.00 | 1.00E-308 | 92 | 3468 | 2.7% |
| t(1;11) translocation | 72.86 | 1.92E-11 | 1 | 13 | 7.7% |
| substance-use risk | 15.93 | 2.09E-05 | 1 | 65 | 1.5% |
| treated with assisted reproductive technology | 10.24 | 1.63E-04 | 1 | 118 | 0.8% |
| maternal hypertensive disorders in pregnancy | 3.06 | 9.42E-03 | 1 | 922 | 0.1% |

#### **Table S11.** EWAS atlas enrichment analysis results for all CpGs selected into the multi-block sparse Partial Least Squares Discriminant Analysis (MB-sPLS-DA) multi-omics model including the empirical design matrix.

Enriched traits based on enrichment analysis with 746 CpGs selected by the 5-component MB-sPLS-DA model including the empirical design matrix. The fourth column (DMC) shows how many of the 746 CpGs have been previously associated with the trait in the first column. The fifth column (background) shows how many CpGs have previously been associated with the trait in column 1. The last column (%) shows the percentage of CpGs previously associated with the trait in column 1 that were also selected by the MB- sPLS-DA model.

| **trait** | **OR** | ***p*** | **DMC** | **Background** | **%** |
| --- | --- | --- | --- | --- | --- |
| gender | 3.90 | 3.97E-31 | 54 | 15433 | 0.3% |
| breast cancer risk | 342.60 | 1.44E-21 | 5 | 24 | 20.8% |
| Claes-Jensen syndrome | 15.24 | 3.55E-20 | 12 | 1050 | 1.1% |
| Parkinson's disease (PD) | 38.92 | 4.14E-20 | 8 | 278 | 2.9% |
| Klinefelter syndrome | 45.38 | 3.19E-16 | 6 | 179 | 3.4% |
| down syndrome | 2.69 | 2.09E-13 | 32 | 14649 | 0.2% |
| Kabuki syndrome (KS) | 7.69 | 9.27E-13 | 11 | 1891 | 0.6% |
| respiratory allergies (RA) | 16.39 | 3.19E-11 | 6 | 485 | 1.2% |
| neurodevelopmental presentations and congenital anomalies (ND/CAs) | 10.80 | 1.62E-10 | 7 | 856 | 0.8% |
| gestational diabetes mellitus | 3.59 | 2.98E-10 | 18 | 6599 | 0.3% |
| household socioeconomic status in childhood | 10.63 | 7.35E-08 | 5 | 620 | 0.8% |
| ancestry | 2.43 | 4.03E-07 | 21 | 10618 | 0.2% |
| severe acute malnutrition | 260.23 | 4.94E-05 | 1 | 6 | 16.7% |
| preterm birth | 2.06 | 7.29E-05 | 14 | 10662 | 0.1% |
| Alzheimer's disease (AD) | 4.70 | 8.25E-05 | 5 | 1392 | 0.4% |
| ankylosis spondylitis (AS) | 15.99 | 1.43E-04 | 2 | 165 | 1.2% |
| leukoaraiosis (LA) | 7.41 | 2.08E-04 | 3 | 531 | 0.6% |
| Werner syndrome | 4.81 | 3.54E-04 | 4 | 1088 | 0.4% |
| obesity | 2.08 | 6.39E-04 | 13 | 8134 | 0.2% |
| Amyloid-β plaques | 10.55 | 6.68E-04 | 2 | 249 | 0.8% |
| Coffin-Siris syndrome (CSS) | 9.79 | 8.75E-04 | 2 | 268 | 0.7% |
| occupational pesticide exposure | 48.13 | 9.34E-04 | 1 | 28 | 3.6% |
| maternal socioeconomic status | 44.87 | 1.07E-03 | 1 | 30 | 3.3% |
| waist circumference (WC) | 5.12 | 1.38E-03 | 3 | 766 | 0.4% |
| autism spectrum disorders (ASD) | 5.11 | 1.40E-03 | 3 | 768 | 0.4% |
| maternal Hepatitis B virus (HBV) infection | 7.89 | 1.91E-03 | 2 | 332 | 0.6% |
| atopy | 2.10 | 2.20E-03 | 10 | 6198 | 0.2% |
| type 2 diabetes (T2D) | 2.08 | 3.88E-03 | 9 | 5633 | 0.2% |
| organophosphate exposure | 18.85 | 5.48E-03 | 1 | 70 | 1.4% |
| psoriasis | 2.20 | 6.21E-03 | 7 | 4138 | 0.2% |
| folic acid supplement during pregnancy | 5.56 | 6.47E-03 | 2 | 470 | 0.4% |
| facial aging | 16.90 | 6.73E-03 | 1 | 78 | 1.3% |
| maternal stress | 14.96 | 8.47E-03 | 1 | 88 | 1.1% |

#### **Table S12**. Prediction parameters of the multi-block sparse Partial Least Squares Discriminant Analyses (MB-sPLS-DA) models in the test and clinical data.

Balanced error rates (BER), prediction sensitivity, specificity, and accuracy, and model Area Under the Curve (AUC) of the prediction of the MB-sPLS-DA models in the test and clinical data per component. The overall AUC is reported per component for the MB-sPLS-DA models, as well as for the metabolomics, DNA methylation, and polygenic score blocks included in the MB-sPLS-DA models separately. The AUC *p*-values have been adjusted separately for the test and clinical data for multiple testing using the FDR of 5% for 48 tests (*q*).

|  |  |  |  |  |  | **AUC** | | | **Metabolomics** | | | **DNA methylation** | | | **Polygenic scores** | | |
| --- | --- | --- | --- | --- | --- | --- | --- | --- | --- | --- | --- | --- | --- | --- | --- | --- | --- |
| **Design matrix** | **component** | **BER** | **Sensitivity** | **Specificity** | **Accuracy** | **AUC** | ***p*** | ***q*** | **AUC** | ***p*** | ***q*** | **AUC** | ***p*** | ***q*** | **AUC** | ***p*** | ***q*** |
| **Test data** | | | | | | | | | | | | | | | | | |
| Empirical | 1 | 0.51 | 41.9 | 56.9 | 50.5 | 0.63 | 7.43E-05 | 3.57E-04 | 0.49 | 0.80 | 0.88 | 0.60 | 0.01 | 0.03 | 0.47 | 0.37 | 0.80 |
| Empirical | 2 | 0.49 | 46.2 | 55.6 | 51.6 | 0.67 | 7.27E-08 | 4.36E-07 | 0.51 | 0.85 | 0.89 | 0.49 | 0.73 | 0.88 | 0.51 | 0.76 | 0.88 |
| Empirical | 3 | 0.51 | 46.2 | 52.5 | 49.8 | 0.69 | 6.80E-09 | 4.67E-08 | 0.51 | 0.87 | 0.89 | 0.50 | 0.95 | 0.95 | 0.51 | 0.73 | 0.88 |
| Empirical | 4 | 0.52 | 45.3 | 50.6 | 48.4 | 0.73 | 1.43E-10 | 1.14E-09 | 0.53 | 0.45 | 0.85 | 0.48 | 0.56 | 0.88 | 0.53 | 0.42 | 0.85 |
| Empirical | 5 | 0.47 | 51.3 | 54.4 | 53.1 | 0.76 | 9.61E-11 | 1.14E-09 | 0.56 | 0.12 | 0.33 | 0.48 | 0.61 | 0.88 | 0.53 | 0.46 | 0.85 |
| Null | 1 | 0.48 | 48.7 | 55.0 | 52.3 | 0.65 | 2.19E-07 | 1.17E-06 | 0.48 | 0.49 | 0.87 | 0.58 | 0.02 | 0.07 | 0.52 | 0.51 | 0.88 |
| Null | 2 | 0.50 | 46.2 | 54.4 | 50.9 | 0.71 | 1.35E-10 | 1.14E-09 | 0.51 | 0.74 | 0.88 | 0.48 | 0.65 | 0.88 | 0.51 | 0.70 | 0.88 |
| Null | 3 | 0.49 | 41.0 | 61.9 | 53.1 | 0.77 | 2.13E-12 | 3.40E-11 | 0.53 | 0.42 | 0.85 | 0.52 | 0.62 | 0.88 | 0.51 | 0.72 | 0.88 |
| Null | 4 | 0.51 | 39.3 | 59.4 | 50.9 | 0.79 | 1.72E-12 | 3.40E-11 | 0.54 | 0.26 | 0.62 | 0.52 | 0.64 | 0.88 | 0.51 | 0.80 | 0.88 |
| Null | 5 | 0.50 | 40.2 | 60.6 | 52.0 | 0.79 | 1.41E-12 | 3.40E-11 | 0.55 | 0.16 | 0.41 | 0.51 | 0.86 | 0.89 | 0.51 | 0.78 | 0.88 |
| Full | 1 | 0.47 | 52.1 | 53.1 | 52.7 | 0.57 | 1.35E-01 | 3.61E-01 | 0.48 | 0.59 | 0.88 | 0.59 | 0.01 | 0.04 | 0.58 | 0.03 | 0.09 |
| Full | 2 | 0.46 | 50.4 | 57.5 | 54.5 | 0.59 | 1.20E-02 | 4.42E-02 | 0.49 | 0.79 | 0.88 | 0.56 | 0.11 | 0.33 | 0.54 | 0.30 | 0.68 |
| **Clinical data** | | | | | | | | | | | | | | | | | |
| Empirical | 1 | 0.53 | 42.2 | 52.6 | 47.9 | 0.63 | 7.43E-05 | 3.57E-04 | 0.46 | 0.36 | 0.54 | 0.44 | 0.26 | 0.40 | 0.42 | 0.08 | 0.23 |
| Empirical | 2 | 0.56 | 37.5 | 51.3 | 45.1 | 0.67 | 7.27E-08 | 4.36E-07 | 0.47 | 0.60 | 0.81 | 0.44 | 0.19 | 0.30 | 0.42 | 0.08 | 0.23 |
| Empirical | 3 | 0.58 | 43.8 | 39.7 | 41.5 | 0.69 | 6.80E-09 | 4.67E-08 | 0.46 | 0.45 | 0.63 | 0.42 | 0.09 | 0.23 | 0.41 | 0.07 | 0.22 |
| Empirical | 4 | 0.57 | 39.1 | 46.2 | 43.0 | 0.73 | 1.43E-10 | 1.14E-09 | 0.51 | 0.80 | 0.90 | 0.41 | 0.05 | 0.19 | 0.43 | 0.14 | 0.30 |
| Empirical | 5 | 0.57 | 37.5 | 48.7 | 43.7 | 0.76 | 9.61E-11 | 1.14E-09 | 0.52 | 0.75 | 0.89 | 0.40 | 0.05 | 0.19 | 0.43 | 0.15 | 0.30 |
| Null | 1 | 0.52 | 46.9 | 48.7 | 47.9 | 0.65 | 2.19E-07 | 1.17E-06 | 0.43 | 0.18 | 0.30 | 0.50 | 0.97 | 0.99 | 0.43 | 0.17 | 0.30 |
| Null | 2 | 0.50 | 48.4 | 51.3 | 50.0 | 0.71 | 1.35E-10 | 1.14E-09 | 0.52 | 0.74 | 0.89 | 0.49 | 0.80 | 0.90 | 0.43 | 0.16 | 0.30 |
| Null | 3 | 0.51 | 42.2 | 56.4 | 50.0 | 0.77 | 2.13E-12 | 3.40E-11 | 0.52 | 0.76 | 0.89 | 0.46 | 0.40 | 0.58 | 0.43 | 0.15 | 0.30 |
| Null | 4 | 0.51 | 42.2 | 56.4 | 50.0 | 0.79 | 1.72E-12 | 3.40E-11 | 0.51 | 0.90 | 0.96 | 0.47 | 0.61 | 0.81 | 0.43 | 0.15 | 0.30 |
| Null | 5 | 0.50 | 42.2 | 57.7 | 50.7 | 0.79 | 1.41E-12 | 3.40E-11 | 0.50 | 0.94 | 0.98 | 0.48 | 0.66 | 0.84 | 0.43 | 0.18 | 0.30 |
| Full | 1 | 0.48 | 32.8 | 71.8 | 54.2 | 0.57 | 1.35E-01 | 2.98E-01 | 0.42 | 0.08 | 0.23 | 0.57 | 0.16 | 0.30 | 0.50 | 1.00 | 1.00 |
| Full | 2 | 0.50 | 50.0 | 50.0 | 50.0 | 0.59 | 1.20E-02 | 5.22E-02 | 0.48 | 0.67 | 0.84 | 0.58 | 0.12 | 0.30 | 0.49 | 0.85 | 0.93 |

#### **Table S13.** Correlational patterns with high cross-omics correlations of the multi-omics variables identified in the multi-block sparse Partial Least Squares Discriminant Analysis (MB-sPLS-DA) model including the empirical design matrix.

Here, only high absolute correlations of the PLS variates (*r* ≥ 0.60) between variables of at least two omics blocks of the 5-component MB-sPLS-DA model with empirical design matrix are given. For each CpG, column 5 lists with which polygenic score (PGS) or metabolite the high correlation is observed, and column 6 indicates in which correlation pattern this correlation falls. All CpGs were looked up in the EWAS atlas, for all CpGs included in the EWAS atlas, column 7 lists the trait(s) this CpG has previously been associated with according to the EWAS atlas. If the CpG was not listed in the EWAS atlas and the CpG was located in a gene, this gene was looked up in the EWAS atlas, the final column gives the top traits this gene has been associated with in the EWAS atlas and the number of associations for the gene-trait combination. The annotation (genome build 37) of the selected CpG is provided, including the chromosome-base pair position, and the gene(s) in which the CpG is located. For the polygenic scores (PGSs), Attention-Deficit Hyperactivity Disorder is abbreviated as “ADHD”, and Educational Attainment as “EA”, and the ‘_NTf’ suffix denotes the PGSs non-transmitted by father. For the metabolites, the ‘amines.’ prefix indicates these metabolites were measured on the Liquid Chromatography Mass Spectrometry (LC-MS) amines platform, and the ‘OA.’ prefix indicates these metabolites were measured on the Gas Chromatography (GC-) MS organic acids platform. The full correlation matrix is included in **Data S10**.

| **CpGs** | **Chr** | **Position** | **Gene** | **PGSs/metabolites** | **Correlation sign** | **Pattern** | **EWAS atlas trait (CpG)** | **EWAs atlas trait (*N* associations / gene)** |
| --- | --- | --- | --- | --- | --- | --- | --- | --- |
| cg12886033 | 1 | 65449013 | LINC01359 | OA.Citric.acid | negative | 1 |  |  |
|  |  |  |  | OA.Fumaric.acid |  |  |  |  |
| cg14508705 | 1 | 172360182 | DNM3 | OA.Citric.acid | negative | 1 |  | Hepatocellular carcinoma (HCC) (15); Smoking (8); aging (6); Systemic lupus erythematosus (SLE) (4); Down syndrome (4) |
|  |  |  |  | OA.Fumaric.acid |  |  |  |  |
| cg15841349 | 12 | 129348564 | GLT1D1 | OA.Fumaric.acid | negative | 1 |  | Colorectal laterally spreading tumor (5); Oral squamous cell carcinoma (OSCC) (4); Aging (3); Smoking (3); Breast cancer prognosis (3) |
| cg21432062 | 3 | 4908643 |  | OA.Fumaric.acid | negative | 1 | Inflammatory bowel disease |  |
| cg11710553 | 4 | 105892960 |  | OA.Citric.acid | negative | 1 |  |  |
|  |  |  |  | OA.Fumaric.acid |  |  |  |  |
| cg22848658 | 6 | 135354586 | HBS1L | OA.Fumaric.acid | negative | 1 |  | Aging (2); Fractional exhaled nitric oxide (2); Primary Sjögren's Syndrome (pSS) (2); Myalgic encephalomyelitis/chronic fatigue syndrome (2); End-stage kidney disease attributed to diabetic kidney disease (1) |
| cg20704654 | 20 | 30072118 | NCRNA00028  REM1 | OA.Isocitrate | negative | 2 | Aging; Gender |  |
| cg11206167 | 5 | 42924367 |  | OA.Isocitrate | negative | 2 | Gender |  |
| cg05056638 | 8 | 24800824 |  | OA.Isocitrate | negative | 2 | Gender; In utero arsenic exposure; Leukoaraiosis |  |
| cg08415582 | 8 | 57030523 |  | OA.Isocitrate | negative | 2 | Gender |  |
| cg13784456 | 10 | 132970405 | TCERG1L | amines.Homocysteine | positive | 3 |  | B Acute Lymphoblastic Leukemia with t(1;19)(q23;p13.3) (14); Infertility (12); Hepatocellular carcinoma (HCC) (10); Colorectal laterally spreading tumor (5); Smoking (4) |
| cg06144718 | 10 | 133048392 | TCERG1L | amines.Homocysteine | positive | 3 |  | B Acute Lymphoblastic Leukemia with t(1;19)(q23;p13.3) (14); Infertility (12); Hepatocellular carcinoma (HCC) (10); Colorectal laterally spreading tumor (5); Smoking (4) |
| cg03469862 | 11 | 68924853 |  | ADHD | positive | 4 | Prostate cancer; Pre- and post lenalidomide treatment in patients with myelodysplastic syndrome with isolated deletion (5q) |  |
| cg03469862 | 11 | 68924853 |  | amines.3.Methoxytyrosine | positive | 4 | Prostate cancer; Pre- and post lenalidomide treatment in patients with myelodysplastic syndrome with isolated deletion (5q) |  |
|  |  |  |  | amines.L.Glutamine |  |  |  |  |
|  |  |  |  | amines.L.Isoleucine |  |  |  |  |
|  |  |  |  | amines.L.Leucine |  |  |  |  |
|  |  |  |  | amines.L.Phenylalanine |  |  |  |  |
|  |  |  |  | amines.L.Serine |  |  |  |  |
|  |  |  |  | amines.L.Tryptophan |  |  |  |  |
|  |  |  |  | amines.L.Tyrosine |  |  |  |  |
|  |  |  |  | amines.L.Valine |  |  |  |  |
| cg09674340 | 1 | 202509286 | PPP1R12B | EA_NTf | negative | 5 |  | Gender (10); Fractional exhaled nitric oxide (3); Allergic sensitization (2); Low birth weight (2); Smoking (2) |

#### **Table S14.** EWAS atlas enrichment analysis results for all CpGs selected into the multi-block sparse Partial Least Squares Discriminant Analysis (MB-sPLS-DA) multi-omics model including a null design matrix.

Enriched traits based on enrichment analysis with 1,831 CpGs selected by the 5-component MB- sPLS-DA model including a null design matrix. The fourth column (DMC) shows how many of the 1,831 CpGs have been previously associated with the trait in the first column. The fifth column (background) shows how many CpGs have previously been associated with the trait in column 1. The last column (%) shows the percentage of CpGs previously associated with the trait in column 1 that were also selected by the MB-sPLS-DA model.

| **Trait** | **OR** | ***p*** | **DMC** | **Background** | **%** |
| --- | --- | --- | --- | --- | --- |
| Claes-Jensen syndrome | 13.58 | 3.65E-39 | 26 | 1050 | 2.5% |
| gender | 2.81 | 4.11E-35 | 103 | 15433 | 0.7% |
| Klinefelter syndrome | 41.66 | 5.27E-32 | 13 | 179 | 7.3% |
| ancestry | 3.15 | 4.77E-28 | 67 | 10618 | 0.6% |
| respiratory allergies (RA) | 13.48 | 5.94E-19 | 12 | 485 | 2.5% |
| breast cancer risk | 139.97 | 1.10E-17 | 5 | 24 | 20.8% |
| household socioeconomic status in childhood | 9.59 | 1.34E-14 | 11 | 620 | 1.8% |
| Kabuki syndrome (KS) | 5.11 | 1.56E-14 | 18 | 1891 | 1.0% |
| gestational diabetes mellitus | 2.83 | 1.57E-13 | 35 | 6599 | 0.5% |
| down syndrome | 2.01 | 1.88E-13 | 65 | 14649 | 0.4% |
| Parkinson's disease (PD) | 13.70 | 8.84E-12 | 7 | 278 | 2.5% |
| Alzheimer's disease (AD) | 5.00 | 9.26E-11 | 13 | 1392 | 0.9% |
| facial anomalies syndrome (ICF) | 15.05 | 3.42E-09 | 5 | 181 | 2.8% |
| systemic lupus erythematosus (SLE) | 2.24 | 8.47E-09 | 33 | 7848 | 0.4% |
| neurodevelopmental presentations and congenital anomalies (ND/CAs) | 5.63 | 1.14E-08 | 9 | 856 | 1.1% |
| human herpesvirus 6B infection | 7.94 | 8.83E-08 | 6 | 406 | 1.5% |
| follicular thyroid carcinoma | 2.29 | 3.99E-07 | 24 | 5575 | 0.4% |
| SETD1B-related syndrome | 2.74 | 3.98E-06 | 14 | 2721 | 0.5% |
| thyroid lesion | 3.04 | 2.16E-05 | 10 | 1753 | 0.6% |
| papillary thyroid carcinoma | 1.92 | 7.00E-05 | 22 | 6073 | 0.4% |
| tooth mobility | 18.89 | 8.34E-05 | 2 | 58 | 3.4% |
| folic acid supplement during pregnancy | 4.54 | 5.21E-04 | 4 | 470 | 0.9% |
| Coffin<U+2013>Siris syndrome (CSS) | 5.99 | 6.42E-04 | 3 | 268 | 1.1% |
| osteoarthritis (OA) | 2.60 | 7.02E-04 | 8 | 1634 | 0.5% |
| sperm motility | 48.04 | 1.08E-03 | 1 | 12 | 8.3% |
| maternal smoking | 1.72 | 1.12E-03 | 20 | 6153 | 0.3% |
| leukoaraiosis (LA) | 4.01 | 1.13E-03 | 4 | 531 | 0.8% |
| atopy | 1.71 | 1.27E-03 | 20 | 6198 | 0.3% |
| food allergy | 8.26 | 1.67E-03 | 2 | 130 | 1.5% |
| prenatal phthalate exposure | 3.13 | 1.83E-03 | 5 | 850 | 0.6% |
| ankylosis spondylitis (AS) | 6.49 | 3.89E-03 | 2 | 165 | 1.2% |
| occupational pesticide exposure | 19.59 | 5.40E-03 | 1 | 28 | 3.6% |
| primary Sjögren’s Syndrome (pSS) | 1.80 | 5.56E-03 | 12 | 3526 | 0.3% |
| maternal socioeconomic status | 18.23 | 6.15E-03 | 1 | 30 | 3.3% |
| adrenocortical carcinoma | 2.34 | 6.95E-03 | 6 | 1364 | 0.4% |
| type 2 diabetes (T2D) | 1.60 | 7.27E-03 | 17 | 5633 | 0.3% |
| preterm birth | 1.40 | 7.52E-03 | 31 | 10662 | 0.3% |
| fruit consumption | 2.88 | 8.08E-03 | 4 | 739 | 0.5% |

#### **Table S15.** EWAS atlas enrichment analysis results for the CpGs included in correlation pattern 2 and 3 of the multi-block sparse Partial Least Squares Discriminant Analysis (MB-sPLS-DA) model including a null design matrix.

Enriched traits based on enrichment analysis with 49 CpGs that were included in correlation pattern 2 and 299 CpGs that were included in correlation pattern 3 of the 5-component MB-sPLS-DA model including a null design matrix. Correlation pattern assignment assignment is provided in **Data S13**. The fourth column (DMC) shows how many of the CpGs have been previously associated with the trait in the first column. The fifth column (background) shows how many CpGs have previously been associated with the trait in column 1. The last column (%) shows the percentage of CpGs previously associated with the trait in column 1 that were also included in correlation pattern 2 or 3 of the MB-sPLS-DA model including a null design matrix.

| **Trait** | **OR** | ***p*** | **DMC** | **Background** | **%** |
| --- | --- | --- | --- | --- | --- |
| **Correlation pattern 2** | | | | | |
| prenatal arsenic exposure | 5.21 | 4.22E-08 | 9 | 4860 | 0.2% |
| tooth mobility | 99.09 | 1.30E-07 | 2 | 58 | 3.4% |
| prenatal paracetamol exposure | 5.98 | 8.53E-05 | 4 | 1866 | 0.2% |
| Behcet's disease | 3.84 | 1.21E-04 | 6 | 4364 | 0.1% |
| folic acid supplement | 55.42 | 6.79E-04 | 1 | 51 | 2.0% |
| vitamin B12 supplement | 9.46 | 9.90E-04 | 2 | 589 | 0.3% |
| endometrial carcinoma | 44.69 | 1.03E-03 | 1 | 63 | 1.6% |
| myopia | 5.42 | 1.05E-03 | 3 | 1541 | 0.2% |
| obesity | 2.39 | 3.34E-03 | 7 | 8134 | 0.1% |
| polycystic ovary syndrome (PCOS) | 20.37 | 4.65E-03 | 1 | 137 | 0.7% |
| asthma | 1.81 | 6.18E-03 | 8 | 13639 | 0.1% |
| mortality | 2.81 | 9.42E-03 | 4 | 3951 | 0.1% |
| **Correlation pattern 3** | | | | | |
| tooth mobility | 116.67 | 6.87E-08 | 2 | 58 | 3.4% |
| prenatal arsenic exposure | 5.44 | 1.32E-07 | 8 | 4860 | 0.2% |
| Behcet's disease | 4.52 | 2.69E-05 | 6 | 4364 | 0.1% |
| prenatal paracetamol exposure | 7.04 | 2.82E-05 | 4 | 1866 | 0.2% |
| myopia | 6.37 | 4.62E-04 | 3 | 1541 | 0.2% |
| folic acid supplement | 65.09 | 4.94E-04 | 1 | 51 | 2.0% |
| vitamin B12 supplement | 11.12 | 5.48E-04 | 2 | 589 | 0.3% |
| endometrial carcinoma | 52.54 | 7.48E-04 | 1 | 63 | 1.6% |
| obesity | 2.81 | 7.83E-04 | 7 | 8134 | 0.1% |
| polycystic ovary syndrome (PCOS) | 23.94 | 3.40E-03 | 1 | 137 | 0.7% |
| mortality | 3.31 | 3.76E-03 | 4 | 3951 | 0.1% |
| asthma | 1.95 | 3.84E-03 | 7 | 13639 | 0.1% |
| maternal alcohol consumption | 3.71 | 6.66E-03 | 3 | 2640 | 0.1% |
| rheumatoid arthritis (RA) | 2.84 | 9.04E-03 | 4 | 4602 | 0.1% |

#### **Table S16.** EWAS atlas enrichment analysis results for all CpGs selected into the multi-block sparse Partial Least Squares Discriminant Analysis (MB-sPLS-DA) multi-omics model including a full design matrix.

Enriched traits based on enrichment analysis with 65 CpGs selected by the 2-component MB- sPLS-DA model with a full design matrix. The fourth column (DMC) shows how many of the 65 CpGs have been previously associated with the trait in the first column. The fifth column (background) shows how many CpGs have previously been associated with the trait in column 1. The last column (%) shows the percentage of CpGs previously associated with the trait in column 1 that were also selected by the MB-sPLS-DA model including a full design matrix.

| **Trait** | **OR** | ***p*** | **DMC** | **Background** | **%** |
| --- | --- | --- | --- | --- | --- |
| glucocorticoid exposure | 50.61 | 6.63E-26 | 10 | 3468 | 0.3% |
| primary Sjögren’s Syndrome (pSS) | 8.68 | 1.48E-03 | 2 | 3526 | 0.1% |
| maternal phthalate exposure | 36.53 | 1.52E-03 | 1 | 416 | 0.2% |

#### **Table S17.** Correlational patterns with high cross-omics correlations of the multi-omics variables identified in the multi-block sparse Partial Least Squares Discriminant Analysis (MB-sPLS-DA) model including a full design matrix.

Here, only high absolute correlations of the PLS variates (*r* ≥ 0.60) between variables of at least two omics blocks in the 2-component MB-sPLS-DA model with a full design matrix are given. For each CpG or polygenic score (PGS), column 5 lists with which metabolite or PGS the high correlation is observed, and column 6 indicates in which correlation pattern this correlation falls. All CpGs were looked up in the EWAS atlas, for all CpGs included in the EWAS atlas, column 7 lists the trait(s) this CpG has previously been associated with according to the EWAS atlas. If the CpG was not listed in the EWAS atlas and the CpG was located in a gene, this gene was looked up in the EWAS atlas, the final column gives the top traits this gene has been associated with in the EWAS atlas and the number of associations for the gene-trait combination. The annotation (genome build 37) of the selected CpG is provided, including the chromosome-base pair position, and the gene(s) in which the CpG is located. For the polygenic scores (PGSs), Attention-Deficit Hyperactivity Disorder is abbreviated as “ADHD”, and Educational Attainment as “EA”, and the ‘_NTf’ suffix denotes the PGSs non-transmitted by father. For the metabolites, the ‘amines.’ prefix indicates these metabolites were measured on the Liquid Chromatography Mass Spectrometry (LC-MS) amines platform, and the ‘OA.’ prefix indicates these metabolites were measured on the Gas Chromatography (GC-) MS organic acids platform. The full correlation matrix is included in **Data S15**.

| **PGSs/CPGs** | **Chr** | **Position** | **Gene** | **Metabolites/PGSs** | **Correlation sign** | **Pattern** | **EWAS atlas trait (CpG)** | **EWAS atlas (*N* associations / gene)** |
| --- | --- | --- | --- | --- | --- | --- | --- | --- |
| ADHD_NTf |  |  |  | amines.Glutathione | positive | 1 |  |  |
|  |  |  |  | amines.3.Methoxytyramine |  |  |  |  |
|  |  |  |  | amines.Methionine.sulfone |  |  |  |  |
|  |  |  |  | amines.3.Methoxytyrosine |  |  |  |  |
|  |  |  |  | amines.O.Acetyl.L.serine |  |  |  |  |
|  |  |  |  | amines.5.Hydroxy.L.tryptophan |  |  |  |  |
|  |  |  |  | amines.SDMA |  |  |  |  |
|  |  |  |  | amines.ADMA |  |  |  |  |
|  |  |  |  | amines.Serotonine |  |  |  |  |
|  |  |  |  | amines.Cysteine |  |  |  |  |
|  |  |  |  | amines.Ethanolamine |  |  |  |  |
|  |  |  |  | amines.L.Tryptophan |  |  |  |  |
|  |  |  |  | amines.Gamma.aminobutyric.acid |  |  |  |  |
|  |  |  |  | amines.L.Valine |  |  |  |  |
|  |  |  |  | amines.L.Glutamine |  |  |  |  |
|  |  |  |  | amines.L.Isoleucine |  |  |  |  |
|  |  |  |  | amines.L.Tyrosine |  |  |  |  |
|  |  |  |  | amines.L.Leucine |  |  |  |  |
|  |  |  |  | amines.L.Phenylalanine |  |  |  |  |
|  |  |  |  | amines.Norepinephrine |  |  |  |  |
|  |  |  |  | amines.L.Serine |  |  |  |  |
| cg05410331 | 9 | 115085154 | MIR3134  PTBP3 | amines.3.Methoxytyramine | positive | 1 |  | smoking (6); ancestry (2); Werner syndrome (1); mortality (1); aging (1) |
|  |  |  |  | amines.5.Hydroxy.L.tryptophan |  |  |  |  |
|  |  |  |  | amines.L.Serine |  |  |  |  |
|  |  |  |  | amines.Ethanolamine |  |  |  |  |
|  |  |  |  | amines.3.Methoxytyrosine |  |  |  |  |
|  |  |  |  | ADHD_NTf |  |  |  |  |
|  |  |  |  | amines.L.Valine |  |  |  |  |
|  |  |  |  | amines.L.Glutamine |  |  |  |  |
|  |  |  |  | amines.ADMA |  |  |  |  |
|  |  |  |  | amines.L.Isoleucine |  |  |  |  |
|  |  |  |  | amines.O.Acetyl.L.serine |  |  |  |  |
|  |  |  |  | amines.L.Leucine |  |  |  |  |
|  |  |  |  | amines.SDMA |  |  |  |  |
|  |  |  |  | amines.L.Phenylalanine |  |  |  |  |
|  |  |  |  | amines.Serotonine |  |  |  |  |
| cg21444670 | 17 | 57048710 | PPM1E | steroids.Cortisol | positive | 2 |  | aging (26); B Acute Lymphoblastic Leukemia with t(1;19)(q23;p13.3) (11); asthma (3); oral squamous cell carcinoma (OSCC) (2); down syndrome (1) |
| cg05153029 | 20 | 19769815 |  | steroids.Cortisol | positive | 2 | glucocorticoid exposure |  |
